# A Modeling Analysis on Eliminating Racial/Ethnic Disparities in HIV Incidence in the United States

**DOI:** 10.1101/2024.08.15.24312083

**Authors:** Evin Uzun Jacobson, Alex Viguerie, Laurel Bates, Katherine Hicks, Amanda A. Honeycutt, Justin Carrico, Cynthia Lyles, Paul G. Farnham

## Abstract

**Background:** Despite progress in HIV prevention and treatment, resulting in overall incidence reductions in the United States, large racial/ethnic (r/e) disparities in HIV incidence remain due to stigma, discrimination, racism, poverty, and other social and structural factors.

**Setting:** We used the HIV Optimization and Prevention Economics (HOPE) model to analyze which intervention strategies provide the most effective path towards eliminating r/e disparities in HIV incidence.

**Methods:** We considered four intervention scenarios for 2023-2035, which focused on eliminating r/e disparities by 2027 in the HIV care continuum only, HIV prevention services only, both continuum and prevention services, and a final scenario where prevention and care levels for Black and Hispanic/Latino were set to maximum feasible levels. The primary outcome is the incidence-rate-ratio (IRR) for Black and Hispanic/Latino populations compared to Other populations (of whom 89% are White) with the goal of IRRs ≤ 1 by 2035.

**Results:** All scenarios reduced IRRs but only *Maximum Feasible* eliminated HIV incidence disparities by 2035, with respective IRRs of 0.9 and 1.1 among the Black and Hispanic/Latino populations, compared to 6.5 and 4.1 in the baseline scenario. *Continuum-only* was more effective at reducing disparities (2035 IRRs of 4.7 for Black and 3.1 for Hispanic/Latino populations) than *Prevention-only* (6.1 and 3.7 respectively).

**Conclusions:** With no prioritized changes, our simulation showed that r/e disparities in HIV incidence persist through 2035. Elimination of r/e incidence disparities by 2035 is only possible if maximum HIV prevention and care levels for Black and Hispanic/Latino populations can be realized by 2027.

## Introduction

The *Ending the HIV Epidemic in the U.S. (EHE)* initiative, launched in 2019, aims to reduce annual new HIV infections by 90 percent in the United States by 2030. However, despite substantial progress in the prevention and treatment of HIV in the United States, certain subpopulations continue to comprise a disproportionate share of new HIV infections, particularly Black or African American (hereafter “Black”) and Hispanic/Latino populations (CDC, 2024a; CDC, 2024b). These racial and ethnic disparities in HIV result from stigma, discrimination, racism, poverty, and other social and structural factors and create significant challenges in achieving the goals of the EHE initiative (Alang & Blackstock et al., 2023; Dailey et al., 2023; Raiford et al., 2023; Bonacci et al., 2021). These underlying social and structural barriers make it difficult to realize equity with respect to access to and use of HIV prevention services, such as pre-exposure prophylaxis (PrEP) and syringe services programs (SSPs) (Chan, 2020, Kanny, 2019, Jenness, 2019, Bonacci, 2021, Handanagic, 2021, and Pyra, 2021); HIV testing and diagnosis (McCree, 2021, Lyons, 2021, and Hoover, 2021); and antiretroviral treatment to achieve viral suppression (Sullivan, 2021). Among Black and Hispanic/Latino populations, access to and use of HIV prevention and treatment services is almost universally lower than among all other racial and ethnic populations.

Prior modeling studies have analyzed whether scaling up effective HIV services could reduce disparities in service utilization and, over the long term, could reduce racial and ethnic disparities in HIV incidence (Nosyk, 2020a and 2020b; Quan, 2021). These studies focused on six distinct metropolitan areas in the United States and found that scaling up services in proportion to current levels of use by race and ethnicity is not sufficient to meaningfully reduce disparities in incidence. However, scaling up services in proportion to current HIV incidence rates by race and ethnicity can have a substantial impact on reducing, but not eliminating, racial and ethnic disparities in incidence. This impact varied among the six cities in this analysis.

In the current modeling study, we conducted simulations to explore whether and to what extent increasing HIV prevention and treatment services for Black and Hispanic/Latino populations could reduce or eliminate disparities in HIV incidence over time. Our simulations analyzed the impact on HIV incidence of eliminating the racial/ethnic disparities in HIV prevention and care outcomes by focusing on the HIV care continuum only, HIV prevention services only, or both. We also examined the corresponding spending levels for each intervention focus area and the spending increases necessary to eliminate disparities in incidence between Black, Hispanic/Latino, and “Other” (defined as all other racial ethnic populations, of whom 89% are white) populations.

## Methods

We used the HIV Optimization and Prevention Economics (HOPE) Model, a dynamic, compartmental model programmed in MATLAB (Mathworks, Natick, Massachusetts) that simulates the portion of the U.S. population aged 13 years and older that is sexually active and/or injects drugs (Khurana, 2018, Sansom, 2021, Chen 2022, Jacobson 2022). The population in HOPE is stratified into 273 subpopulations, including assigned sex at birth (M/F), age, HIV transmission, and racial/ethnic (r/e) groups (categorized as Black; Hispanic/Latino; and Other race/ethnicity). Persons with HIV (PWH) are distributed across 23 compartments, by HIV stage (uninfected, acute infection, 4 chronic states defined by CD4 levels), and by continuum-of-care status (undiagnosed, diagnosed, linked to care, on antiretroviral treatment [ART], and viral load suppressed). Supplement A describes the differential equations, model calculations, all model inputs and sources, and details on model calibration.

We modeled four scenarios of improving HIV care and prevention outcomes from 2023 through 2027 for Black and Hispanic/Latino populations with an analytic time period of 2023 through 2035. We compared HIV incidence outcomes from 2023 to 2035 from these scenarios to a baseline scenario that reflects a continuation of pre-2022 trends in continuum and prevention efforts from 2023 to 2035. This baseline scenario simulates what would be expected in the absence of additional expanded prevention and care services reaching Black and Hispanic/Latino populations. We assessed health disparities using HIV incidence rate ratios (IRRs) for Black and Hispanic/Latino populations relative to the Other population, which are calculated by dividing the annual incidence per 100,000 for Black or Hispanic/Latino populations by the annual incidence per 100,000 for the Other population; an IRR of one implies the incidence rates in the two groups are equal. Our research question was to determine whether and to what extent eliminating disparities in HIV prevention and care outcomes for Black and Hispanic/Latino populations could reduce or eliminate the racial/ethnic disparities in HIV incidence, with the goal of achieving HIV IRRs ≤ 1 for both Black and Hispanic/Latino populations by 2035.

The first three scenarios simulated improvements starting in 2023 so that, by 2027: the HIV care continuum levels for Black, Hispanic/Latino would equal the level for the Other population (*Continuum-only* scenario); HIV prevention levels, including PrEP and SSPs, would be equal (*Prevention-only* scenario); or both the HIV care continuum and prevention levels would be equal (*Continuum+Prevention* scenario), respectively. The fourth scenario simulated improvements so that HIV care continuum and prevention outcomes reached what we considered to be the maximum feasible levels for the Black and Hispanic/Latino populations by 2027 (*Maximum-feasible* scenario). In all 4 of these scenarios, baseline continuum and prevention efforts were maintained for the Other r/e group.

Before describing our intervention scenarios in detail, we note that in the years 2020-22, we modeled the effect of the coronavirus-2019 pandemic (COVID-19) on HIV by changing levels of testing (DiNenno 2022, Viguerie 2023a), ART use and adherence (Huang 2022, ATLAS), PrEP use and adherence (Zhu 2022, ATLAS), sexual activity levels (Dana 2023, Viguerie 2023b), and population mortality (Viguerie 2024). As our different intervention scenarios begin in 2023, COVID-19 effects are uniform for all scenarios. Complete details regarding the definition and implementation of COVID-19 effects on model inputs are provided in Supplement A.

We simulated each scenario in the HOPE model by adjusting input values for 2023 through 2035 (Table 8.1 in Supplement A), as follows:

### *Continuum-only* scenario

We adjusted r/e-specific rates for continuum-of-care progression so that all r/e groups achieved the same level of the group with the highest baseline level for the following three measures by 2027: percentage of PWH aware of their infection, percentage of diagnosed PWH linked to care (LTC), and percentage of diagnosed PWH virally suppressed (VS). For each measure, the Other r/e group had the highest baseline levels in 2022, with 95% aware, 90% LTC, and 74% VS (Figure 1, Table 8.2 in Supplement A). Starting in 2023, we adjusted the input values related to each measure, so that Black and Hispanic/Latino populations achieved similar levels as the Other r/e group by 2027 (Figure 1, Table 8.2 in Supplement A). In the *Continuum-only* scenario, we made no changes to existing prevention efforts (PrEP and SSPs).

**Figure 1:**
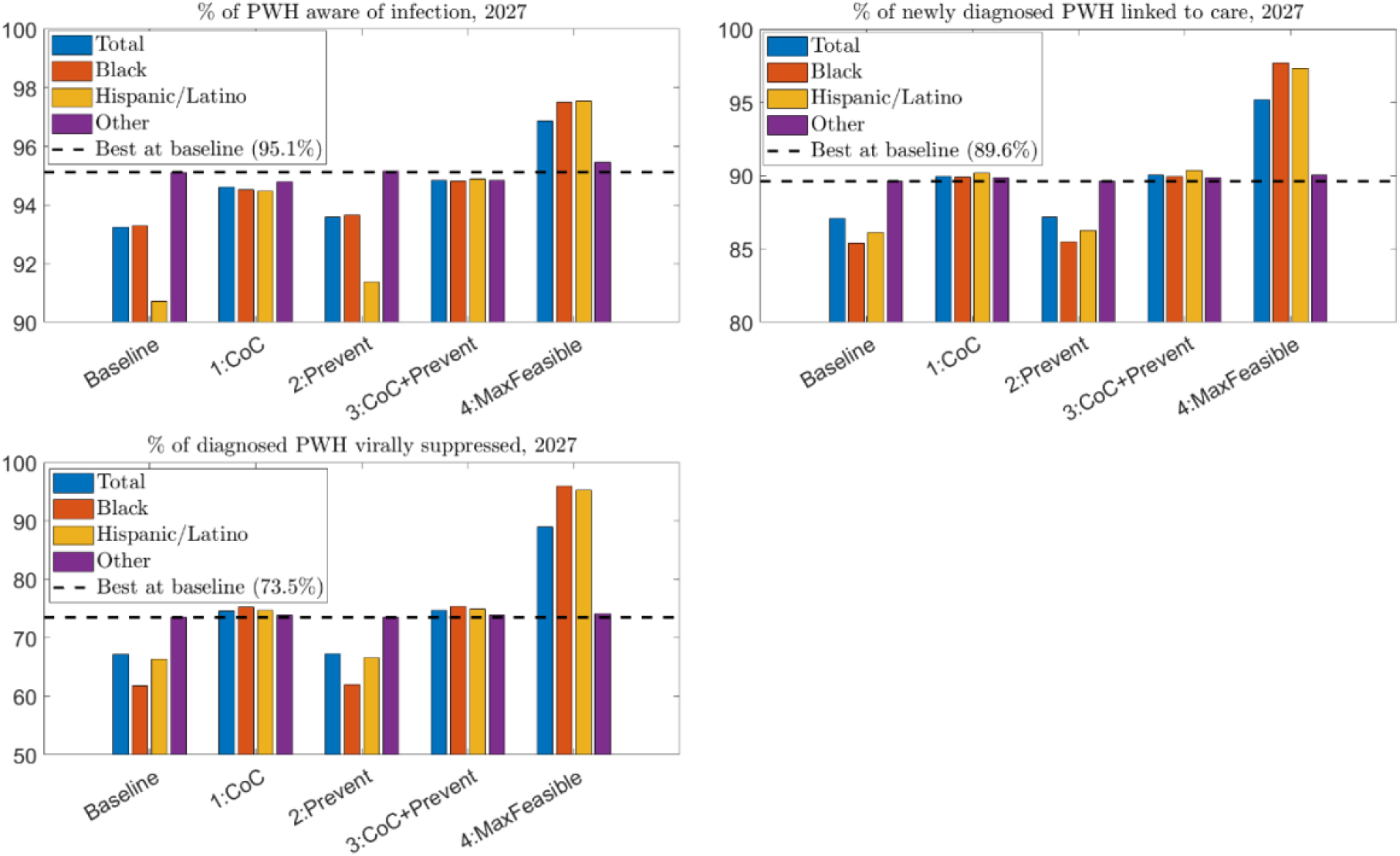
Key continuum-of-care indicators in 2027 for each scenario, by race/ethnicity

### *Prevention-only* scenario

We applied a similar procedure for the prevention interventions. We set r/e specific rates starting in 2023 for PrEP initiation and SSP participation, so that all r/e groups achieved the highest baseline levels across the 3 r/e groups for the percentage of uninfected people who inject drugs (PWID) served by SSPs and the percentage of PrEP-eligible people (as defined under 2017 CDC guidelines) using PrEP in 2027 (Table 8.2 in Supplement A). For both interventions, the Other r/e group had the highest baseline levels in 2027, with 88% of uninfected PWID in SSPs and 60% of PrEP-eligible using PrEP (Figure 2). We adjusted, starting in 2023, the number of uninfected PWID served by SSPs annually and the proportion of the eligible Black and Hispanic/Latino populations using PrEP, such that these populations achieved the same levels of SSP and PrEP utilization as the Other r/e group by 2027 (Figure 2). In the *Prevention-only* scenario, we made no changes to existing HIV care continuum efforts as compared to the baseline scenario. This analysis considered only oral PrEP, and did not consider alternative PrEP delivery methods, such as long-acting injectable PrEP.

**Figure 2:**
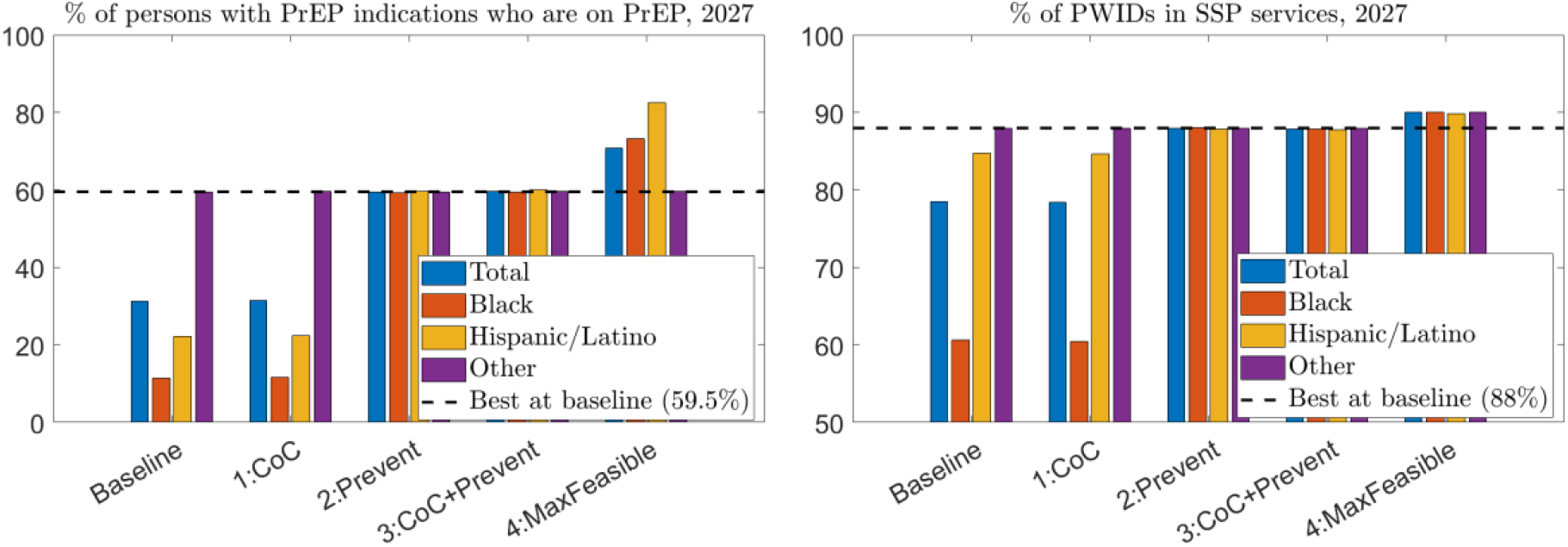
Key prevention indicators in 2027 for each scenario, by race/ethnicity

### *Continuum + Prevention* scenario

For Black and Hispanic/Latino populations, we set all continuum-of-care progression rates to the values used in the *<u>Continuum-only</u>* scenario and prevention participation rates to the values used in the *Prevention-only* scenario (Table 8.2 in Supplement A).

### *Maximum Feasible* scenario

We set rates for care continuum progression and prevention participation for Black and Hispanic/Latino populations to the maximum feasible levels. Specifically, rates were set so that the percent aware, percent LTC, and percent VS increased to approximately 98% by 2027.

Additionally, PrEP initiation and dropout rates were adjusted to increase the percentage of PrEP-eligible people in each r/e group on PrEP to 80% among men who have sex with men and to 50% among heterosexuals and PWID. SSP r/e-specific participation rates were set to achieve 90% of PWID in each r/e group in SSPs. The number of persons eligible for PrEP was obtained from AHEAD estimates (USDHHS, 2022). These input changes resulted in continuum-of-care and prevention outcome measures for Black and Hispanic/Latino populations that are higher than for the Other population (Table 8.2 in Supplement A; Figure 1; Figure 2).

We simulated these scenarios so that the HIV care continuum and/or prevention outcome goals were reached by 2027 but continued the simulations out to 2035 to allow for additional time to observe the impact on HIV incidence and, most importantly, the racial/ethnic-specific incidence rates.

We also estimated annual spending on continuum and prevention efforts and HIV treatment in the different scenarios as the number of modeled persons receiving interventions each year times per-unit intervention cost estimates. Per-person treatment costs were estimated from the literature and were applied to the number of PWH in each scenario. Further details, including complete unit cost estimates and data sources, are provided in Supplement A (Section 9).

## Results

By 2035, all four intervention scenarios resulted in lower incidence overall and for each r/e group compared to the baseline scenario. The *Continuum-only* scenario resulted in an overall incidence level of 9.1 new infections per 100,000 persons compared with a baseline incidence of 13.3 per 100,000 persons, while the *Prevention-only* scenario reduced incidence to 12.1 per 100,000 (Table 1). The additional benefit of increasing prevention efforts over the *Continuum-only* scenario in terms of reduced overall incidence in the *Continuum + Prevention* scenario was relatively small, decreasing to 8.4 new infections per 100,000 persons. The *Maximum Feasible* scenario resulted in a total incidence of 3.4 new infections per 100,000 persons for all racial/ethnic groups, practically eliminating all disparities among the groups.

**Table 1:** Incidence rate ratio and incidence per 100,000 population, by race/ethnicity, 2035.

| Outcome | Racial-ethnic group | Scenario |  |  |  |  |
| --- | --- | --- | --- | --- | --- | --- |
|  |  | Baseline | Continuum-only | Prevention-only | Continuum + Prevention | Maximum Feasible |
| Incidence rate ratio versus Other race/ethnicity | Black | 6.5 | 4.7 | 6.1 | 4.3 | 0.9 |
|  | Hispanic/Latino | 4.1 | 3.1 | 3.7 | 2.8 | 1.1 |
| Incidence per 100,000 people | All | 13.3 | 9.1 | 12.1 | 8.4 | 3.4 |
|  | Black | 37.0 | 22.6 | 33.5 | 20.2 | 3.3 |
|  | Hispanic/Latino | 23.4 | 14.9 | 20.4 | 13.0 | 3.7 |
|  | Other | 5.7 | 4.8 | 5.5 | 4.7 | 3.4 |

For the baseline scenario, we estimated that the IRR in 2035 would be 6.5 for Black and 4.1 for Hispanic / Latino populations when compared to the Other population. For the *Continuum-only, Prevention-only*, and *Continuum + Prevention* scenarios, racial/ethnic disparities in HIV incidence, as defined by the IRR, declined but were not eliminated by 2035 relative to the baseline (Table 1). The *Continuum-only* scenario showed a sizeable reduction in the IRRs versus the Other population for both Black and Hispanic/Latino populations compared with the baseline (4.7 for Black and 3.1 for Hispanic/Latino). In the *Prevention-only* scenario, there was a minor decrease in the IRR compared with baseline (6.1 for Black and 3.7 for Hispanic/Latino), indicating that the effect of eliminating HIV care continuum differences was larger than that of eliminating differences in HIV prevention levels. In the *Continuum + Prevention* scenario, the IRRs for Black and Hispanic/Latino populations versus Other declined to 4.3 and 2.8, respectively, although that reflected only a small reduction compared to the *Continuum-only* scenario. The *Maximum Feasible* scenario was the only scenario that resulted in the approximate elimination of racial/ethnic incidence disparities by 2035. The IRR for Black versus Other was 0.9, while the IRR for Hispanic/Latino versus Other was 1.1 in 2035 (Table 1).

The additional spending above baseline differed substantially across the intervention scenarios. The *Continuum-only* scenario required the least additional total spending at $9.5 billion (Table 2, Figure 3), with spending increases of $7.1 and $7.0 billion for expanding HIV care continuum services among Black and Hispanic/Latino populations, respectively, and a reduction in spending of $4.6 billion for Other populations. This reduction in spending was due to reductions in treatment costs from lower HIV incidence in the Other population resulting from the spillover effects of increased intervention efforts among the Black and Hispanic/Latino populations, coupled with no increased intervention spending for the Other population. The *Prevention-only* and *Continuum + Prevention* scenarios required about 4 and 5 times more additional spending than the *Continuum-only* Scenario, respectively, ($38.5 billion and $48.6 billion; Table 2, Figure 3). The *Maximum Feasible* scenario required a substantial increase of $162 billion in additional spending compared to the baseline scenario, including respective increases of $101.70 and $73.1 billion for expanding HIV care continuum and prevention services among Black and Hispanic/Latino populations. In each scenario, increased continuum and prevention spending among Black and Hispanic/Latino groups was partially offset by net reductions in treatment spending in all 3 r/e populations (not shown) resulting from reduced incidence in both the Black and Hispanic/Latino populations and spillover effects into the Other population.

**Table 2:**
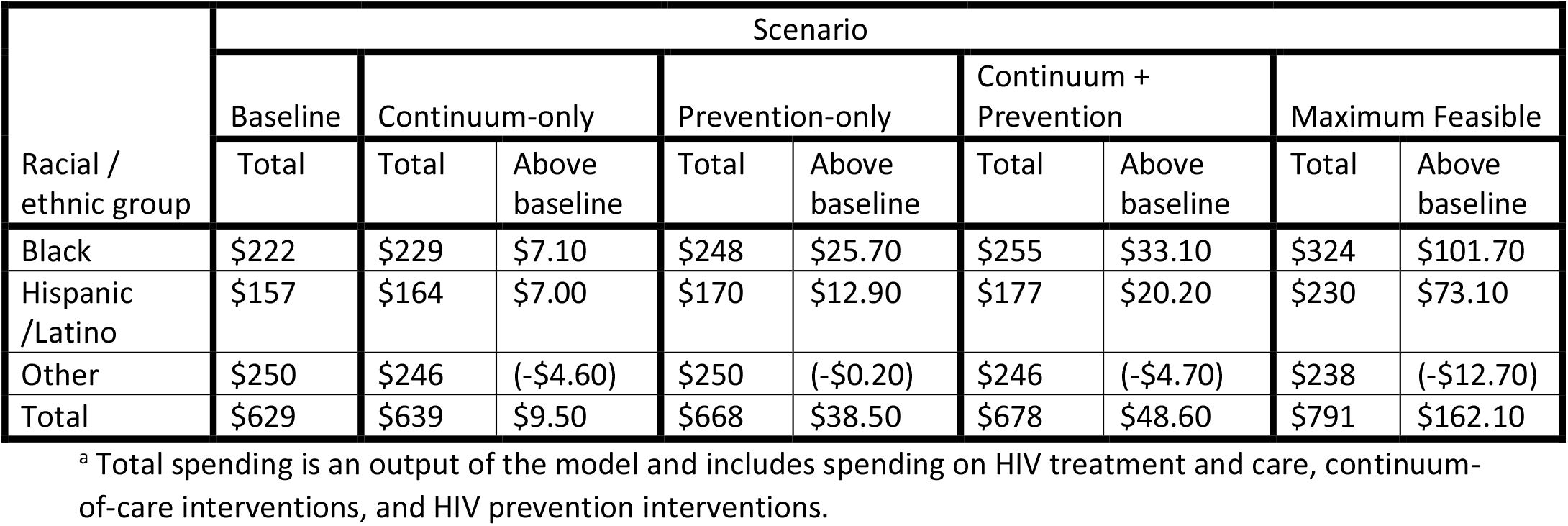
Spending^a^ in 2023-35 for each scenario, by race/ethnicity (billions of 2022 USD)

**Figure 3:**
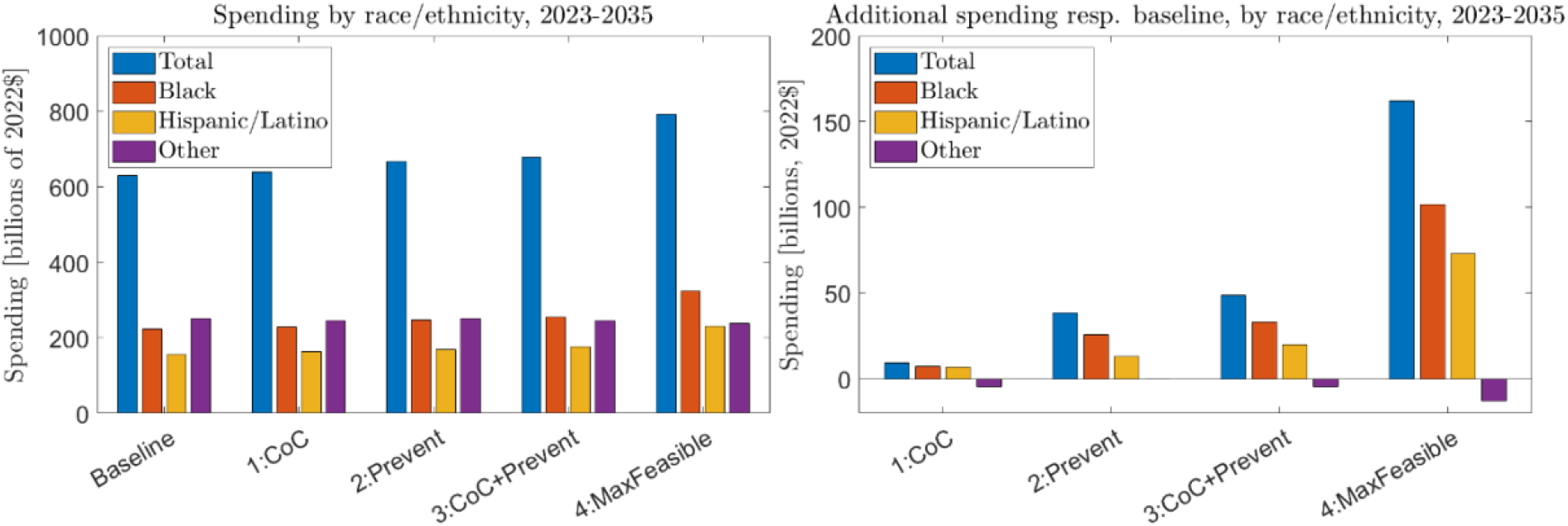
Total spending and spending above baseline in 2023-35 for each scenario, overall and by race/ethnicity

## Discussion

Elimination of racial/ethnic disparities is an important goal of the EHE initiative. We used the HOPE model to identify intervention focus areas that can provide a path toward reducing or eliminating disparities in HIV incidence. Our model predicted that, with no intervention, large disparities in IRRs would persist through 2035, and that while eliminating r/e disparities in the HIV care continuum by 2027 greatly reduced incidence disparities by 2035, it did not eliminate them. We found that increasing the focus on prevention-based interventions, particularly PrEP and SSP, was less effective than increasing the focus on HIV care continuum interventions, and that combining a focus on both prevention-based and care continuum interventions reduced incidence disparities only slightly more than care continuum interventions alone. Eliminating racial/ethnic disparities in HIV incidence required reaching HIV prevention and care continuum levels by 2027 for Black and Hispanic/Latino populations that far exceeded the levels of the Other population. Our simulation model predicted that incidence disparity elimination is only possible if HIV care continuum efforts ensure that Black and Hispanic/Latino populations are able to reach very high rates of awareness, linkage, and VS, coupled with substantial increases in PrEP and SSP coverage. Increased intervention efforts will require additional spending compared to baseline levels; the cost of the complete disparity-elimination scenario was $160 billion higher than the baseline scenario over the 2023–2035 time horizon.

We accounted for the effects of the COVID-19 pandemic on HIV testing, ART use/adherence, PrEP use/adherence, sexual activity levels, and population mortality from 2020-2022 following the approach described in (Viguerie, 2024). A comparison with a hypothetical, COVID-free scenario showed that the effects of the pandemic on HIV disparities were small and transient, essentially disappearing by 2025 scenario (Figure 5.2 in Supplement A).

### Limitations

Our analysis is limited in several important ways. Our national-level model may not properly account for varying levels of racial/ethnic disparities across different jurisdictions. We did not model the White, Asian, Pacific Islander and American Indian populations specifically, and instead combined these populations together in a single “other” category. The American Indian and Pacific Islander PWH populations are too small to inform the necessary model parameters (CDC, 2024a). For Asian Americans, indicators of interest, including HIV incidence/prevalence rates, awareness of status, and viral suppression, were similar to, or better than, those of White Americans, and Asian Americans represent a very small portion of estimated new infections and prevalence (CDC, 2024a; CDC, 2024b); hence, consolidating the groups would have little to no impact on the disparity analysis. Nonetheless, due to the aggregation, our analysis is not capable of making conclusions about any of these groups specifically.

We also did not examine disparities in HIV incidence in other population stratifications besides race/ethnicity, including age, HIV transmission group, and sex assigned at birth. Similarly, we did not study disparities within r/e populations. Our spending estimates are based on unit intervention costs remaining at 2022 levels. As such, future changes in costs of HIV testing, ART and/or PrEP, or other new developments may result in different levels of spending than estimated.

In this analysis, we examined the effects of eliminating racial/ethnic disparities in the two key HIV intervention focus areas – HIV Prevention (specifically the use of PrEP and SSPs only), and the HIV care continuum (awareness of status, linkage to HIV care, and viral suppression), as well as the impact of reaching maximum feasible levels in both areas, on racial/ethnic disparities in HIV incidence. However, this modeling analysis does not address how best to achieve these care continuum and prevention outcomes. There may be several approaches needed to achieve these high outcome levels, which may include individual, community, and structural-level interventions. However, these interventions are not the focus of this modeling analysis. Economic, program, and intervention evaluation studies and systematic reviews focusing on identifying evidence-based interventions can inform program planners on which intervention approaches would work best (HIV Prevention Research Synthesis Project 2024, Higa 2022, Shrestha 2023, Dunlap 2024).

Additionally, this analysis investigated expanding oral PrEP and did not consider newer delivery methods of PrEP, such as long-acting injectable PrEP. Recent biomedical advances, particularly in long-acting PrEP and ART, have much promise in accelerating progress and could be critical tools for reducing or eliminating disparities. Given their current higher costs and low use, however, implementation and scale up may be slow and higher costs may actually be a barrier to rapid and equitable scale up. An important next step in research would be to assess the potential impact on disparities of these long-acting PrEP and ART advances. We also determined PrEP eligibility using 2017 CDC guidelines (CDC 2018) and did not consider any potential effects related to modifying PrEP eligibility. Finally, our analysis focused only on those intervention benefits directly concerning HIV incidence and the HIV continuum of care. Possible ancillary benefits of interventions outside of HIV, such as reductions in other STIs or increased engagement with healthcare providers, were not considered.

### Implications

Our analysis provides policymakers, program planners, and HIV prevention and healthcare providers with several important insights into reducing disparities in HIV incidence and ensuring equitable outcomes across different racial/ethnic populations in the United States. We identify four key takeaways.

First, our findings suggest that racial/ethnic disparities in HIV incidence cannot solely be explained disparities in the HIV care continuum and prevention outcomes. Indeed, we found that, even when eliminating disparities in the HIV care continuum and prevention outcomes, corresponding disparities in HIV incidence, while significantly reduced, nonetheless persisted. This suggests that racial/ethnic disparities in HIV incidence go beyond differential HIV prevention and care efforts and likely reflect other inequities as well.

Second, eliminating racial/ethnic disparities in HIV incidence will require significantly higher levels of HIV prevention and care among Black and Hispanic/Latino populations, as compared to other populations. As discussed, HIV incidence disparities cannot be attributed to disparities in HIV prevention and care efforts alone, and likely reflect a complex, multidimensional interaction of underlying, systemic inequities. Even if disparities in HIV prevention and care outcomes are eliminated, disparities in HIV prevalence will continue to fuel disparities in HIV incidence. To compensate for such structural inequities and disparities in HIV prevalence, levels of HIV prevention and care among Black and Hispanic/Latino populations must be increased far beyond those of other populations to eliminate disparities in HIV incidence. As our spending estimates show, this will require increased investment in HIV prevention efforts, particularly towards Black and Hispanic/Latino populations.

Third, the most effective way to reduce disparities in incidence, according to our study, is to improve HIV care continuum levels among Black and Hispanic/Latino populations. Interventions aimed towards improving awareness of HIV status, linkage to care after diagnosis, and viral suppression among persons living with diagnosed HIV are e of utmost importance. Increasing PrEP use and SSPs will yield additional benefits and are certainly worthwhile. However, to reduce HIV incidence disparities, HIV care continuum interventions should be the primary focus of HIV prevention efforts, with the ultimate endpoint of viral suppression, particularly given that approximately 80% of new infections are linked to PWH who are undiagnosed or who are not virally suppressed (Jacobson 2022, Li Vital Signs, 3-22-2019).

The fourth and final takeaway is closely related to the third. Our spending estimates suggest that, in addition to being the most effective approach for reducing incidence disparities, focusing on the HIV care continuum is also more cost efficient. The *Continuum-only* scenario showed significantly larger reductions in IRR compared to the *Prevention-only* scenario, at less than 25% of the added cost. Further, while the *Continuum + Prevention* scenario required nearly 5 times more additional spending than the *Continuum-only* scenario, the improvements in disparity outcomes were comparatively marginal.

Reducing racial/ethnic disparities in HIV incidence will require a concerted effort, innovative strategies and increased investments to expand the access to and improve the effectiveness of HIV prevention and care services within Black and Hispanic/Latino communities.

## Supporting information

Technical appendix

## Data Availability

All data produced in the present study are available upon reasonable request to the authors.

---

**Supplementary Figure A:**
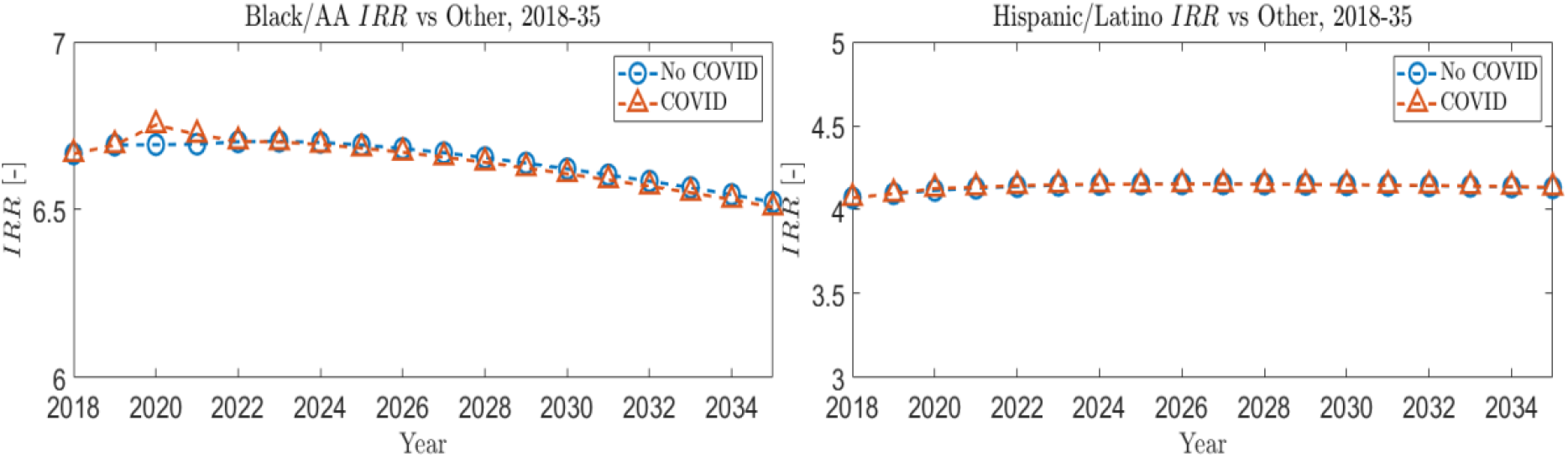
Racial/ethnic disparities in HIV incidence in time as compared to a hypothetical scenario without COVID-19 effects.

## References

1. Alang S, Blackstock O. Health Justice: A Framework for Mitigating the Impacts of HIV and COVID-19 on Disproportionately Affected Communities. Am J Public Health. 2023;113(2):194–201. doi:10.2105/AJPH.2022.307139

2. Bonacci RA, Smith DK, Ojikutu BO. (2021) Toward Greater Pre-exposure Prophylaxis Equity: Increasing Provision and Uptake for Black and Hispanic/Latino Individuals in the U.S. Am J Prev Med. 2021;61(5S1):S60-S72. doi:10.1016/j.amepre.2021.05.027

3. Centers for Disease Control and Prevention. Estimated HIV incidence and prevalence in the United States, 2018–2022. HIV Surveillance Supplemental Report 2024a;29(No. 1). https://stacks.cdc.gov/view/cdc/156513 Published May 2024. Accessed July 18, 2024.

4. Centers for Disease Control and Prevention. Monitoring selected national HIV prevention and care objectives by using HIV surveillance data—United States and 6 territories and freely associated states, 2022. HIV Surveillance Supplemental Report 2024b;29(No. 2). https://stacks.cdc.gov/view/cdc/156511 Published May 2024. Accessed July 18, 2024.

5. Chan SS, Chappel AR, Maddox KEJ, et al. Pre-exposure prophylaxis for preventing acquisition of HIV: A cross-sectional study of patients, prescribers, uptake, and spending in the United States, 2015-2016. PLoS Med. 2020;17(4):e1003072. Published 2020 Apr 10. doi:10.1371/journal.pmed.1003072Dailey A, Gant Z, Hu X, Lyons SJ, Okello A, Johnson AS. A Census Tract-Level Examination of Diagnosed HIV Infection and Social Vulnerability Themes Among Black/African American, Hispanic/Latino, and White Adults, 2019-USA. J Racial Ethn Health Disparities. Published online February 17, 2023. doi:10.1007/s40615-023-01533-5

6. Jenness SM, Maloney KM, Smith DK, et al. (2019) Addressing Gaps in HIV Preexposure Prophylaxis Care to Reduce Racial Disparities in HIV Incidence in the United States. Am J Epidemiol. 2019;188(4):743–752. doi:10.1093/aje/kwy230

7. Handanagic S, et al. (2021) HIV Infection and HIV-Associated Behaviors Among Persons Who Inject Drugs - 23 Metropolitan Statistical Areas, United States, 2018. MMWR Morb Mortal Wkly Rep. 2021;70(42):1459–1465. Published 2021 Oct 22. doi:10.15585/mmwr.mm7042a1

8. Hoover KW, Khalil GM, Cadwell BL, Rose CE, Peters PJ. Benchmarks for HIV Testing: What Is Needed to Achieve Universal Testing Coverage at U.S. Ambulatory Healthcare Facilities. J Acquir Immune Defic Syndr. 2021;86(2):e48–e53. doi:10.1097/QAI.0000000000002553 Kanny D, Jeffries WL 4th, Chapin-Bardales J, et al. Racial/Ethnic Disparities in HIV Preexposure Prophylaxis Among Men Who Have Sex with Men - 23 Urban Areas, 2017. MMWR Morb Mortal Wkly Rep. 2019;68(37):801–806. Published 2019 Sep 20. doi:10.15585/mmwr.mm6837a2

9. Khurana, N, Yaylali, E, Farnham, P G, Hicks, K. A., Allaire, B T, Jacobson, E, & Sansom, S L (2018). Impact of improved HIV care and treatment on PrEP effectiveness in the United States, 2016– 2020. Journal of Acquired Immune Deficiency Syndrome, 78(4), 399–405.

10. Lyons SJ, Dailey AF, Yu C, Johnson AS. Care Outcomes Among Black or African American Persons with Diagnosed HIV in Rural, Urban, and Metropolitan Statistical Areas - 42 U.S. Jurisdictions, 2018. MMWR Morb Mortal Wkly Rep. 2021;70(7):229–235. Published 2021 Feb 19. doi:10.15585/mmwr.mm7007a1

11. McCree DH, Chesson HW, Eppink ST, Beer L, Henny KD. Changes in Racial and Ethnic Disparities in Estimated Diagnosis Rates of Heterosexually Acquired HIV Infection Among Heterosexual Males in the United States, 2014-2018. J Acquir Immune Defic Syndr. 2020;85(5):588–592. doi:10.1097/QAI.0000000000002495

12. Nosyk B, Zang X, Krebs E, et al. (2020a) Ending the HIV epidemic in the USA: an economic modelling study in six cities. Lancet HIV. 2020;7(7):e491–e503. doi:10.1016/S2352-3018(20)30033-3

13. Nosyk B, Krebs E, Zang X, et al. (2020b) “Ending the Epidemic” Will Not Happen Without Addressing Racial/Ethnic Disparities in the United States Human Immunodeficiency Virus Epidemic. Clin Infect Dis. 2020;71(11):2968–2971. doi:10.1093/cid/ciaa566

14. Pyra M, Brewer R, Rusie L, Kline J, Willis I, Schneider J. (2021) Long-term HIV Pre-exposure Prophylaxis Trajectories Among Racial & Ethnic Minority Patients: Short, Declining, & Sustained Adherence [published online ahead of print, 2021 Oct 9]. J Acquir Immune Defic Syndr. 2021;10.1097. doi:10.1097/QAI.0000000000002833

15. Quan AML, Mah C, Krebs E, et al. Improving health equity and ending the HIV epidemic in the USA: a distributional cost-effectiveness analysis in six cities. Lancet HIV. 2021;8(9):e581–e590. doi:10.1016/S2352-3018(21)00147-8

16. Raiford JL, Yuan X, Carree T, Beer L. Understanding Disparities in Antiretroviral Therapy Adherence and Sustained Viral Suppression Among Black, Hispanic/Latina, and White Women in the United States - Medical Monitoring Project, United States, 2015-2019. J Acquir Immune Defic Syndr. 2023;93(5):413–421. doi:10.1097/QAI.0000000000003214

17. Sullivan PS, Knox J, Jones J, et al. Understanding disparities in viral suppression among Black MSM living with HIV in Atlanta Georgia. J Int AIDS Soc. 2021;24(4):e25689. doi:10.1002/jia2.25689.

18. U.S. Health and Human Services. (2019, August 18). America’s HIV Epidemic Analysis Dashboard (AHEAD). Retrieved from https://ahead.hiv.gov/. Accessed December 8, 2022.

19. DiNenno EA, Delaney KP, Pitasi MA, et al. HIV Testing Before and During the COVID-19 Pandemic — United States, 2019–2020. MMWR Morb Mortal Wkly Rep 2022; 71: 820–4.

20. Huang Y-LA, Zhu W, Wiener J, Kourtis AP, Hall HI, Hoover KW. Impact of Coronavirus Disease 2019 (COVID-19) on Human Immunodeficiency Virus (HIV) Pre-exposure Prophylaxis Prescriptions in the United States—A Time-Series Analysis. Clinical Infectious Diseases 2022; published online Jan 18. DOI:10.1093/cid/ciac038.

21. Viguerie A, Song R, Johnson AS, Lyles CM, Hernandez A, Farnham PG. Isolating the Effect of COVID-19-Related Disruptions on HIV Diagnoses in the United States in 2020. JAIDS Journal of Acquired Immune Deficiency Syndromes 2023a; 92: 293–9.

22. R. Dana et al., “Engaging Black or African American and Hispanic or Latino Men Who Have Sex With Men for HIV Testing and Prevention Services Through Technology: Protocol for the iSTAMP Comparative Effectiveness Trial,” JMIR Res Protoc, vol. 12, p. e43414, Jan. 2023, doi: 10.2196/43414.

23. Zhu W, Huang YA, Wiener J, et al. Impact of the coronavirus disease 2019 pandemic on prescriptions for antiretroviral drugs for HIV treatment in the United States, 2019–2021. AIDS 2022; 36: 1697–705.

24. Viguerie A, Jacobson E, Hicks KA, et al., Assessing the impact of COVID-19 on HIV Outcomes in the United States: A modeling study. Sexually Transmitted Diseases. 51(4):299–304, 2023b

25. Jacobson E U, Hicks K A, Carrico J, Purcell, D W, Green T A, Mermin J H, & Farnham P G. (2022). Optimizing HIV prevention efforts to achieve EHE incidence targets. Journal of Acquired Immune Deficiency Syndrome, 89(4), 374–380.

26. Chen Y H, Farnham P G, Hicks K A, & Sansom S L (2022). Estimating the HIV effective reproduction number in the United States and evaluating HIV elimination strategies. Journal of Public Health Management Practice, 28(2).

27. Sansom S L, Hicks K A, Carrico J, Jacobson E U, Shrestha R K, Green T A, & Purcell D W. (2021). Optimal allocation of societal HIV prevention resources to reduce HIV incidence in the United States. American Journal of Public Health, 111(1), 150–158.

28. Viguerie A, Song R, Bosh K, Lyles CM, and Farnham, P.G (2024). Mortality among Persons with HIV in the United States during the COVID-19 pandemic: a population-level analysis. J Acquir Immune Defic Syndr. 95(2), 126–132.

29. HIV Prevention Research Synthesis Project. Compendium of Evidence-Based Interventions and Best Practices for HIV Prevention. Centers for Disease Control and Prevention. https://www.cdc.gov/hiv/research/interventionresearch/compendium/index.html. Updated 16 May 2024. Accessed 7 June 2024.

30. Higa D H, Crepaz N, Mullins M M, Adegbite-Johnson A, Gunn J K L, Denard C, Mizuno Y, & the Prevention Research Synthesis Project. (2022). Strategies to improve HIV care outcomes for people with HIV who are out of care. AIDS, 36(6), 853–862. doi:10.1097/QAD.0000000000003172.

31. Shrestha RK, Fanfair RN, Randall L, Lucas C, Nichols L, Camp N, Brady K, Jenkins H, Altice F, DeMaria A, Villanueva M, Weidle PJ. Costs and cost-effectiveness of a collaborative data-to-care intervention for HIV treatment and care in the United States. J Int AIDS Society 2023; 26: e26040

32. Dunlap LJ, Orme S, Zarkin GA, Holtgrave DR, Maulsby C, Rodewald AM, Holtyn AF, Silverman K. Cost and Cost-Effectiveness of Incentives for Viral Suppression in People Living with HIV. AIDS Behav. 2022 Mar;26(3):795–804. doi: 10.1007/s10461-021-03439-x. Epub 2021 Aug 26. Erratum in: AIDS Behav. 2024 Feb 10;: PMID: 34436714.

33. Centers for Disease Control and Prevention. Preexposure prophylaxis for the prevention of HIV infection in the United States—2017 Update: a clinical practice guideline, 2018.

