## Supplementary material for "A Modeling Analysis on Eliminating Racial/Ethnic Disparities in HIV Incidence in the United States": Technical appendix

August 2024

Technical Report 16 for HOPE Model version 9.10

Most of the material in this technical report has been previously published (O’Leary et al., 2017; Jacobson et al., 2018; Khurana et al., 2018; Chen et al., 2021; Jacobson et al., 2022). However, the model is updated with each new version.

_________________________________
RTI International is a registered trademark and a trade name of Research Triangle Institute.

Contents

Section	Page

1.	Introduction	1-1

2.	Model Overview and Purpose	2-1

3.	Model Population and Compartments	3-1

4.	Initial Population	4-1

5.	Movement into and out of the Model, between Subpopulations, between Compartments (Except Due to Infection), and within Compartments	5-1

5.1	Transitions into and out of the model	5-1

5.2	Transitions between Subpopulations (Aging Only)	5-13

5.3	Transitions between Compartments Due to Disease Progression	5-13

5.3.1	Role of CD4 Count in Disease Progression and Transmission	5-14

5.4	Transitions between Compartments Due to Progression along the Care Continuum	5-14

5.4.1	Methods for Calculating Progression along the Care Continuum	5-15

5.4.2	Diagnosis Rates	5-17

5.5	Transitions between Compartments Due to PrEP Participation	5-18

6.	Force of Infection	6-1

6.1	Sexual and Needle-Sharing Partnerships	6-1

6.2	Per-Partnership Transmission Risk	6-4

6.2.1	Per-Sex-Act Sexual and Needle Transmission Probabilities	6-11

6.2.2	Number of Sex Acts and Needles Shared per Partner	6-12

6.2.3	Calculation of Per-Partnership Transmission Risk	6-12

6.3	Calculation of Force of Infection for Individuals Not on PrEP and Not Participating in SSP	6-14

6.4	Calculation of Force of Infection for Individuals on PrEP	6-16

6.5	Calculation of Force of Infection for Individuals in SSP	6-18

7.	Differential Equations that Define the Model	7-1

7.1	Number of People without HIV	7-2

7.2	Individuals with Acute HIV Infection	7-3

7.3	Individuals with Chronic HIV Infection and CD4 ≥ 200	7-4

7.4	Individuals with Chronic HIV Infection and CD4<200	7-6

7.5	Absorbing States	7-7

8.	Analysis Scenarios	8-7

8.1	Model Inputs For Disparities Analysis	8-7

9.	Calculation of Model Outcomes	9-1

9.1	Health Outcomes	9-1

9.2	Economic Outcomes	9-1

9.2.1	Testing Cost Inputs	9-1

9.2.2	Calculation of Testing and Notification Costs	9-3

9.2.3	Calculation of Transitions Between HIV Continuum-of-Care Stages and Maintaining Viral Load Suppression	9-4

9.2.4	Calculation of HIV Treatment and Care Costs	9-5

9.2.5	Calculation of SSP Costs	9-7

9.2.6	Calculation of PrEP Costs	9-7

10.	Model Calibration and Validation	10-8

10.1	Model Calibration to Published Data	10-8

10.1.1	Establish Calibration Outcome Targets	10-8

10.1.2	Establish Inputs to Vary in Calibration	10-8

10.1.3	Identification of Base and Alternative Input Sets	10-17

10.2	Internal and External Validation of the Model	10-20

10.2.1	Strengths and Limitations of Calibration Set LB20230224_2	10-20

11.	Model Sensitivity and Uncertainty Analyses	11-22

11.1	Uncertainty Analysis	11-22

References	R-1

Appendix A: Definitions	A-1

Figures

Number	Page

Figure 3.1	Model Flow Diagram between Compartments due to HIV Infection, Progression along the Care Continuum, Progression of HIV, and Death	3-4

Figure 5.1	Inputs, Flows, and States Reflecting Oral and Injectable PrEP Use in HOPE	5-20

Tables

Number	Page

Table 3.1.	Population Stratification Criteria and Categories Applied in Each	3-1

Table 4.1.	Percentage of Population with Multiple HIV Transmission Risk Factors, Distribution of PWID by Race / Ethnicity, Relative HIV Prevalence for People with Multiple vs. Fewer HIV Transmission Risk Factors, and Population Sizes by Key Stratifications in Initial Population (2010)	4-2

Table 4.2.	HIV Prevalence in Initial Population (2010)	4-4

Table 4.3.	Percentage Circumcised, and Percentage of Each Transmission Group Sexually Active in Initial Population (2010)	4-7

Table 4.4.	Distribution of Initial HIV-Infected Population across Continuum-of-Care Stages by Race/Ethnicity for All Subpopulations besides MSM ages 13-34 (2010)	4-7

Table 4.5.	Distribution of Initial HIV-Infected Population across Continuum-of-Care Stages by Race/Ethnicity for MSM ages 13-34 (2010)	4-9

Table 4.6.	Distribution of Initial HIV-Infected Population across HIV Stages, by Continuum-of-Care Stage (2010)	4-10

Table 5.1.	Rate of Aging into Population and Inputs that Determine All HIV Progression and Death Rates	5-3

Table 5.2.	Inputs for Calculating Rates of Undiagnosed HIV-Infected and People without HIV Getting Tested in Time Periods 1, 2-4, and 5^a^	5-7

Table 5.3.	Testing Performance Parameters	5-8

Table 5.4.	Annual Probability of Initiating ART in Time Periods 1, 2-4, and 5^a^	5-9

Table 5.5.	Other Continuum-of-Care Probabilities	5-10

Table 5.6	Inputs defining COVID-19 Effects on Continuum-of-Care Progression^a^	5-16

Table 5.9	Inputs defining PrEP participation in HOPE	5-21

Table 6.1.	Distribution of Sexual Partners by Sex and Transmission Group	6-2

Table 6.2.	Distribution of Sexual Partners by Race/Ethnicity, HET and PWID	6-2

Table 6.3.	Distribution of Sexual Partners by Race/Ethnicity, MSM	6-2

Table 6.4.	Distribution of Sexual Partners by Age, HET and PWID	6-3

Table 6.5.	Distribution of Sexual Partners by Age, MSM	6-3

Table 6.5.	Distribution of Sexual Partners by Number of HIV Transmission Risk Factors, by Transmission Group	6-3

Table 6.6.	Distribution of Needle-Sharing Partners by Sex (PWID Only)	6-4

Table 6.7.	Distribution of Needle-Sharing Partners by Race/Ethnicity (PWID Only)	6-4

Table 6.8.	Distribution of Needle-Sharing Partners by Age Group (PWID Only)	6-4

Table 6.9.	Per-Act HIV Transmission Risk and Reductions in Risk Due to Circumcision, Viral Load Suppression, Condom Use, and Awareness of HIV Infection	6-5

Table 6.10.	Sexual Partners and Sex Acts	6-6

Table 6.11.	Other Risk Behaviors and Inputs that Determine Transmission Risk	6-10

Table 6.12.	Five Sources of Infection that Contribute to Overall Force of Infection for Each Subpopulation	6-14

Table 6.13.	Inputs for Defining Reduction in Infection Risk if on PrEP	6-17

Table 6.14.	Participation and Infection Risk Reduction for SSP	6-19

Table 7.1.	Model Compartments	7-1

Table 8.1	Relevant model inputs for the five different scenarios	8-7

Table 8.2	Key health equity indicators in 2027 for the five different scenarios	8-9

Table 9.3.	Testing Cost Inputs	9-2

Table 9.4.	Costs of Transitions along the HIV Continuum-of-Care and Maintenance of VLS	9-5

Table 9.5.	Annual HIV Treatment and Care Costs	9-6

Table 9.6.	Annual PrEP Costs	9-7

Table 9.1.	Bounds and Final Values of Continuum-of-Care Parameters Varied in Calibration^a^	10-9

Table 9.2.	Bounds and Final Values of Inputs Defining Behaviors and Infectivity Varied in Calibration^a^	10-11

Table 9.3.	Values Generated by the Model using the Base Analysis Set vs. Target Values and Bounds Considered for Outcomes Targeted in Calibration	10-18

Table A.1.	Definitions of Symbols Applied in This Document	2

Table A.2.	Definitions of Indices Applied in This Document	6

Disclaimer: Previous versions of this document were published as supplementary material to the following papers (in order by publication date):

O’Leary A, DiNenno E, Honeycutt A, Allaire B, Neuwahl S, Hicks K, Sansom S. Contribution of anal sex to HIV prevalence among heterosexuals: a modeling analysis. AIDS and Behavior. 2017 Oct;21:2895-903.

Jacobson EU, Hicks KA, Tucker EL, Farnham PG, Sansom SL. Effects of reaching national goals on HIV incidence, by race and ethnicity, in the United States. Journal of Public Health Management and Practice. 2018 Jul 1;24(4):E1-8.

Khurana N, Yaylali E, Farnham PG, Hicks KA, Allaire BT, Jacobson E, Sansom SL. Impact of improved HIV care and treatment on PrEP effectiveness in the United States, 2016–2020. JAIDS Journal of Acquired Immune Deficiency Syndromes. 2018 Aug 1;78(4):399-405.

Chen YH, Farnham PG, Hicks KA, Sansom SL. Estimating the HIV Effective Reproduction Number in the United States and Evaluating HIV Elimination Strategies. Journal of Public Health Management and Practice. 2021 Aug 2.

Jacobson EU, Hicks KA, Carrico J, Purcell DW, Green TA, Mermin JH, Farnham PG. Optimizing HIV prevention efforts to achieve EHE incidence targets. JAIDS Journal of Acquired Immune Deficiency Syndromes. 2022 Apr 1;89(4):374-80.

### 1. Introduction

This document presents the technical details of Version 9.10 of the HIV Optimization and Prevention Economics (HOPE) Model, a differential equations model representing the U.S. HIV epidemic developed by the Centers for Disease Control and Prevention’s (CDC’s) Division of HIV/AIDS Prevention and RTI International. Section 2 gives a brief overview of the model and its purpose. Section 3 explains the core structure of the model, including the modeled population, the compartments used in the model, and the notation applied in this document. Section 4 describes the initial model population. Section 5 discusses the transitions that result in the flow of the population between compartments (except the flow due to infection) and the flow into and out of the model. Section 6 describes how the force of infection is calculated. Section 7 explains the differential equations that are applied in the model. Section 8 describes the calculation of model outcomes. Section 9 discusses the model’s calibration and validation. Section 10 presents the methods of sensitivity and uncertainty analyses. Appendix A includes tables defining the symbols and indices applied in this document.

As noted in the disclaimer at the beginning of this document, several versions of this report have previously been published alongside papers that use the HOPE Model. Each version prior to this has been adapted for the specific analysis presented in that publication. Besides reflecting the assumptions and structure of the model version used (9.10), the following is a list of how this version of this document has been adapted to specifically represent this analysis:

- Sections 5, 6, and 7: All input tables reflect the set of values used for calibrated inputs for this analysis (the set with identifier “LB20230224_2”).
- Section 8: This section contains scenarios modeled for this analysis.
- Section 10: The calibration methods represent those applied in generating the set of values used to populate all calibrated inputs in this analysis (set LB20230224_2). The ranges considered for all calibrated inputs and the point estimates and acceptable ranges of output values for all targeted outcomes are also specific to the analysis, as are the final values of the inputs and resulting values of the targeted outcomes when those input values were applied in the HOPE Model.
- Section 11: The sensitivity and uncertainty analyses methods and results are specific to this analysis.

### 2. Model Overview and Purpose

The objectives of the HOPE model are to estimate the impact on health outcomes related to HIV in the U.S. population resulting from changes in the distribution of people with HIV along the HIV care continuum and to better understand the impact of different risk behaviors on the prevalence of HIV in the United States. The model uses discretized difference equations that are implemented in MATLAB software (Mathworks, Natick, Massachusetts). The equations represent the dynamics of the HIV-uninfected and HIV-infected populations, including their interaction and progression through various clinical stages of HIV as well as the care continuum. We consider progression through the care continuum to include HIV diagnosis, linkage to care, ART initiation, and achievement of viral suppression. Transmission risks from sex acts (vaginal and anal) and shared needles were considered. The model can be used to examine HIV prevention interventions aimed at people with and without HIV. The HOPE model also includes the flexibility to consider the effects of the SARS-Cov-2 (COVID-19) pandemic on mortality rates (details provided in Section 5.1), rates of progression along the HIV care continuum (details in Section 5.4.1), PrEP usage (details in Section 5.5), and sexual behaviors (details in Section 6.2).

Our model was originally based on a model published by Sorensen and colleagues (2012) that included gay, bisexual, and other men who have sex with men (collectively referred to as MSM) in New York City; we expanded that model to include the national epidemic data for multiple transmission groups (MSM, people who inject drugs [PWID], and heterosexuals [HETs]). These groups were further categorized by sex (male, female), race/ethnicity (Black, Hispanic/Latino, white/other), age group (13–17, 18–24, 25–34, 35–44, 45–54, 55–64, and 65+ years), and number of HIV transmission risk factors (fewer / multiple). We also adjusted the model’s care-continuum stages to more precisely represent the effects of antiretroviral therapy (ART) and being virally suppressed (VLS). The model is dynamic in that the risk of people without HIV acquiring HIV at any given time is a function of the current size, disease stages, and HIV care-continuum stages of the portion of their pool of sexual and needle-sharing partners with HIV. As a result, changes that affect people with HIV (PWH) in the model affect the spread of the disease to people without HIV in the model.

Five time periods are observed. The first, 2010 to 2019, is aimed at replicating historical data and trends. The second through fourth periods offer optional functionality to reflect disruptions due to the COVID-19 pandemic in years 2020 to 2022. The fifth period is for forecasting future trends under various assumptions to represent hypothetical scenarios. For example, the fifth period can assume the same trends as one of the previous periods or changes in trends that may result from policy or funding changes. The years defining each period are user-modifiable, but the second to fifth periods are optional and, if used, must begin after the first period.

### 3. Model Population and Compartments

The population observed in the model is stratified by age group *j* (*j* = 1, …, 7), number of HIV transmission risk factors *k* (*k* = 1, 2), transmission group *l* (*l* = 1, 2, 3), sex *m* (*m* = 1, 2), circumcision status *n* (*n* = 1, 2), and race/ethnicity *o* (*o* = 1, 2, 3). Table 3.1 lists the categories applied in the model for each of these stratification criteria. The model assumes that PWID have multiple HIV transmission risk factors, MSM are all male, and only males are stratified by circumcision status; therefore, not all combinations of these stratifications (i.e., 504 = 7x2x3x2x2x3) are represented. Males are stratified by seven groups based on age, five groups combining transmission group and number of HIV transmission risk factors (MSM with multiple HIV transmission risk factors, MSM with fewer HIV transmission risk factors, HET with multiple HIV transmission risk factors, HET with fewer HIV transmission risk factors, PWID), two groups based on circumcision status, and three groups based on race/ethnicity, resulting in 210 (7x5x2x3) male subpopulations. Females are stratified by seven groups based on age, three groups combining transmission and number of HIV transmission risk factors (HETs with multiple HIV transmission risk factors, HETs with fewer HIV transmission risk factors, and PWID), and three groups based on race/ethnicity, resulting in 63 (7x3x3) female subpopulations. Accounting for both males and females, there are 273 (210+63) subpopulations *p* (*p* =1, …, 273).

Table 3.1.	Population Stratification Criteria and Categories Applied in Each

| Stratification Criterion (Represented by) | Categories (Represented by) |
| --- | --- |
| Age group (*j*) (years) | 13–17 (1) |
|  | 18–24 (2) |
|  | 25–34 (3) |
|  | 35–44 (4) |
|  | 45–54 (5) |
|  | 55–64 (6) |
|  | 65+ (7) |
| Number of HIV transmission risk factors (*k*)^a^ | Fewer (1) |
|  | Multiple (2) |
| Transmission group (*l*) | HET (1) |
|  | MSM (2)^b^ |
|  | PWID (3) |
| Sex (*m*) | Male (1) |
|  | Female (2) |
| Circumcision status (*n*) | Uncircumcised (1) |
|  | Circumcised (2) |
| Race/ethnicity (*o*) | Black (1) |
|  | Hispanic/Latino (2) |
|  | White/other (3) |

Note: HET = heterosexual; MSM = men who have sex with men; PWID = people who inject drugs

^a^ Populations with “multiple” versus “fewer” risk factors were defined by criteria that varied between transmission risk groups. Risk factors for heterosexuals (HETs) include the following: 2+ sex partners in the last 12 months, and either (a) had a bacterial STI, defined as gonorrhea, or (b) had mostly condomless sexual contacts in the last 12 months (more than half the time). Risk factors for men who have sex with men (MSM) include the following: 2+ anal sex partners in, the last 12 months and either (a) had a bacterial STI, defined as gonorrhea or chlamydia, or (b) had any condomless sexual contacts within the past 12 months. We assumed all persons who inject drugs (PWID) had multiple HIV transmission risk factors. These criteria were defined to be consistent with CDC clinical practice guidelines (CDC, 2017b). These categories were reflected by “high and low risk levels” in previous versions of the HOPE Model.

^b^ The MSM population is meant to capture all men who have sex with men, not just those who self-identify as MSM.

The model’s 30 compartments (defined by *c*) included 28 main compartments for individuals actively moving through the model and 2 compartments for individuals who were no longer actively followed in the model due to death. The 28 former compartments were defined by disease stage (*h*) and continuum-of-care stage (*r*) for PWH; and, for people without HIV, PrEP status.

Figure 3.1 displays how the model’s compartments are applied in the model; each compartment is labeled in that figure with its corresponding number *c*. They are defined by HIV stage, HIV continuum-of-care status, and PrEP status. Individuals enter the population people without HIV and not on PrEP (*c* = 1). They may die or age out of the population from any of the main compartments.

Six disease stages (*h* = 0, …, 5) were defined by the presence of HIV infection and, for PWH, HIV progression:

- *h* = 0: People without HIV
- *h* = 1: PWH with acute HIV infection (“Acute” hereafter)
- *h* = 2: PWH with CD4 count greater than 500 cells/mm^3^ (but not acute) (“CD4>500” hereafter)
- *h* = 3: PWH with CD4 count between 350 cells/mm^3^ and 500 cells/mm^3^ (“CD4 350-500” hereafter)
- *h* = 4: PWH with CD4 count between 200 cells/mm^3^ and 350 cells/mm^3^ (“CD4 200-350” hereafter)
- *h* = 5: PWH with CD4 count less than 200 cells/mm^3^ (“CD4<200” hereafter)

Five continuum-of-care stages (*r* = 1, …, 5) were defined by HIV status, awareness of infection, linkage to HIV care, status based on effects of treatment with ART, and VLS status:

- *r* = 1: People without HIV and PWH unaware of infection (“Unaware” hereafter)
- *r* = 2: PWH aware of infection, but not linked to HIV care (“Aware” hereafter)
- *r* = 3: PWH linked to HIV care, but not on ART (“Linked to HIV care” or “LTC” hereafter)
- *r* = 4: PWH on ART, but not VLS (“ART-not-VLS” or “ANV” hereafter)
- *r* = 5: PWH with VLS (in care and on ART) (“VLS” hereafter)

For the acute disease stage (*h* = 1), compartments were defined by only the first three continuum-of-care stages: unaware, aware, and LTC (*r* = 1, 2, 3), i.e., we assume that individuals with acute HIV are not on ART. For each of the four disease stages for individuals with chronic HIV (*h* = 2, 3, 4, 5), compartments were defined by all continuum-of-care stages for PWH (*r* = 1, 2, 3, 4, 5). People without HIV (*h* = 0) also were categorized by PrEP status, and among those on PrEP, by type of PrEP delivery (oral or injectable) and adherence level (high or low) (*c* = 1 to 5).

The population represented in the model at any given time was distributed among the 28 main compartments. Individuals in the population were defined by the compartment they occupied, as well as by other demographic and behavioral factors. We let $X_{p}^{c}(t)$ equal the number of individuals in subpopulation *p* in compartment *c* at time *t*. Appendix Table A.1 lists and defines all symbols used in this document.

Figure 3.1	Model Flow Diagram between Compartments due to HIV Infection, Progression along the Care Continuum, Progression of HIV, and Death

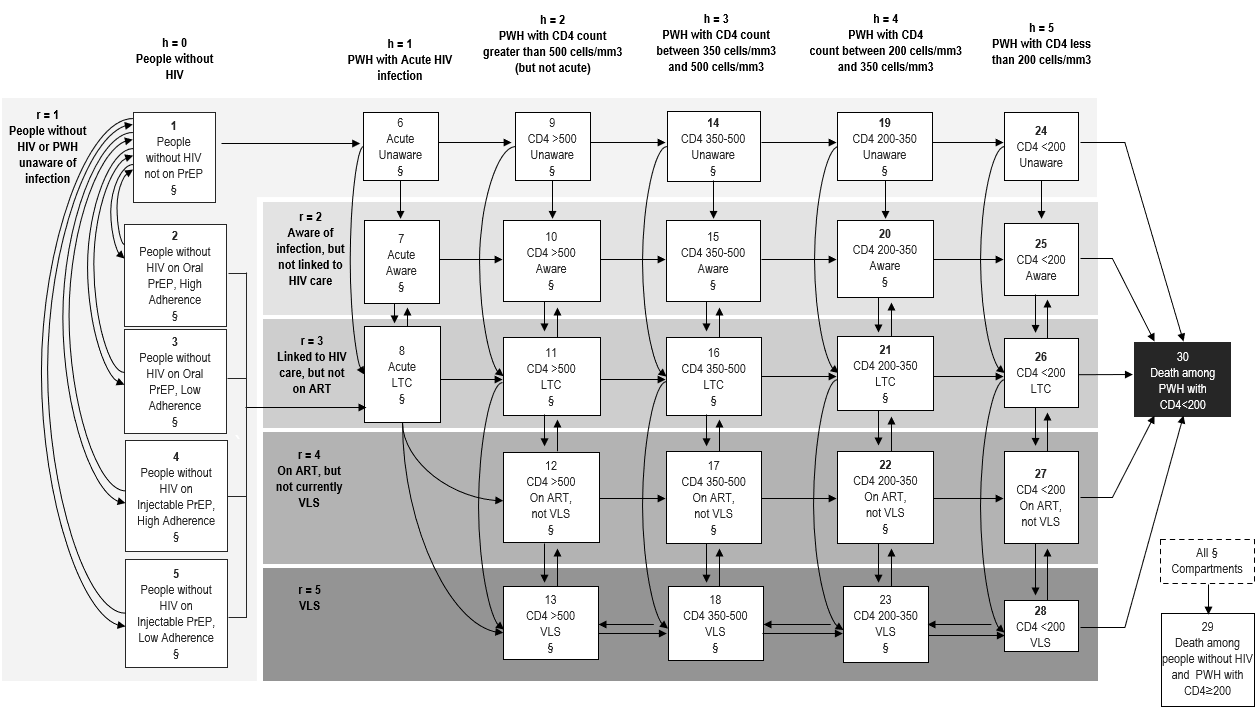

Note: ART = antiretroviral therapy; LTC = linked to HIV care; PrEP = pre-exposure prophylaxis; PWH = people with HIV; VLS = viral load suppression.

### 4. Initial Population

The model was initiated so that the observed population was distributed among the model’s 28 main compartments and was characterized to match the total population and PWH in the United States in 2010. The size of the population was set to capture sexually active individuals in the U.S. population; it is flexibly programmed to either include or exclude ages 13 to 17 based on user settings. The size of and distribution among demographic subpopulations were determined by a set of parameters listed in Tables 4.1 through 4.6.

Table 4.1.	Percentage of Population with Multiple HIV Transmission Risk Factors, Distribution of PWID by Race / Ethnicity, Relative HIV Prevalence for People with Multiple vs. Fewer HIV Transmission Risk Factors, and Population Sizes by Key Stratifications in Initial Population (2010)

| Parameter | Female | | | | | | Male | | | | | | | Total | Source |
| --- | --- | --- | --- | --- | --- | --- | --- | --- | --- | --- | --- | --- | --- | --- | --- |
|  | Black | | Hispanic | | White/other | | Black | Hispanic | | | | White/other | |  |  |
| Percentages of HET, PWID, and MSM with multiple HIV transmission risk factors | | | | | | | | | |  | | | | | |
| HET | 10.6% | 6.6% | | 7.7% | | | 10.6% | | 6.6% | | | | 7.7% |  | RTI unpublished analysis using NHANES 2009-2016 data.^a^ |
| PWID | -----------------------------------------100.0%------------------------------------------------- | | | | | | | | | | | | |  | Assumed^b^ |
| MSM | ------------------------------------------37.3%------------------------------------------------- | | | | | | | | | | | | |  | RTI unpublished analysis using NHANES 2009-2016 data.^c^ |
| Percentages of PWID population that is each race/ethnicity, by sex | | | | | | | | | | |  | | | | |
|  | 33.80% | | 22.50% | | 43.7% | 33.80% | | 22.50% | | | | 43.7% | |  | CDC unpublished analysis of NHBS 2015 IDU Cycle |
| Initial population size, total and by transmission group and number of HIV transmission risk factors^d^ | | | | | | | | | | | | | | | |
| U.S. population aged 13+ | 16,161,713 | | 18,661,868 | | 96,245,883 | 14,277,026 | | 19,165,505 | | | | 91,290,325 | | 255,802,320 | U.S. Census Bureau (2010) |
| HET (Sexually Active), by number of HIV transmission risk factors | | | | | | | | | | | | | | |  |
| Multiple | 1,502,598 | | 1,086,199 | | 6,533,202 | 1,244,626 | | 1,054,391 | | | | 5,960,552 | | 17,381,569 | Calculated^e^ |
| Fewer | 12,727,671 | | 15,410,849 | | 78,834,210 | 10,542,536 | | 14,959,567 | | | | 71,924,211 | | 204,399,043 | Calculated^e^ |
| PWID |  | |  | |  |  | |  | | | |  | |  |  |
|  | 150,499 | | 100,184 | | 194,580 | 388,925 | | 258,900 | | | | 502,841 | | 1,595,929 | Calculated^f^ |
| MSM |  | |  | |  |  | |  | | | |  | |  |  |
| Overall |  | |  | |  | 625,754 | | 888,471 | | | | 3,155,299 | | 4,669,524 | Rosenberg et al. (2018)^g^ |
| Multiple |  | |  | |  | 233,406 | | 331,400 | | | | 1,176,927 | | 1,741,732 | Calculated^c^ |
| Fewer |  | |  | |  | 392,348 | | 557,071 | | | | 1,978,372 | | 2,927,792 | Calculated^c^ |
| Relative HIV prevalence for people with multiple vs. fewer HIV transmission risk factors | | | | | | | | | | | | | | | |
| MSM | ---------------------------------------------4.00------------------------------------------------ | | | | | | | | | | | | |  | Assumption based on NHBS 2017 (CDC, 2019b).^h^ |
| HET |  | | | | | | | | | | | | |  | Assumption based on Woodring (2015) and SME^i^ |
| Black | --------------------------------------------20.00------------------------------------------------ | | | | | | | | | | | | |  |  |
| Hispanic | --------------------------------------------10.00------------------------------------------------ | | | | | | | | | | | | |  |  |
| Other | ---------------------------------------------5.00------------------------------------------------- | | | | | | | | | | | | |  |  |

Note: CDC = Centers for Disease Control and Prevention; HET = heterosexual; HIV = human immunodeficiency virus; MSM = men who have sex with men; N/A = not applicable; NHANES = National Health and Nutrition Examination Survey; NHBS = National HIV Behavioral Surveillance; PWID = people who inject drugs

^a^ Determined by an unpublished RTI analysis of the percentage of HETS in NHANES waves 2009-2016 who met the following CDC 2017 PrEP eligibility criteria: (1) had not already been defined as MSM or PWID, and (2) had 2+ sex partners in the last 12 months, and either had a bacterial STI, defined as gonorrhea, or had mostly condomless (more than half the time) sexual contacts in the last 12 months (CDC, 2018d).

^b^ All PWID assumed to have multiple HIV transmission risk factors.

^c^ Determined by an unpublished RTI analysis of the percentage of MSM (defined as men who had any male sexual partners in the past 12 months) in NHANES waves 2009-2016 who met the following CDC 2017 PrEP eligibility criteria for MSM: (1) had 2+ anal sex partners in the last 12 months and (2) either had a bacterial sexually-transmitted infection with gonorrhea or chlamydia OR had any condomless sexual contacts within the past 12 months (CDC, 2018d).

^d^ Initial total U.S. population size inputs used in HOPE are stratified by age, sex, and race/ethnicity. For simplicity, the values reported here are totals by sex and race/ethnicity across all ages.

^e^ The number of sexually active HETs overall was determined by subtracting the number of MSM and PWID from the total US population aged 13+, then multiplying the resulting number by the percentage of that population that is sexually active (88.9%). The percentage of the population sexually active is based on percentages of individuals reporting sex with opposite partner in their lifetimes, as estimated from Lansky et al. (2015). These numbers were then multiplied by the percentage of sexually active HETs with multiple HIV transmission risk factors to get counts of sexually active HETs by number of HIV transmission risk factors.

^f^ Tempalski et al. (2013), identified 1,595,929 total PWID in the US who had injected in their lifetime. An unpublished analysis of the 2015 NHBS IDU Cycle showed that PWID were distributed by gender as 72.1% males and 27.9% females. This distribution was applied to the overall PWID estimate from Tempalski to determine the size of the adult population that is PWID for each gender.

^g^ Rosenberg et al. (2018) reported the estimated rates of new HIV diagnoses for MSM in the United States in 2014 by race/ethnicity. Reported numbers and rates per 100 of new HIV diagnoses among MSM, by race, and ethnicity were used to calculate total MSM population sizes.

^h^ Estimated so that prevalence among MSM with multiple HIV transmission risk factors equaled 23.5%, approximating estimates of prevalence among MSM prevalence of 23.4% from NHBS 2017 (CDC, 2019b).

^i^ Estimated so that prevalence among HETs with fewer HIV transmission risk factors equaled 0.06%, approximating estimates of prevalence among HETs with fewer HIV transmission risk factors (defined as no history of MSM contact or sexually transmitted diseases) of 0.06% Woodring et al. (2015), using NHANES 2007-2012 data. Relative prevalence values were differentiated by race, based on expert opinion.

Table 4.2.	HIV Prevalence in Initial Population (2010)

| Transmission group / age | Female | | | Male | | | Source |
| --- | --- | --- | --- | --- | --- | --- | --- |
|  | Black | Hispanic | White/other | Black | Hispanic | White/other |  |
| HET |  |  |  |  |  |  |  |
| 13-24 | 0.22% | 0.04% | 0.02% | 0.04% | 0.01% | 0.00% | CDC (2019a, 2018b, 2019e)^a^ |
| 25-34 | 0.88% | 0.15% | 0.05% | 0.21% | 0.05% | 0.01% |  |
| 35-44 | 1.44% | 0.29% | 0.09% | 0.66% | 0.14% | 0.03% |  |
| 45-54 | 1.41% | 0.41% | 0.08% | 1.23% | 0.29% | 0.04% |  |
| 55-64 | 0.65% | 0.24% | 0.03% | 0.55% | 0.21% | 0.03% |  |
| 65+ | 0.20% | 0.09% | 0.01% | 0.22% | 0.08% | 0.01% |  |
| PWID |  |  |  |  |  |  |  |
| 13-24 | 4.78% | 1.68% | 1.18% | 1.43% | 1.02% | 0.64% | CDC (2019a, 2018b, 2019e)^a^ |
| 25-34 | 20.11% | 11.09% | 9.26% | 3.14% | 4.81% | 2.04% |  |
| 35-44 | 18.62% | 10.91% | 9.50% | 7.81% | 9.75% | 4.57% |  |
| 45-54 | 24.70% | 15.08% | 14.00% | 17.43% | 16.40% | 8.45% |  |
| 55-64 | 24.28% | 11.09% | 7.35% | 18.12% | 8.79% | 5.56% |  |
| 65+ | 13.83% | 7.61% | 4.36% | 24.85% | 11.88% | 8.59% |  |
| MSM |  |  |  |  |  |  |  |
| 13-24 |  |  |  | 20.39% | 5.46% | 2.19% | CDC (2019a, 2018b, 2019e)^a^ |
| 25-34 |  |  |  | 36.32% | 14.60% | 7.16% |  |
| 35-44 |  |  |  | 38.88% | 22.33% | 13.73% |  |
| 45-54 |  |  |  | 38.62% | 25.56% | 17.89% |  |
| 55-64 |  |  |  | 20.72% | 14.03% | 8.19% |  |
| 65+ |  |  |  | 8.65% | 5.72% | 2.84% |  |

Note: CDC = Centers for Disease Control and Prevention; HET = heterosexual; HIV = human immunodeficiency virus; MSM = men who have sex with men; N/A = not applicable; NHBS = National HIV Behavioral Surveillance; PWH = people with HIV; PWID = people who inject drugs

^a^ Surveillance data (CDC, 2019a) reported counts of PWH by age group (13-24, 25-34, 35-44, 45-54, and 55+), by sex, and by race/ethnicity (Black, Hispanic, white, all races). Values for MSM by race/ethnicity and age group were from surveillance data in CDC (2019a); MSM+PWID from 2019 surveillance data were allocated to the separate categories of MSM and PWID using age group and race/ethnicity-specific assumption based on the percentage of MSM and MSM+PWID who were MSM alone. Youth (13-24 years) surveillance data from CDC (2018b) were used to estimate the allocation of Black and Hispanic PWH in the 13-24-year age group across transmission groups by sex. Similarly, surveillance data for older adults (50+ years; CDC, 2018c) were used to calculate the allocation of 55+ Black and Hispanic PWH across transmission groups by sex and by age group (55-64 and 65+). For ages 25-54, male Black and Hispanic PWH were allocated to HET and PWID transmission groups using percentage allocations to HET by age group from surveillance data; female Black and Hispanic PWH were similarly allocated to HET or PWID transmission groups. Counts of the number of other race PWH by sex, age group, and transmission group were obtained by subtracting Black and Hispanic counts from surveillance totals. Prevalence calculated as percentage of the U.S. population were calculated using sex, age group, race/ethnicity, and prevalence counts divided by the total sexually-active population sizes used in HOPE for each subpopulation defined by transmission group, sex, race/ethnicity, and age group. For HETs and MSM, the prevalence percentages overall were then used in conjunction with “Relative HIV prevalence for people with multiple vs. fewer HIV transmission risk factors” (Table 4.1) to calculate HIV prevalence for people with multiple and fewer HIV transmission risk factors separately.

Table 4.3.	Percentage Circumcised, and Percentage of Each Transmission Group Sexually Active in Initial Population (2010)

| Parameter | Value | Source |
| --- | --- | --- |
| Percentage of males circumcised, by race/ethnicity |  | Introcaso et al. (2013) |
| Black | 75.7% |  |
| Hispanic/Latino | 44.0% |  |
| White/other | 90.8% |  |
| Percentage of population sexually active, by transmission risk group | | |
| HET | 88.9% | Percentage calculated from Lansky et al., (2015)^a^ |
| MSM | 100% | Assumption |
| PWID | 100% | Assumption |

Note: HET = heterosexual; NHANES = National health and Nutrition Examination Survey; MSM = men who have sex with men; PWID = people who inject drugs

^a^ Lansky et al. (2015) estimated that 86.7% of the US population aged 13 years and older is sexually active heterosexuals (individuals who reported having an opposite sex partner within lifetime but not having had same-sex intercourse with a man or injected drugs). The percentage of HETs who are sexually active was calculated based on that percentage, 2010 Census estimates of number of individuals ages 13+ (U.S. Census Bureau [2010]), and total HET population size (where total HET population estimate is described in Table 4.1).

Table 4.4.	Distribution of Initial HIV-Infected Population across Continuum-of-Care Stages by Race/Ethnicity for All Subpopulations besides MSM ages 13-34 (2010)

| Continuum-of-Care Stage | Black | Hispanic/ Latino | White/ Other | Source |
| --- | --- | --- | --- | --- |
| Unaware (*r* = 1) | 16.7% | 17.4% | 13.9% | Percentages calculated from Gardner et al. (2011) ^a^ |
| Aware (*r* = 2) | 32.6% | 30.4% | 28.5% |  |
| Linked to HIV care (*r* = 3) | 20.9% | 18.1% | 22.1% |  |
| ART-not-VLS (*r* = 4) | 8.2% | 7.2% | 5.1% |  |
| VLS (*r* = 5) | 21.6% | 26.9% | 30.4% |  |

Note: ART = antiretroviral therapy; VLS = viral load suppressed

^a^ The non-race-specific data from Gardner et al. (2011) was adjusted to be race-specific by calculating the relative proportion of the race in each of these stages using the race-specific distribution of population in 2010. The weighted average was set to equal the original Gardner (2011) value.

Table 4.5.	Distribution of Initial HIV-Infected Population across Continuum-of-Care Stages by Race/Ethnicity for MSM ages 13-34 (2010)

| Continuum-of-Care Stage | Black  13-24 | Black  25-34 | Hispanic/ Latino  13-24 | Hispanic/ Latino  25-34 | Other  13-24 | Other  25-34 | Source |
| --- | --- | --- | --- | --- | --- | --- | --- |
| Unaware (*r* = 1) | 62.7% | 26.8% | 70.0% | 34.7% | 70.3% | 19.8% | CDC (2019a)^a^ |
| Aware (*r* = 2) | 14.5% | 26.2% | 10.3% | 19.2% | 9.4% | 21.6% | Calculated ^b^ |
| Linked to HIV care (*r* = 3) | 9.3% | 16.7% | 6.1% | 11.4% | 7.3% | 16.7% |  |
| ART-not-VLS (*r* = 4) | 3.7% | 6.6% | 2.5% | 4.6% | 1.7% | 3.8% |  |
| VLS (*r* = 5) | 9.8% | 23.6% | 11.1% | 30.0% | 11.3% | 38.0% | Unpublished CDC data^c^ |

Note: ART = antiretroviral therapy; HIV = human immunodeficiency virus; VLS = viral load suppressed

^a^ Estimates provided for 13-24 and 25-34. Assumed same rates for ages 13-17 and 18-24. Derived values for Other race/ethnicity based on overall, Black, and Hispanic

^b^ Determined the values for rows 2-4 by subtracting the percent VLS from the overall percent diagnosed and proportioning the values consistent with the overall continuum-of-care proportions by race.

^c^ Percent VLS calculated from internal analyses by the Quantitative Sciences and Data Management Branch using surveillance data. Based on HIV viral suppression at most recent viral load test 2010-2016, among males aged ≥13 years with HIV infection attributed to male-to-male sexual contact, from 19 jurisdictions.

Table 4.6.	Distribution of Initial HIV-Infected Population across HIV Stages, by Continuum-of-Care Stage (2010)

| Continuum-of-Care Stage | Acute | CD4  > 500 | CD4 350–500 | CD4  200–350 | CD4  < 200 | Source |
| --- | --- | --- | --- | --- | --- | --- |
| Unaware  (*r* = 1) | 2.0% | 28.3% | 17.0% | 17.0% | 35.7% | Crepaz et al. (2020)^a^ |
| Aware  (*r* = 2) | 2.0% | 46.1% | 17.0% | 17.0% | 17.8% | Crepaz et al. (2020)^b^ |
| Linked to HIV care (*r* = 3) | 0.0% | 37.5% | 37.5% | 15.0% | 10.0% | Assumption^c^ |
| ART-not-VLS (*r* = 4) | 0.0% | 27.8% | 34.4% | 24.4% | 13.4% | CDC (2014)^d^ |
| VLS (*r* = 5) | 0.0% | 46.4% | 22.8% | 17.4% | 13.4% | CDC (2014) ^e^ |

Note: ART = antiretroviral therapy; VLS = viral load suppressed

^a^ Crepaz et al. (2020) reported that among those with a reported CD4 value, 30.3% were 500+, 34.1% 200-499, 35.7% <200. Two percentage points of the 30.3% with CD4>500 were assumed to be acute. The percentage of those with CD4 200-499 was split evenly between 200-350 and 350-500. Values for CD4 200-500 were adjusted slightly so that 35.7% had CD4<200..

^b^ Applied same source as for unaware (Crepaz et al. [2020]) but assumed that half of individuals with Chronic HIV Infection and CD4<200 would have been linked to care, therefore shifting the distribution toward higher CD4 counts; that was reflected by increasing the percentage with CD4>500.

^c^ Assumed that none had acute, a small amount (10%) had CD4<200, the vast majority (75%) would be in earlier non-symptomatic stages of HIV, and the remainder would have CD4 200-350.

^d^ Assumed same distribution as VLS from CDC (2014), but based on the recommendations in 2010, fewer people with CD4>500 would have been prescribed ART. Therefore, 25% of CD4>500 reported in CDC (2014c) were shifted to 350-500, and 15% of CD4>500 to 200-350

^e^ From CDC (2014) Table 3 "Stage of disease and CD4 counts of patients during the 12 months before the interview—Medical Monitoring Project, United States, 2010." Used percentage of patients with geometric mean CD4 count in each range. The MMP population includes adult PWH who received care from known providers of outpatient HIV medical care in the United States.

### 5. Movement into and out of the Model, between Subpopulations, between Compartments (Except Due to Infection), and within Compartments

The number of individuals in the model’s population changes over time by individuals aging into the population, dying with CD4<200, or dying with HIV with CD4≥200 or people without HIV moving between subpopulations solely due to aging. They move between compartments due to disease progression and progression along the HIV care continuum. The values of the parameters that affect these dynamics are specified in Tables 5.1 through 5.5. Many of these parameter values were calibrated within defined ranges to match specific target outcomes; further details on the calibration process are provided in Section 10.1.

#### 5.1 Transitions into and out of the model

Individuals can only enter the model by aging into the population (at either age 13 or 18, depending on user settings about whether age 13–17 is included in the population). All enter as people without HIV and not on PrEP (*c* = 1).

The number entering the population by transmission group, sex, race/ethnicity, and number of HIV transmission risk factors will vary over time and will be determined as the product of (1) the current number of people without HIV in the youngest age group of the same sex and race/ethnicity, number of HIV transmission risk factors, and circumcision status (as determined by inputs specified in Section 4); (2) an overall entry rate; and (3) relative adjustment values to annual overall entry rate by sex and race/ethnicity.

If the 13–17-year-old age group is included in the modeled population, the overall rate of aging into the population is equal to 0.22 per person in the 13 to 17 age group in the initial population (Table 5.1). This is calculated as 1 ÷ 5 years in the 13 to 17 age group, and then adjusted slightly to keep the population stable over time. If the 13 to 17 age group is not included, the overall rate of aging in is equal to 0.1429 per person in the 18 to 24 age group in the initial population (where 0.1429 = 1 ÷ 7 years in the 18 to 24 age group) (Table 5.1).

The relative adjustments to the annual overall entry rate by sex and race/ethnicity (Table 5.1) were determined so that the total population sizes in 2016 by sex and race/ethnicity approximately matched 2016 population estimates for those same categories in U.S. census data. As a result of this calculation method, the distribution of individuals across demographic subpopulations approximately reflects the changing race/ethnicity distribution of the U.S. population.

Individuals leave the model by dying (death among persons with CD4<200 or among all other model compartments). Death leads them to either the “death among PWH with CD4<200” (if they had CD4<200 at death) or “death among people without HIV and PWH with CD4≥200” states (if they had CD4≥200 or were without HIV at death); individuals move to the “death among PWH with CD4<200” stage only from the HIV infection with CD4<200 stage (CD4 < 200; *h* = 5). Mortality rates are determined by four methods; the values and sources for the inputs used to calculate those rates are listed in Table 5.1:

- For individuals with CD4<200 and not on ART (*c* = 24 to 26), their mortality rate is equal to 1 / the *Number of years in each stage for PWH not on ART* for CD4<200.
- For PWH with VLS (*c* = 13, 18, 23, 28), their mortality rates are equal to *Annual probability of death for PWH with VLS, by disease stage.*
- For all other individuals who are HIV-infected (*c* = 6 to 12, 14 to 17, 19 to 22, 27), their mortality rates are equal to *Annual probability of death for PWH with VLS, by disease stage* times a multiplier that is specific to the continuum-of-care stage (*r*).
- For all people without HIV (*c* = 1, 2, 3, 4, 5), mortality is assumed to be based on Life Tables in Arias (2012), as specified by *Annual probability of death for people without HIV.*

Effects of the COVID-19 pandemic on mortality may be applied in time periods 2 to 4. The same effects are applied to mortality rates for people without HIV and mortality rates for all PWH except individuals not on ART with CD4<200. Effects of the pandemic are also considered on rates of progression along the HIV care continuum (details in Section 5.4.1), PrEP initiation (details in Section 5.5), and sexual behaviors (details in Section 6.2).

Table 5.1.	Rate of Aging into Population and Inputs that Determine All HIV Progression and Death Rates

| Parameter | 13–17 Years | 18–24 Years | 25–34 Years | 35–44 Years | 45–54 Years | 55–64 Years | 65+ Years | Source |
| --- | --- | --- | --- | --- | --- | --- | --- | --- |
| Annual overall rate of aging into population per person in youngest age group | | | | | | | | |
|  | ------------0.22 if youngest age group = 13–17-------------  -------------0.1429 if youngest age group = 18–24------------ | | | | | | | Assumed^a^ |
| Relative adjustment to annual overall entry rate by sex and race/ethnicity | | | | | | | | |
|  | Black males: -7.5%  Hispanic/Latino males: 5.0%  Other males: −1.5%  Black females: −2.0%  Hispanic/Latino females: 9.0%  Other females: -1.5% | | | | | | | Determined so that the total population sizes projected in HOPE in 2016 by sex and race/ethnicity approximated 2016 population estimates for those same categories in U.S. census data, adjusting to include only sexually active individuals (U.S. Census Bureau, 2017)^b^ |
| Number of years in each stage for PWH not on ART (*r* = 1, 2, 3) | | | | | | | | |
| Acute | -----------------------------0.17---------------------------------- | | | | | | | Fiebig et al. (2003) |
| CD4 > 500 | -----------------------------5.00---------------------------------- | | | | | | | Set by CDC so that the numbers of deaths would align with surveillance data when the distribution of PWH along the continuum of HIV care were also in line with surveillance data |
| CD4 350–500 | -----------------------------5.00---------------------------------- | | | | | | |  |
| CD4 200–350 | -----------------------------5.00---------------------------------- | | | | | | |  |
| CD4 < 200^c^ | -----------------------------3.00---------------------------------- | | | | | | | Juusola et al. (2012), increased to 3 years to produce # of deaths consistent with CDC surveillance data. |

(continued)

Table 5.1.	Rate of Aging into Population and Inputs that Determine All HIV Progression and Death Rates (continued)

| Parameter | 13–17 Years | 18–24 Years | 25–34 Years | 35–44 Years | | 45–54 Years | | 55–64 Years | | 65+ Years | Source |
| --- | --- | --- | --- | --- | --- | --- | --- | --- | --- | --- | --- |
| Adjustment factor to number of years in each CD4 stage if not on ART, by age group | | | | | | | | | | |  |
|  | 1.00 | 0.95 | 0.88 | 0.80 | | 0.75 | | 0.72 | | 0.72 | Hall et al., (2007) provides the percentage of people without AIDS 1 and 3 years after HIV diagnosis and the percentage of people surviving 1 and 3 years after AIDS diagnosis for different age groups. Factors relative to the 13–17 age group were calculated from the data. |
| Annual rate of progressing one disease stage (to lower CD4 count) if on ART, but not VLS (*r* = 4) | | | | | | | | | | | |
| CD4 > 500 | ----------------*0.0317*---------------- | | | | ------*0.0257*----- | | | | *0.0308* | | Determined by calibration^d^ |
| CD4 350–500 | ---------------*-0.0410*---------------- | | | | -----*-0.0239*----- | | | | *0.0226* | |  |
| CD4 200–350 | ----------------*0.0212-*--------------- | | | | ------*0.0446*----- | | | | *0.0619* | |  |
| Annual rate of progressing by one disease stage (to lower CD4 count) while VLS (*r* = 5) | | | | | | | | | | |  |
| CD4 > 500 | ------------------------------ *0.0389*-------------------------------- | | | | | | | | | | Determined by calibration^d^ |
| CD4 350–500 | ------------------------------ *0.0393*-------------------------------- | | | | | | | | | |  |
| CD4 200–350 | ------------------------------ *0.0315*-------------------------------- | | | | | | | | | |  |
| Annual rate of improving one disease stage (to higher CD4 count) while VLS (*r* = 5) | | | | | | | | | | |  |
| CD4 350–500 | ----------------*0.5067*---------------- | | | | | | ---*-0.4734*------ | | *0.4575* | | Determined by calibration^d^ |
| CD4 200–350 | ----------------*0.5295*---------------- | | | | | | ---*-0.4949*------ | | *0.4723* | |  |
| CD4 < 200 | ----------------*0.4900*---------------- | | | | | | ---*-0.5117*------ | | *0.4922* | |  |
| Annual rate of death for PWH with VLS, by disease stage (*r* = 5; range across race/ethnicities) | | | | | | | | | | | |
| CD4 > 200 | 0.0039 | 0.0039 | 0.0041 | 0.0067 | | 0.0126 | | 0.0138 | | 0.0296 | NA-ACCORD (2014)^e^  Bosh et al. (2020)^e^ |
| CD4 < 200 | 0.0071 | 0.0071 | 0.0075 | 0.0088 | | 0.0220 | | 0.0240 | | 0.0537 |  |

(continued)

Table 5.1.	Rate of Aging into Population and Inputs that Determine All HIV Progression and Death Rates (continued)

| Parameter | 13–17 Years | 18–24 Years | 25–34 Years | 35–44 Years | 45–54 Years | 55–64 Years | 65+ Years | Source |
| --- | --- | --- | --- | --- | --- | --- | --- | --- |
| Relative risk of death vs. PWH who are VLS (may vary between time period 1 and time periods 2-5, but same values currently used) | | | | | | | | Assumed based on Krebs et al. (2019), Krentz et al. (2014), Krueger et al. (2019)^f^ |
| Not on ART (*r* = 1,2,3) | -----------------------------1.20------------------------------ | | | | | | |  |
| On ART, but not VLS (*r* = 4) | -----------------------------1.20------------------------------ | | | | | | |  |
| Relative risk of death for PWID without HIV vs. non-PWID without HIV | | | | | | | | |
| PWID | -----------------------------2.54------------------------------- | | | | | | | Vlahov et al. (2008)^g^ |
| Annual probability death for people without HIV | | | | | | | | |
| HET or MSM |  | | | | | | |  |
| Female | 0.0002 | 0.0004 | 0.0006 | 0.0013 | 0.0030 | 0.0063 | 0.0433 | Arias (2012) |
| Male | 0.0005 | 0.0013 | 0.0014 | 0.0021 | 0.0049 | 0.0105 | 0.0472 |  |
| PWID |  | | | | | | |  |
| Female | 0.0006 | 0.0011 | 0.0016 | 0.0032 | 0.0075 | 0.0160 | 0.1099 | Calculated^g^ |
| Male | 0.0013 | 0.0032 | 0.0036 | 0.0054 | 0.0124 | 0.0266 | 0.1199 | Calculated^g^ |
| Percent increase in annual mortality rates (regardless of HIV status) by age group due to COVID-19, by time period (time periods 2 to 4 only) | | | | | | | | Kochanek, et al (2020), Ahmad et al, 2021^h^ |
| Time period 2 | 18.5% | 18.5% | 22.6% | 23.6% | 19.2% | 16.4% | 14.7% |  |
| Time period 3 | 18.5% | 18.5% | 22.6% | 23.6% | 19.2% | 16.4% | 14.7% |  |
| Time period 4 | 4.6% | 4.6% | 5.7% | 5.9% | 4.8% | 4.1% | 3.7% |  |

Table 5.1.	Rate of Aging into Population and Inputs that Determine All HIV Progression and Death Rates (continued)

Note: ART = antiretroviral therapy; B = black; H = Hispanic/Latino; O = white/other race; CDC = Centers for Disease Control and Prevention; HET = heterosexual; HIV = human immunodeficiency virus; HOPS = HIV Outpatient Study; MSM = men who have sex with men; NA-ACCORD = North American AIDS Cohort Collaboration on Research and Design; NCHS = National Center for Health Statistics; PWH = people with HIV; PWID = people who inject drugs; VLS = viral load suppressed

^a^ If the youngest age group is 13–17, the overall entry rate was set approximately equal to 1 ÷ (number of years in the 13-17 age group) and adjusted so that the population size was stable over time (until 2015). If the youngest age group is 18-24, the rate of aging into the model is equal to 1 ÷ (number of years in the 18-24 age group).

^b^ Estimated so that 2016 HOPE population sizes approximated 2016 census data of 13,327,093 for Black males, 18,938,691 for Hispanic/Latino males, 80,285,306 for white/other males, 14,759,511 for Black females, 18,484,810 for Hispanic/Latino females, 83,398,464 for white/other females, adjusted to include only sexually-active and ages 13+ years.

^c^ Departure from CD4 < 200 stage leads to death among persons with CD4<200

^d^ It is assumed that PWH who are on ART but not VLS experience slowed disease progression and that those who are VLS experience mostly improvement in CD4 rather than disease progression. These assumptions were found to be mathematically essential for the model to simultaneously replicate both the incidence and prevalence trends reported in CDC surveillance data. See Section 10 for further details on the calibration process.

^e^ Source based on ages 18 or older. The probabilities are assumed to be the same for age groups 13–17, 18–24, and 25–34 and for ages 45–54 and 55–64 years. Values calculated directly from NA-ACCORD (2014) data for 2010 based on mortality rates of NA-ACCORD participants. Applied age-based multipliers from Bosh et al. (2020) to better reflect 2010-2017 trends and match surveillance data.

^f^ Assumption of mortality risk that is higher for PWH who are not VLS versus those who are supported by Krebs et al. (2019), Krentz et al. (2014), Krueger et al. (2019), as well as unpublished data from the HOPS trial (an NA-ACCORD clinical trial site). Specific values of multipliers were estimated to 1.2 by CDC so that the numbers of deaths would align with surveillance data when the distribution of PWH along the continuum of HIV care were also in line with surveillance data. Overall death rates still vary by rows due to differences in mortality by CD4 and the variation in distribution between CD4 stages by row.

^g^ Calculated as the risk from CDC/NCHS life tables multiplied by the relative risk of death for PWID versus non-PWID population.

^h^ Change in mortality from 2019 to 2020 estimated 2019 mortality data extracted from Kochanek, et al (2020) and 2020 mortality extracted from MMWR (Ahmad et al, 2021). The rate per person 65+ was calculated by dividing total persons 65+ by the number of deaths between ages 65-74, 75-84, 85+. The same increases are applied in 2020 and 2021 and a quarter of those increases in 2022. The model includes a setting to indicate whether to apply COVID effects on mortality rates in model calculations. The default setting is “no” so, by default, the effects listed in this table are not applied.

Table 5.2.	Inputs for Calculating Rates of Undiagnosed HIV-Infected and People without HIV Getting Tested in Time Periods 1, 2-4, and 5^a^

| Parameter | Value | Source |
| --- | --- | --- |
| Annual rate of PWH getting tested for reference case (HET, Black, CD4 > 500, age 13-17) | | |
|  | *0.0790* | Determined by calibration |
| Multiplier for annual rate of getting tested by HIV stage, race/ethnicity, transmission group, and age group versus the reference case | | |
| HIV status (reference: non-acute, CD4 > 500) |  | |
| Without HIV | 1.41 | Derived from CDC (2013e); CDC (2017a)^b^ |
| Acute | 0.9613 | Determined by calibration |
| CD4 350–500 | 2.1291 |  |
| CD4 200–350 | 5.2216 |  |
| CD4 < 200 | 4.3662 |  |
| Race/ethnicity (reference: Black) |  | Determined by calibration |
| Hispanic/Latino | 0.8640 |  |
| White/other | 1.2127 |  |
| Transmission group (reference: HET) |  | Determined by calibration |
| PWID | 4.0310 |  |
| MSM | 4.6357 |  |
| Age group (reference: 13–17) |  |  |
| 18–24 | 0.6348 | Determined by calibration |
| 25–34 | 2.1245 |  |
| 35–44 | 2.2451 |  |
| 45–54 | 2.2803 |  |
| 55–64 | 2.2693 |  |
| 65+ | 2.1262 |  |

Note: HET = heterosexual; HIV = human immunodeficiency virus; MSM = men who have sex with men; PWH = people with HIV; PWID = people who inject drugs

^a^ All inputs in this tables have separate input cells for time periods 1, 2-4, and 5. The default values of those inputs are the same across those three sets of periods.

^b^ Estimated so that the percentage of people without HIV getting tested across all races and transmission groups is consistent with published data on the total number of tests conducted and the percentage of HIV tests that are positive (CDC [2013e]; CDC [2017]). It is calculated as a function of the rate of PWH getting tested for the reference case, which is calibrated, and therefore varies between calibration sets.

Table 5.3.	Testing Performance Parameters

| Input | Value | Source |
| --- | --- | --- |
| Test sensitivity,^a^ acute HIV |  |  |
| Rapid screen | 0.0173 | Average of Oraquick and Clearview tests from Pilcher et al. (2013)^b^ |
| Conventional screen | Before 2016: 0.5090 | Before 2016: Hutchinson et al. (2013) (3G test) |
|  | 2016 and after: 0.8276 | 2016 and after: Hutchinson et al. (2013), which cited Chavez et al. (2011) (4G test) |
| Second test | Before 2016: 0.0  2016 and after: 0.4522 | Before 2016: Assumption based on length of Western blot window period (Feibig et al., 2003) 2016 and after: Assumes Geenius HIV 1/2 supplemental assay (Bio-Rad, 2014) |
| Test sensitivity, chronic HIV^c^ |  |  |
| Rapid screen | Before 2016: 0.995  2016 and after: 0.998 | Before 2016: OraQuick ADVANCE: Rapid HIV-1/2 Antibody Test package insert. Average of 2 specimen types: sensitivity Oral Fluid (0.993) - Whole blood/finger stick (0.997)  2016 and after: Reflects more sensitive rapid tests: Unigold =0.997, Insti=0.998, Determine=0.998, Clearview =0.999, Chembio = 0.998 (0.997-0.999) |
| Conventional screen | Before 2016: 0.9968  2016 and after: 0.9986 | Before 2016: Hutchinson et al. (2013), which cited package insert data (3G)  2016 and after: Hutchinson et al. (2013), which cited Chavez et al. (2011) (4G) |
| Second test | Before 2016^d^: 1.0  2016 and after: 0.998 | Before 2016: Assumption in Hutchinson et al. (2013)  2016 and after: Geenius HIV 1/2 supplemental assay package insert. Average of 3 specimen types. |

^a^ The window period during which acute HIV is not detectable is factored into the calculation of test sensitivity for individuals with acute HIV.

^b^ Calculated as average test sensitivity for Oraquick and Clearview tests for acute HIV.

^c^ Early infection, the time after acute infection but before viral set point when transmission risk per contact is elevated and testing is less sensitive is not accounted for in the model.

^d^ Because all negative second tests are followed up by a third test (nucleic acid amplification test [NAT]) with 100% sensitivity, the sensitivity of the screening process to determine progression does not consider sensitivity of the second test. The only effect is on cost.

Table 5.4.	Annual Probability of Initiating ART in Time Periods 1, 2-4, and 5^a^

| Parameter | Acute | CD4 > 500 | CD4  350–500 | CD4  200–350 | CD4 < 200 | Source |
| --- | --- | --- | --- | --- | --- | --- |
| Annual probability of initiating ART, without adjustment for relative risk of initiating ART by race/ethnicity | | | | | | |
|  | 0.000 | 0.540 | 0.600 | 0.750 | 0.920 | Fleishman et al. (2012)^b^ |
| Relative risk of initiating ART, by race/ethnicity | | | | | | |
| Black | ---------------------------------------------*1.0007*-------------------------------------- | | | | | Determined by calibration |
| Hispanic | ---------------------------------------------*0.9856*-------------------------------------- | | | | |  |
| White/other | ---------------------------------------------*1.0073*-------------------------------------- | | | | |  |

Note: ART = antiretroviral therapy; HIV = human immunodeficiency virus; PWH = people with HIV

^a^ All inputs in this tables have separate input cells for time periods 1, 2-4, and 5. The default values of those inputs are the same across those three sets of periods.

^b^ Examined ART use as a function of sex, race/ethnicity, HIV risk group, age, and CD4 history (no test< 500 cells/mm^3^, one or more tests between 500 and 350 cells/mm^3^, one test ≤ 350 cells/mm^3^, and two or more tests ≤ 350 cells/mm^3^). Fleishman et al., (2012) reported the proportion of patients in care who initiated ART in 2008 with CD4 levels of interest based on the HIV Research Network (HIVRN) study of PWH (> 18 years of) age who first presented for clinical care during the period from January 1997 to December 2007. We assumed that one test with ≤ 350 cells/mm^3^ is approximation for CD4 250-350 in our model and two tests with ≤ 350 cells/mm^3^ is for CD4<200.

Table 5.5.	Other Continuum-of-Care Probabilities

| Parameter | Black | Hispanic | White/other | Source |
| --- | --- | --- | --- | --- |
| Percentage of tests performed in nonclinical (vs. clinical) settings | -------------------------16%------------------------------- | | | CDC (2013e) |
| Percentage of tests that are rapid vs. conventional (same values applied for 2010-2015 and 2016+) | | | |  |
| Clinical setting | 50% | 50% | 50% | Assumption based on Huang et al. (2016) |
| Nonclinical setting | 100% | 100% | 100% | Assumed that all tests in nonclinical setting are rapid |
| Probability of being notified of status if tested with the following: | | | | |
| Conventional test | -------------------------0.80------------------------------- | | | Huang et al. (2016) |
| Rapid test | -------------------------1.00------------------------------- | | |  |
| Annual probability of diagnosed individual linked to HIV care at diagnosis | | | | |
|  | 0.790 | 0.830 | 0.860 | CDC (2015) |
| Annual probability of diagnosed individual linked to HIV care each year after first year if CD4>350 (*h* = 1, 2, 3)^a^ | | | | |
|  | *0.1457* | *0.1611* | *0.1356* | Determined by calibration |
| Relative risk of linkage to HIV care, after diagnosis if CD4 ≤ 350, by disease stage (reference: CD4>350)^a^ | | | | |
| CD4 200–350 (*h* = 4) | ------------------------------*6.7088*------------------------ | | | Determined by calibration |
| CD4 < 200 (*h* = 5) | ------------------------------*5.5057*------------------------ | | |  |
| Annual probability of dropping out of care if linked to HIV care (from *r* = 3 to *r* = 2)^a^ | | | |  |
|  | 0.4689 | 0.4674 | 0.4472 | Determined by calibration |
| Annual probability of dropping out of ART and moving to linked-to-HIV-care (from *r* = 4 to *r* = 3), by race/ethnicity^a^ | | | | |
|  | 0.2456 | 0.2524 | 0.1942 | Determined by calibration |

(continued)

Table 5.5.	Other Continuum-of-Care Probabilities (continued)

| Parameter | Black | Hispanic | White/other | Source |
| --- | --- | --- | --- | --- |
| Relative risk of dropping out of ART and moving to linked-to-HIV-care (from *r* = 4 to *r* = 3), by age group (versus annual probability by race/ethnicity, reported above)^a^ | | | | |
| 13–17 | ------------------------------*1.12*------------------------- | | | Calculated from odds ratio found by Sabin et al. (2009) (for the odds of discontinuing HAART within the first year for reasons other than virological failure) and from estimates of annual probability of dropping off ART if on ART-not-VLS, which are determined by calibration. |
| 18–24 | ------------------------------*1.12*------------------------- | | |  |
| 25–34 | ------------------------------*1.06*------------------------- | | |  |
| 35–44 | ------------------------------*1.03*------------------------- | | |  |
| 45–54 | ------------------------------*1.10*------------------------- | | |  |
| 55–64 | ------------------------------*1.14*------------------------- | | |  |
| 65+ | ------------------------------*1.14*------------------------- | | |  |
| Annual probability of dropping off of ART if on ART, but not VLS (from *r* = 4 to *r* = 2)^b^ and moving to Aware stages (among those on ART and not VLS who are not in care) ^a^ | 0.000 | 0.000 | 0.000 | Flow not considered in base model. |
| Annual probability of loss of VLS if VLS (from *r* = 5 to *r* = 4) ^a^ | 0.2502 | 0.1370 | 0.1917 | Determined by calibration |
| Relative risk of loss of VLS if VLS (from *r* = 5 to *r* = 4), by transmission group (reference: HET) ^a^ | | | |  |
| MSM | ------------------------------*2.7241*------------------------ | | | Determined by calibration |
| PWID | ------------------------------*2.2686*------------------------ | | |  |
| Relative risk of loss of VLS if VLS (from *r* = 5 to *r* = 4), by age group (reference: 13–17) ^a^ | | | |  |
| 18–24 | ------------------------------*2.1103*------------------------ | | | Determined by calibration |
| 25–34 | ------------------------------*3.3217*------------------------ | | |  |
| 35–44 | ------------------------------*2.9540*------------------------ | | |  |
| 45–54 | -----------------------------*-0.9078*------------------------ | | |  |
| 55–64 | ------------------------------*0.8251*------------------------ | | |  |
| 65+ | ------------------------------*0.9865*------------------------ | | |  |

(continued)

Table 5.5.	Other Continuum-of-Care Probabilities (continued)

| Parameter | Black | Hispanic | White/other | Source |
| --- | --- | --- | --- | --- |
| Annual probability of transitioning from VLS to LTC no ART affects (from r = 5 to r = 3) ^a,b^ | 0.000 | 0.000 | 0.000 | Flow not considered in base model. |
| Annual probability of transitioning from VLS to Aware (from r = 5 to r = 2)^a,b^ | 0.000 | 0.000 | 0.000 | Flow not considered in base model. |
| Annual probability of becoming VLS if ART-not-VLS (from r = 4 to r = 5) ^a^ | 0.5307 | 0.4728 | 0.6064 | Determined by calibration. |
| Relative risk of becoming VLS if on ART but not VLS (from r = 4 to r = 5), by transmission group (reference: HET) ^a^ | | | | |
| MSM | ------------------------------*2.5789*------------------------- | | | Determined by calibration |
| PWID | ------------------------------*0.8112*------------------------ | | |  |
| Relative risk of becoming VLS if ART-not-VLS (from r = 4 to r = 5), by age group (reference: 13–17) ^a^ | | | | |
| 18–24 | ------------------------------*1.4856*------------------------ | | | Determined by calibration |
| 25–34 | ------------------------------*0.7746*------------------------ | | |  |
| 35–44 | ------------------------------*2.8782*------------------------ | | |  |
| 45–54 | ------------------------------*2.7286*------------------------ | | |  |
| 55–64 | ------------------------------*2.6828*------------------------ | | |  |
| 65+ | ------------------------------*3.2693*------------------------ | | |  |
| Percentage of individuals (from linked to HIV care) who become viral load suppressed at ART initiation | ---------------------------80.0%--------------------------- | | | Althoff et al. (2010) |

Note: ART = antiretroviral therapy; CDC = Centers for Disease Control and Prevention; VLS = viral load suppressed

^a^ This input has separate input cells for time periods 1, 2-4, and 5. The default values of this input are the same across those three sets of periods.

^b^ Because all values for these transitions are currently 0, the arrows representing these transitions are not included in the current model flow diagram

#### 5.3 Transitions between Subpopulations (Aging Only)

Individuals can only transition between demographic subpopulations by aging. That is, all of an individual’s demographic characteristics remain the same for the duration of the model except their age. In general, one would expect a rate of aging out of an age group equal to the reciprocal of the number of years in each interval. However, we apply to that basic rate of aging a multiplicative factor. This factor is required since HOPE is not an individual-based model and progression involves people from all age groups, so aging cannot occur linearly. The factor, therefore, allows the effective average age of individuals in each interval to stay within the interval that it represents (e.g. 13-17 years, 18-24 years) rather than the effective average age shrinking due to people progressing between age groups too quickly (e.g. in six time steps people in the 13-17 age group could feasibly age to the 65+ age group). As a result, the rate of aging out of an age group *j* and into an age group *j* + 1 (where *j* < 7) is calculated as 1 ÷ [(the number of years in age group *j*) x (Adjustment factor for age group *j*)]. The values of these adjustment factors are determined by calibration so that total population sizes and total prevalence counts by age group approximate census and surveillance data, respectively. In the current version of the model, they are equal to *1.0, 1.3, 0.9, 1.7, 1.2, and 1.0* for each age group *j* = {1 to 6}.

In reality, individuals in the U.S. population move between subpopulations in other ways, such as transitioning between transmission risk groups or number of HIV transmission risk factors, or going from uncircumcised to circumcised; however, as a simplifying assumption, those transitions are not considered in this model.

#### 5.4 Transitions between Compartments Due to Disease Progression

Transitions between disease stages occur by infection and by HIV progression. The rates at which individuals transition from any disease stage *h* (where *h ≥ 1*) to *h* + 1 or *h* – 1 (or to death in the case of *h =* 5) are assumed to be constant. For stages in which patients are not on ART (*r* = 1, 2, 3), the rates from *h* to *h* + 1 are equal to 1 ÷ (duration of stage *h*). Otherwise, the rates are specified directly. For VLS stages (*r* = 5), progression may occur from disease stage *h* to *h* + 1 or to *h* – 1, but for all other continuum-of-care stages, progression may only occur from *h* to *h* + 1. All inputs that determine these progressions are specified in Table 5.1.

Individuals who are ART-not-VLS (*r =* 4) experience declines in their CD4 counts but have a slower disease progression than the people who are not on ART (*r =* 1, 2, or 3). Individuals who are VLS (*r =* 5) are very different from individuals in other continuum stages in that they can experience either increases or decreases in their CD4 counts and, in fact, they are more likely to experience an increase than a decrease in their CD4 counts.

##### 5.4.1 Role of CD4 Count in Disease Progression and Transmission

In HOPE, disease stages are defined by CD4 stratum. PWH in those stages vary by three factors: duration of time in the stage, transmission risk, and death rates.

- Duration of each stage: For PWH not on treatment (*r* = 1, 2, or 3), duration of each disease stage *h* is based on published estimates (Table 5.1). Duration of each stage when on treatment (*r* = 4 or 5) is specified by an input defining the rates of progression out of that stage (Table 5.1); those values reflect longer durations in each stage and are determined by calibration.
- Transmission risk: Except for the acute phase, higher CD4 is associated with a reduced risk of transmission (Table 6.8). Although the model does not explicitly consider viral load measurement, these CD4 strata reflect higher transmission rates associated with higher viral loads.
- Death rates: As outlined in section 5.1, the last disease stage, CD4 < 200, is associated with a higher death rate.

Because of these variations, when patients are on treatment but are not VLS (*r* = 4), transmission is effectively reduced by keeping patients in higher CD4 strata longer and the death rate reduced by keeping more patients out of the CD4 < 200 stratum. When patients become virally suppressed (*r* = 5), they tend to move to higher CD4 strata, both reducing transmission and increasing survival time. In addition, they experience the large benefits on transmission (99% reduction in this analysis) that viral suppression confers.

#### 5.5 Transitions between Compartments Due to Progression along the Care Continuum

Transitions between continuum-of-care stages occur because PWH become aware of their status through testing and notification of positive results either without immediate linkage to HIV care (*r* = 1 to *r* = 2) or with immediate linkage to HIV care (*r* = 1 to *r* = 3), are linked to HIV care after diagnosis (*r* = 2 to *r* = 3), depart from care (*r* = 3 to *r =* 2), initiate ART (*r* = 3 to *r* = 4 or *r* = 3 to *r =* 5), drop off of ART (*r =* 4 to *r =* 3), or become VLS or lose viral suppression (*r* = 4 to *r =* 5 or *r* = 5 to *r =* 4).

Individuals who depart from care are assumed to return to the aware stage, which includes individuals who have never been in care. This assumption of aggregating individuals who have never been in care with those who have dropped out of care was a simplifying assumption. The parameters that determine these transitions are outlined in Tables 5.2 to 5.5. Some inputs have values that vary by time periods; many of those inputs have the same values across periods 2-4, which by default covers the period most directly affected by the COVID-19 pandemic. The different methods that can be applied to calculate progression along the care continuum in each model time period are explained in Section 5.4.1. A detailed explanation of the methods applied for calculating testing rates is presented in Section 5.4.2.

##### 5.5.1 Methods for Calculating Progression along the Care Continuum

###### Time Period 1

In the first time period, annual rates of progression are calculated directly from user-inputted annual probabilities or rates and, in some cases, relative risk factors by subpopulation or disease stage. If probabilities of transition are entered, rates are calculated by using Equation (5.1). If relative risk factors apply, they are multiplied by the annual rates by subpopulation or disease stage as appropriate. The inputs used to determine all progression in the first time period are specified in Tables 5.2 to 5.6. Many of those were estimated through a calibration process so that their values resulted in model outcomes (e.g., the percentages of the PWH that were diagnosed and VLS and the number of new infections in one or more years) that closely matched surveillance data, as outlined in Section 9.1.

Annual rate = −ln(1 ‒ Annual probability of transition)	(5.1)

The testing rate is used in the following example to demonstrate how this method is used to calculate progression. The eligible testing pool includes all people without HIV and undiagnosed PWH. The annual rates varied by subpopulation *p* and HIV status *h* (defined by infection status and, for PWH, HIV stage) and were calculated as a product of a base rate and multipliers specific to race/ethnicity, transmission group, HIV status, and age group, as defined by Equation (5.2). Both the base rate and the multipliers were estimated through the model’s calibration process. The calibrated values that determine testing rates in the first time period are defined in Table 5.2.

$\Pi_{p}^{c}\left( t \right)$ =
(*Annual base testing rate of PWH who are HET, Black, CD4 > 500, ages 13-17 years [or ages 18-24 years if ages 13-17 years are not modeled] at t*)
x (*HIV-status testing multiplier*_c_ at time *t*)
x (Race/ethnicity testing multiplier_p_ at time *t*)
x (Transmission group testing multiplier_p_ at time *t*)
x (Age group testing multiplier_p_ at time *t*),

for *c* = {6, 9, 14, 19, 24}	(5.2)

where

- $\Pi_{p}^{c}(t)$ = rate of testing of undiagnosed individuals in compartment *c*, at time *t*, by demographic subpopulation *p*.

###### Time Periods 2 Through 4 (Incorporating COVID-19 Effects on the Continuum of Care)

The HOPE Model can consider effects of the COVID-19 pandemic on diagnosis (through testing rates), ART initiation, dropping off of ART, or loss of VLS in time periods 2 to 4. For those steps along the continuum that may be affected by the pandemic, rates of progression along the HIV care continuum in those time periods are each a product of an unadjusted rate and an effect on that rate. The unadjusted rates are the same for the three periods, but the effects may vary. The values of the unadjusted rates are set, by default, to the same rates as in the first time period. The values of the COVID-19 effects on progression in the second time period (by default, 2020) were set to reflect published data (Table 5.6). Due to a lack of data to inform the inputs after that year, COVID-19 effects in the third period (by default, 2021) were assumed to be half of the effects in the second period (by default, 2020) and no effects were assumed in the fourth period (by default, 2022).

For steps along the care continuum that may be affected by the COVID-19 pandemic, each rate of progression in those time periods is a product of the unadjusted period 2-4 rate (set, by default, to equal period 1 rates) and a multiplier incorporating the COVID-19 effects

COVID-19 disruptions may also be considered in HOPE for mortality rates (see Section 5.1), PrEP initiation (see Section 5.5), and sexual behaviors (see Section 6.2).

Table 5.6	Inputs defining COVID-19 Effects on Continuum-of-Care Progression^a^

| Input | Time period 2 (2020) | Time period 3 (2021) | Time period 4 (2022) | Sources for values / assumptions |
| --- | --- | --- | --- | --- |
| Percent reduction in annual testing rate of people without HIV due to COVID-19 | | | | Calculated from values reported in Table 1 of Patel et al. (2022) for reduction in the number of tests that did not produce a diagnosis at CDC-funded Health Departments during the COVID-19 pandemic in 2020 vs in 2019. Assumed half those reductions in 2021 and no reductions in 2022. |
| Black | 45.3% | 22.6% | 0.0% |  |
| Hispanic/Latino | 47.2% | 23.6% | 0.0% |  |
| Other | 46.1% | 23.0% | 0.0% |  |
| Percent reduction in annual testing rate of people with undiagnosed HIV due to COVID-19 (affects flow from r = 1 to r = 2 and r = 3) | | | | Set to reflect reductions in HIV diagnosis rates during COVID-19 (Viguerie et al., 2022). Assumed half those reductions in 2021 and no reductions in 2022. |
| Black | 17.0% | 8.5% | 0.0% |  |
| Hispanic/Latino | 22.0% | 11.0% | 0.0% |  |
| Other | 16.0% | 8.0% | 0.0% |  |
| Percent reduction in annual rate of initiating ART (r = 3 to r = 4 or r = 5)due to COVID-19, given eligible for ART | 4.5% | 2.3% | 0.0% | Weekly numbers of persons obtaining ARV prescriptions decreased an average of 4.5% (95% CI: -6.0% to -3.0%) over March 2020-March 2021, per Zhu, et al. (2022). Assumed half those reductions in 2021 and no reductions in 2022. |
| Percent increase in annual rate of dropping off of ART due to COVID-19 if on ART but not VLS (r = 4 to r = 3) | | | | Manually calibrated so that reduction in number of people in rows 4 and 5 together in 2020 vs versus 2019= 9% in ages 25-34, 3.8% in ages 35-44, and 2.5% overall in 2020 (to be consistent with reductions in ART prescriptions over March 2020-March 2021 reported by Zhu, et al. (2022), with only very small (1-2%) reductions in the number VLS. Assumed half those reductions in 2021 and no reductions in 2022. |
| 13-17 years | 0.0% | 0.0% | 0.0% |  |
| 18-24 years | 50.0% | 25.0% | 0.0% |  |
| 25-34 years | 100.0% | 50.0% | 0.0% |  |
| 35-44 years | 100.0% | 50.0% | 0.0% |  |
| 45-54 years | 0.0% | 0.0% | 0.0% |  |
| 55-64 years | 100.0% | 50.0% | 0.0% |  |
| 65+ years | 200.0% | 100.0% | 0.0% |  |
| Percent increase in annual rate of losing VLS and moving to on-ART-not-VLS (r = 5 to r = 4) due to COVID-19 | | | | Manually calibrated so that reduction in number of people in rows 4 and 5 together in 2020 vs versus 2019= 9% in ages 25-34, 3.8% in ages 35-44, and 2.5% overall in 2020 (to be consistent with reductions in ART prescriptions over March 2020-March 2021 reported by Zhu, et al. (2022), with only very small (1-2%) reductions in the number VLS. Assumed half those reductions in 2021 and no reductions in 2022. |
| 13-17 years | 0.0% | 0.0% | 0.0% |  |
| 18-24 years | 5.0% | 2.5% | 0.0% |  |
| 25-34 years | 5.0% | 2.5% | 0.0% |  |
| 35-44 years | 5.0% | 2.5% | 0.0% |  |
| 45-54 years | 0.0% | 0.0% | 0.0% |  |
| 55-64 years | 5.0% | 2.5% | 0.0% |  |
| 65+ years | 15.0% | 7.5% | 0.0% |  |

^a^ The model includes a setting to indicate whether to apply COVID effects on continuum of care, PrEP use, and risk behaviors in model calculations. The default setting is “no” so, by default, the effects listed in this table are not applied.

###### Time Period 5

Annual rates of progression along the care continuum are set to be equal to the same rates applied in the first time period.

##### 5.5.2 Diagnosis Rates

The annual diagnosis rate of unaware PWH is a function of the testing rate ($\pi_{p}^{c}(t)$, as determined by any of the methods used to generate testing rates), COVID effects on testing rates of PWH, the percentage of tests performed in nonclinical (vs. clinical) settings, use of rapid (versus conventional) tests in each of those settings, the types of tests used for initial and confirmatory testing (by second and third tests), sensitivity of each of those types of tests, and the likelihood of individuals getting notified of initial testing results. The diagnosis rates are calculated using Equation (5.9) and vary by time *t* and HIV status *h* (captured in compartment *c*), and by race/ethnicity, transmission group, and number of HIV transmission risk factors (captured in subpopulation p):

$\tau_{p}^{c}(t)=\left[ \pi_{p}^{c}\left( t \right) \right][1-\ddot{C}_{t}^{s=2}]\left[ \epsilon_{p}(t)*w_{c ,g=1}*\varpi_{p,g=1}(t)+\left( 1-\epsilon_{p}\left( t \right) \right)*w_{c , g=2}*\varpi_{p,g=2}(t) \right]$	(5.9)

where

- $\tau_{p}^{c}(t)$ = diagnosis rate based on test and notification of unaware PWH in compartment *c*, progressing them from unaware (*r =* 1) to aware (*r* = 2 or *r =* 3), by subpopulation *p*, for *c* = {3, 6, 11, 16, 21};
- $\ddot{C}_{t}^{s}$ = effect of COVID pandemic type s at time t for s = {1 (testing rates of people without HIV); 2 (testing rates of PWH); 3 (ART initiation rates); 4 (rates of dropping off of ART); 5 (losing VLS); 6 (PrEP initiation rates); 7 (numbers of sexual contacts); 8 (mortality rates)}
- $\epsilon_{p}(t)$ = percentage of screens that are rapid (type of test *g* = 1) across all settings, by subpopulation *p* and time *t*, calculated as:

$$\epsilon_{p}(t) =\sum_{v} (\varphi_{v}* \epsilon_{p}^{v})$$

where
  - $\varphi_{v}$ = percentage of screens that occur in setting v; and
  - $\epsilon_{p}^{v}(t)$ = percentage of screens that are rapid (type of test g = 1) in setting v, by subpopulation p and time *t*;
- $\varpi_{p,g}(t)$ = probability of notification given a confirmed positive test result for a previously undiagnosed individual in subpopulation *p* and type of test *g* at time *t*; and
- *w_c,g_ =* test sensitivity by compartment *c* and type of test *g*.

Undiagnosed PWH who are diagnosed progress either to the aware stages without immediate linkage to HIV care (*r =* 1 to *r* = 2) or to the aware stages with immediate linkage to HIV care (*r =* 1 to *r* = 3). people without HIV cannot be diagnosed and do not transition between the main compartments; if tested, they remain in the without-HIV stage (*h* = 0); the only effect of their testing in the model is to incur costs. The model assumed 100% test specificity for all tests.

#### 5.6 Transitions between Compartments Due to PrEP Participation

There are 4 PrEP states (c = 2, 3, 4, 5) in HOPE for people who have low or high adherence to PrEP and are on oral or injectable PrEP. Adherence represents the percentage of prescribed doses that a person on PrEP takes on time. For oral PrEP, high adherence is taking PrEP at least 4 times/week; for injectable, it is getting the injection within 2 weeks of when it is due. The total number of people on PrEP at any time is determined by inputs that define initiation rates (Table 5.9), dropout rates from PrEP (Table 5.9), and infection rates among people on PrEP (Table 6.13 in Section 6.4) (Figure 5.1).

PrEP initiation rates may be affected by the COVID-19 pandemic during time periods 2 through 4 (Table 5.9). PrEP dropout rates are assumed to not be affected by the COVID-19 pandemic.

Among MSM and HETs, only those without HIV and with multiple HIV transmission risk factors are eligible for PrEP in HOPE. All PWID without HIV are eligible for PrEP. PrEP initiation by people without HIV in those subpopulations may occur in any year. Two types of inputs define initiation rates into the 4 PrEP states, overall PrEP initiation rates and 3 inputs for defining the distribution of those PrEP-initiators between oral and injectable PrEP and two adherence levels (low and high) for both oral and injectable PrEP. Note that the inputs defining the distribution between PrEP delivery mechanism are set so that 100% of PrEP users receive oral PrEP in all HOPE time periods. Distribution of PrEP users between oral and injectable PrEP may be varied by varying initiation rates to those compartments over time. People do not change adherence levels or switch between oral and injectable after PrEP initiation.

Figure 5.1	Inputs, Flows, and States Reflecting Oral and Injectable PrEP Use in HOPE

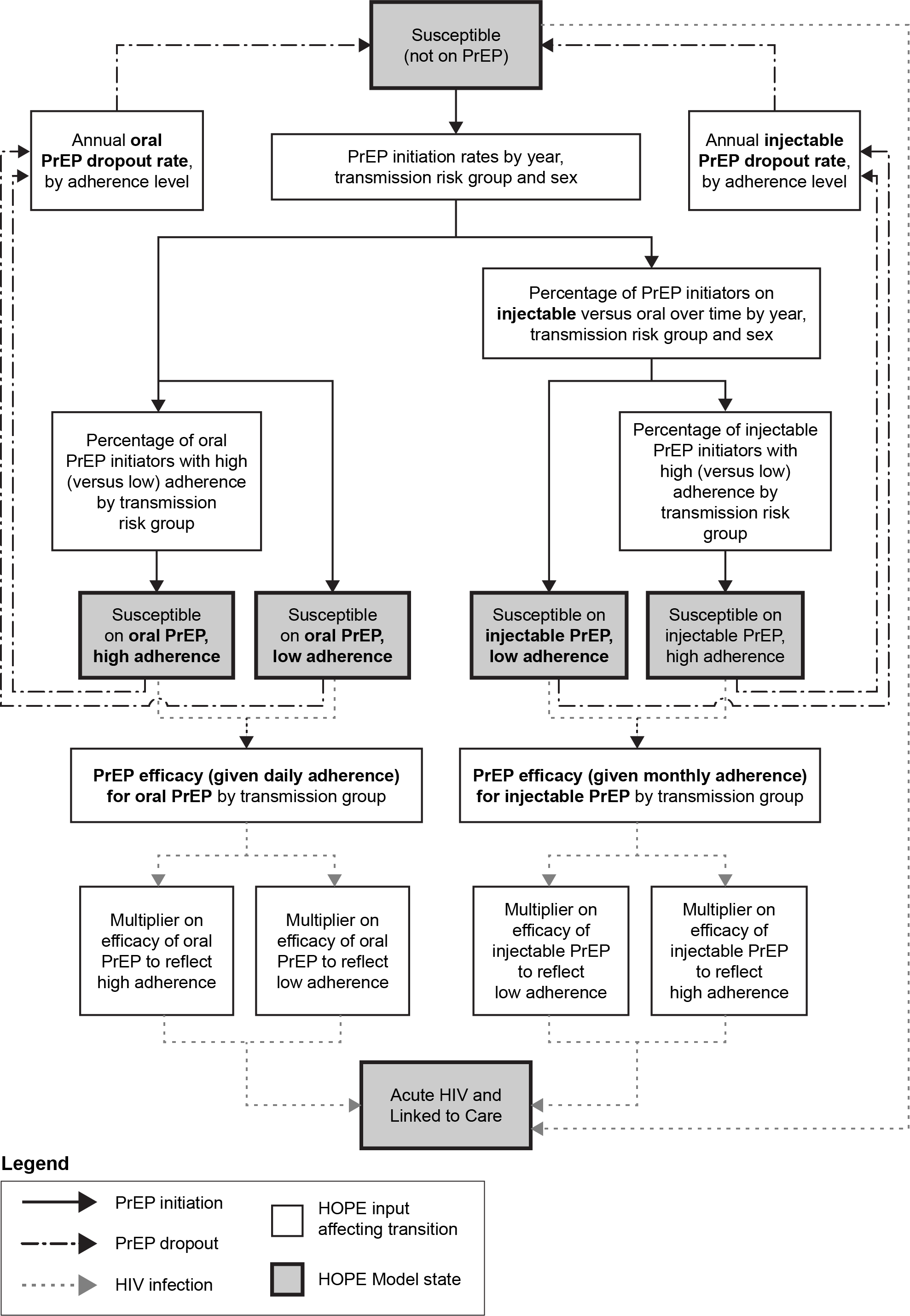

Persistence reflects how long a person stays on PrEP; it is reflected through drop-out rate inputs. Two inputs define drop-out rates from PrEP, one for oral PrEP and one for injectable PrEP. People that drop out of PrEP move back into the not-on-PrEP state. Drop-out rates are allowed to vary between oral and injectable PrEP and by adherence level (high or low).

All people who are on PrEP, regardless of type (oral/injectable) or adherence level, incur costs; however, these costs vary based on PrEP type and adherence level (further discussed in Section 1).

PrEP effectiveness also varies between oral and injectable PrEP and between adherence levels. People on PrEP have reduced incidence of HIV infection (further discussed in Section 6.4). People who become infected while on PrEP are assumed to be immediately diagnosed and linked to care, based on the assumption that those on PrEP are regularly tested and already engaged in medical care. The effect of PrEP on infection rates is a function of two sets of inputs, two inputs defining PrEP efficacy given perfect adherence and four inputs for multipliers on PrEP efficacy values to reflect effectiveness under actual adherence (Table 6.13).

Values to populate HOPE’s inputs and assumptions defining initiation, dropout, and effectiveness for oral and injectable PrEP were first obtained via targeted reviews of the published literature and available data. The identified values were also validated with CDC subject matter experts.

Table 5.9	Inputs defining PrEP participation in HOPE

| Input | | | Male HETs | | Female HETs | | MSM | | PWID | | Sources for values / assumptions |
| --- | --- | --- | --- | --- | --- | --- | --- | --- | --- | --- | --- |
| Annual PrEP initiation rate among eligible not already on PrEP^a^ | | |  | |  | |  | |  | |  |
| 2010-2011 | | 0.00000 | | 0.00000 | | 0.00000 | | 0.00000 | | Assumed no PrEP prior to 2012 | |
| 2012-2014 | | 0.00008 | | 0.00011 | | 0.00872 | | 0.00023 | | Estimated so that numbers on PrEP match surveillance estimates in 2014 (720 male HETs, 1,021 female HETs, 11,674 MSM, 319 PWID, per Huang et al., 2018) | |
| 2015-2019 | | 0.00143 | | 0.00137 | | 0.15416 | | 0.00610 | | Estimated so that numbers on PrEP match surveillance estimates in 2018 (13,223 male HETs, 13,769 female HETs, 191,450 MSM, 7,980 PWID, per CDC, 2021c) | |
| 2020-2021 | | 0.00229 | | 0.00219 | | 0.24665 | | 0.00976 | | Assumed same relative differences in initiation rates between male HETs, female HETs, MSM, and PWID as in 2015-2019 rates, but estimated relative increases versus those 2015-2019 rates so that numbers on PrEP approximated forecasted trends over years 2017 to 2020 (161,185; 218,614; 269,106; 279,130, respectively) from AHEAD data (USDHHS, 2022) and then assumed linear growth from 2020 to the 2030 AHEAD goal for number on PrEP (608,105) (USDHHS, 2022). | |
| Percent reduction in annual rate of initiating PrEP due to COVID-19, given eligible for PrEP^b^ | | | | | | | | | | | Reflects 25% drop in new PrEP enrollees over March 2020-March 2021 reported by Huang et al (2022a). Assumed no reductions in 2021 or 2022. |
| 2020 | | | -------------------------------25.0%----------------------------- | | | | | | | |  |
| 2021 | | | -------------------------------0.00%----------------------------- | | | | | | | |  |
| 2022 | | | -------------------------------0.00%----------------------------- | | | | | | | |  |
| Percentage of PrEP-initiators initiating injectable vs. oral | | | 0% | | 0% | | 0% | | 0% | | Assumed 0% on injectable PrEP for all HOPE time periods. |
| Percentage of oral PrEP initiators with hjgh (vs low) adherence | | | -------------------------------72.3%----------------------------- | | | | | | | | High adherence to oral PrEP is defined in HOPE to be at least 4 pills per week. For the MSM population, Landovitz et al., 2021 reported adherence among those with biomarkers consistent with intake of 4+ pills per week to be 72.3% among participants in the oral arm of the CAB-LA clinical trial. The percentage of those on PrEP in the PWID and HET transmission risk groups are assumed to be equal to that of the MSM risk group. |
| Percentage of injectable PrEP initiators with hjgh (vs low) adherence | | | -------------------------------55.0%----------------------------- | | | | | | | | High visit adherence to injectable PrEP is defined as injections having been received within a window of 2 weeks of the target injection date; low adherence reflects injections being received anytime beyond that window. We assumed 55% with high-adherence, the midpoint of the 50-60% range recommended by CDC subject matter experts on October 4, 2021. |
| Annual dropout rate from use of oral PrEP | | | ----------------------High adherence: 0.92----------------------  ----------------------Low adherence: 0.92---------------------- | | | | | | | | Reflects a 60% probability of dropping off PrEP per year, which is the approximate mean of Chan et al., 2019; Holloway et al., 2020; Coy et al., 2019; Wu et al., 2020; Huang et al., 2021 and Morgan et al, 2018. Due to lack of data, the same dropout rate is used for both the low and high adherence states. |
|  | Annual dropout rate from use of injectable PrEP^b^ | | ----------------------High adherence: 0.19----------------------  ----------------------Low adherence: 0.19---------------------- | | | | | | | | An annual dropout rate = 0.1943 was applied, equal to the dropout rate corresponding with 17.7% of participants dropping out from the injectable arm of the injectable PrEP clinical trial, as reported by Landovitz et al. (2021). Due to lack of data to support a differential, the same dropout rates were applied for the low and high adherence states. |

^a^ When inputs are stratified by transmission risk group and sex, only HETs are stratified between males and females but PWID are not stratified by sex due to the small size of that population

^b^ The model includes a setting to indicate whether to apply COVID effects on continuum of care, PrEP use, and risk behaviors in model calculations. The default setting is “no” so, by default, these effects are not applied.

**Figure 5.2.** IRR between the Black and H/L populations and other populations, considering scenarios with and without the COVID-19 effects. We see that the effect is minimal.

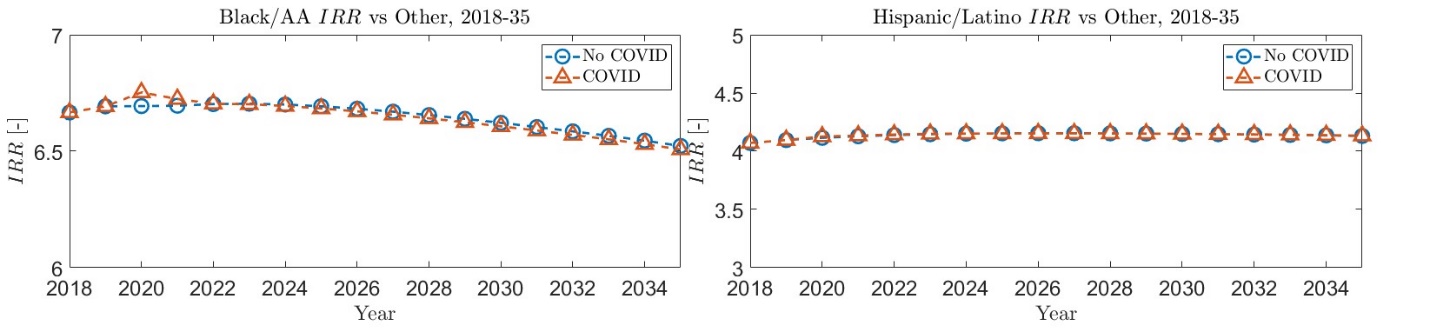

### 6. Force of Infection

This section outlines the methods applied for calculating the force of HIV infection, represented by lambda (λ). Infection via both vaginal and anal sex acts occurs for at least some individuals in all transmission groups. Infection via shared needles occurs for PWID only. Infection risk is calculated per person without HIV.

The force of infection for people without HIV who are not on PrEP (c = 1) or participating in a syringe services program (SSP; not distinguished by a separate compartment) is equal to λ. For people without HIV who are on PrEP or in an SSP, their force of infection is equal to λ reduced by a multiplicative reduction in infection risk; details on the application of those effects are outlined in Sections 6.4 and 6.5.

#### 6.1 Sexual and Needle-Sharing Partnerships

The partner pool of each transmission group was defined so that illogical partnerships, such as sexual partnerships between HET males and HET males and needle-sharing partnerships between PWID and HETs (as HETs do not, by definition, share needles), did not occur. To implement these assumptions, we created two mixing matrices to represent the distributions of sexual and needle-sharing partners of people without HIV, respectively, within any given year by transmission group, sex, number of HIV transmission risk factors, race/ethnicity, and age group.

The distribution of sexual partners was defined by sex and transmission group, race/ ethnicity, age group, and number of HIV transmission risk factors. Tables 6.1 through 6.5 outline the values used to determine the distribution of sexual partners by these categories; Tables 6.6 through 6.8 outline the values used to determine the distribution of needle-sharing partners. The values in several of these mixing matrices were calibrated to ensure that the annual number of new HIV infections (both overall and among key subpopulations) estimated by the model closely matched CDC surveillance data; details on that calibration are outlined in Section 10. We assumed random mixing within each partner pool.

Table 6.1.	Distribution of Sexual Partners by Sex and Transmission Group

| Partnering Populations | | | | | | |
| --- | --- | --- | --- | --- | --- | --- |
| Sex and Transmission Group | HET  Males | HET Females | MSM | PWID Males | PWID Females | Source |
| HET males |  | 96.7% |  |  | 3.3% | Determined by calibration |
| HET females | 97.6% |  | 2.1% | 0.3% |  | Determined by calibration |
| MSM |  | 13.1% | 86.6% |  | 0.2% | Determined by calibration |
| PWID male |  | 52.1% |  |  | 47.9% | Determined by calibration |
| PWID female | 59.5% |  | 1.0% | 39.5% |  | Determined by calibration; PWID females—MSM partnerships assumed |

Note: HET = heterosexual; MSM = men who have sex with men; PWID = people who inject drugs

Table 6.2.	Distribution of Sexual Partners by Race/Ethnicity, HET and PWID

| Race/Ethnicity | Black | Hispanic | White/ Other | Source |
| --- | --- | --- | --- | --- |
| Black | 89.5% | 2.7% | 7.8% | Determined by calibration |
| Hispanic | 6.8% | 66.2% | 27.0% |  |
| White/other | 3.5% | 4.9% | 91.6% |  |

Note: HET = heterosexual; MSM = men who have sex with men

Table 6.3.	Distribution of Sexual Partners by Race/Ethnicity, MSM

| Race/Ethnicity | Black | Hispanic | White/ Other | Source |
| --- | --- | --- | --- | --- |
| Black | 88.5% | 2.5% | 9.0% | Determined by calibration |
| Hispanic | 7.9% | 59.9% | 32.3% |  |
| White/other | 3.1% | 4.3% | 92.6% |  |

Note: MSM = men who have sex with men

Table 6.4.	Distribution of Sexual Partners by Age, HET and PWID

| Age Group | 13–17 | 18–24 | 25–34 | 35–44 | 45–54 | 55–64 | 65+ | Source |
| --- | --- | --- | --- | --- | --- | --- | --- | --- |
| 13–17 | 91.05% | 2.24% | 2.24% | 2.24% | 2.00% | 0.00% | 0.24% | Calculated from Glick et al. (2012) |
| 18–24 | 2.24% | 91.05% | 2.24% | 2.24% | 2.00% | 0.00% | 0.24% |  |
| 25–34 | 9.46% | 9.46% | 62.18% | 9.46% | 9.00% | 0.00% | 0.46% |  |
| 35–44 | 5.25% | 5.25% | 5.25% | 70.00% | 10.00% | 4.00% | 0.25% |  |
| 45–54 | 0.00% | 5.25% | 5.25% | 5.25% | 70.00% | 10.00% | 4.25% |  |
| 55–64 | 0.00% | 0.00% | 2.24% | 5.25% | 15.00% | 75.00% | 2.51% |  |
| 65+ | 0.00% | 0.00% | 0.00% | 2.00% | 7.00% | 10.00% | 81.00% |  |

Note: HET = heterosexual; MSM = men who have sex with men

Table 6.5.	Distribution of Sexual Partners by Age, MSM

| Age Group | 13–17 | 18–24 | 25–34 | 35–44 | 45–54 | 55–64 | 65+ | Source |
| --- | --- | --- | --- | --- | --- | --- | --- | --- |
| 13–17 | 33.0% | 43.0% | 20.0% | 2.6% | 0.5% | 0.5% | 0.5% | Determined by calibration. |
| 18–24 | 10.5% | 34.9% | 37.7% | 10.0% | 2.3% | 2.3% | 2.3% |  |
| 25–34 | 4.0% | 22.8% | 40.1% | 24.4% | 3.8% | 2.5% | 2.5% |  |
| 35–44 | 2.5% | 10.0% | 22.5% | 38.7% | 18.9% | 4.9% | 2.5% |  |
| 45–54 | 2.5% | 4.5% | 8.5% | 20.0% | 38.9% | 20.4% | 5.2% |  |
| 55–64 | 0.5% | 4.5% | 5.4% | 9.8% | 19.6% | 39.8% | 20.4% |  |
| 65+ | 0.5% | 0.5% | 2.4% | 11.8% | 14.2% | 24.1% | 46.5% |  |

Note: MSM = men who have sex with men

Table 6.5.	Distribution of Sexual Partners by Number of HIV Transmission Risk Factors, by Transmission Group

| Transmission Group | Number of HIV Transmission Risk Factors^a^ | Fewer | Multiple | Source |
| --- | --- | --- | --- | --- |
| HET | Fewer | 93.3% | 6.7% | Determined by calibration. Mixing for HETs with multiple HIV transmission risk factors and PWIDs are assumed to be equal. |
|  | Multiple | 22.8% | 77.2% |  |
| PWID^b^ | Multiple | 22.8% | 77.2% |  |
| MSM | Fewer | 82.3% | 17.7% |  |
|  | Multiple | 36.7% | 63.3% |  |

Note: HET = heterosexual; MSM = men who have sex with men; PWID = people who inject drugs

^a^ See Section 3 for more information on how numbers of HIV transmission risk factors are defined. ^b^ All PWID have multiple HIV transmission risk factors.

Table 6.6.	Distribution of Needle-Sharing Partners by Sex (PWID Only)

| Sex | Males | Females | Source |
| --- | --- | --- | --- |
| Males | 81% | 19% | Determined by calibration. |
| Females | 45% | 55% |  |

Note: PWID = people who inject drugs

Table 6.7.	Distribution of Needle-Sharing Partners by Race/Ethnicity (PWID Only)

| Race/Ethnicity | Black | Hispanic | White/ Other | Source |
| --- | --- | --- | --- | --- |
| Black | 80% | 5% | 15% | Data not available to inform this value. Assumed. |
| Hispanic | 5% | 80% | 15% |  |
| White/other | 15% | 5% | 80% |  |

Note: PWID = people who inject drugs

Table 6.8.	Distribution of Needle-Sharing Partners by Age Group (PWID Only)

| Age Group | 13–17 | 18–24 | 25–34 | 35–44 | 45–54 | 55–64 | 65+ | Source |
| --- | --- | --- | --- | --- | --- | --- | --- | --- |
| 13–17 | 91.05% | 2.24% | 2.24% | 2.24% | 2.00% | 0.00% | 0.24% | Assumed same as sexual mixing by age. |
| 18–24 | 2.24% | 91.05% | 2.24% | 2.24% | 2.00% | 0.00% | 0.24% |  |
| 25–34 | 9.46% | 9.46% | 62.18% | 9.46% | 9.00% | 0.00% | 0.46% |  |
| 35–44 | 5.25% | 5.25% | 5.25% | 79.00% | 10.00% | 4.00% | 0.25% |  |
| 45–54 | 0.00% | 5.25% | 5.25% | 5.25% | 70.00% | 10.00% | 4.25% |  |
| 55–64 | 0.00% | 0.00% | 2.24% | 5.25% | 15.00% | 75.00% | 2.51% |  |
| 65+ | 0.00% | 0.00% | 0.00% | 2.00% | 7.00% | 10.00% | 81.00% |  |

Note: PWID = people who inject drugs

#### 6.2 Per-Partnership Transmission Risk

We calculated the risk of infection per serodiscordant partnership for people without HIV according to a Bernoulli process that was a function of per-act sex- and shared needle–transmission risk estimates and the number of sex acts and shared needles per partnership. In this model, per-act transmission risk represents the probability of transmission per sex act or shared needle between an HIV-infected and HIV-uninfected person. This method implicitly assumes that sex acts and shared needles within a partnership are independent, each with the same likelihood of infection based on an average weighted by the likelihood of condom use (Pinkerton et al., 1998). Tables 6.9 through 6.11 list the values applied for all parameters that determined these per-partnership risks, as well as the sources from which those values were obtained.

The model considers that the annual number of sexual contacts may be affected by the COVID-19 pandemic during time periods 2 through 4 (Table 6.11). The model also considers effects of the pandemic on mortality rates (details provided in Section 5.1), rates of progression along the HIV care continuum (details in Section 5.4.1), and PrEP initiation (details in Section 5.5).

Table 6.9.	Per-Act HIV Transmission Risk and Reductions in Risk Due to Circumcision, Viral Load Suppression, Condom Use, and Awareness of HIV Infection

| Parameter | Value | Source |
| --- | --- | --- |
| Base probability of transmission per condomless sex act with uncircumcised (if male) People without HIV | | |
| Vaginal insertive | 0.0005 | Determined by calibration within bounds approximately equal to confidence intervals reported by Patel et al. (2014) (with anal receptive lower bound slightly expanded) |
| Vaginal receptive | 0.0005 |  |
| Anal insertive | 0.0007 |  |
| Anal receptive | 0.0111 |  |
| Relative risk of transmission per sex act by disease stage (versus overall average transmission probabilities by contact type)^a^ | | |
| Acute | 6.8333 | Wawer et al. (2005) |
| CD4 > 500 | 0.5833 |  |
| CD4 350–500 | 0.5833 |  |
| CD4 200–350 | 1.1667 |  |
| CD4 < 200 | 3.5833 |  |
| Base probability of transmission per shared needle | | |
|  | *0.0023* | Determined by calibration within 95% confidence interval (0.0010–0.0050) based on Zaric et al. (2000); Wall et al. (1991), Kaplan and Heimer (1992) |
| Reduction in HIV transmission per sex act due to condom use | | |
| MSM | 0.710 | Smith et al. (2015) |
| HET | 0.802 | Weller & Davis (2002) |
| Reduction in HIV transmission per sex act if HIV-uninfected partner is circumcised vs. uncircumcised | | |
| Vaginal insertive | 0.54 | Siegfried et al. (2009) |
| Male-male anal insertive | 0.00 | Assumption due to lack of evidence otherwise. |
| Male-female anal insertive | 0.00 | Assumption due to lack of evidence otherwise. |
| Reduction in HIV transmission per act if partner is VLS vs. not VLS | | |
| Shared needle | *0.7907* | Determined by calibration. |
| Sex act | 0.99 | Bavinton et al. (2018); Cohen et al. (2016); Rodger et al. (2016); Rodger et al. (2018) |

Note: ART = antiretroviral therapy; HET = heterosexual; HIV = human immunodeficiency virus; MSM = men who have sex with men; VLS = viral load suppressed

^a^ Early infection, the time after acute infection but before viral set point, when transmission risk per sex act or shared needle is elevated and testing is less sensitive, is not accounted for in the model.

Table 6.10.	Sexual Partners and Sex Acts

| Parameter | Female | | | Male | | | Source |
| --- | --- | --- | --- | --- | --- | --- | --- |
|  | Black | Hispanic | Other | Black | Hispanic | Other |  |
| Annual number of sex acts per partner for people without HIV, by number of HIV transmission risk factors | | | | | | | |
| HET |  |  |  |  |  |  |  |
| Multiple | ------------45.3----------- | | | ------------29.1----------- | | | Calculated^a, b^ |
| Fewer | ------------47.3----------- | | | ------------43.6----------- | | | Calculated^a, b^ |
| PWID |  |  |  |  |  |  |  |
| Multiple | 6.6 | 1.5 | 3.7 | 9.8 | 12.6 | 11.9 | Calculated^c^ |
| Annual number of sexual partners per HIV-uninfected person, by number of HIV transmission risk factors | | | | | | | |
| HET |  |  |  |  |  |  |  |
| Multiple | 6.6 | 4.9 | 4.0 | 12.1 | 8.2 | 5.8 | Calculated from Lin et al., (2016), using 2013 NHBS data and Leichliter et al., (2010)^b^ |
| Fewer | 0.8 | 0.7 | 0.7 | 1.2 | 0.9 | 0.7 | Calculated from Lin et al., (2016), using 2006-2008 NSFG data and Leichliter et al., (2010)^b^ |
| PWID |  |  |  |  |  |  |  |
| Multiple | 10.2 | 43.6 | 18.4 | 6.9 | 5.4 | 5.7 | CDC unpublished data based on 2009 NHBS IDU cycle 2 |
| MSM |  |  |  |  |  |  |  |
| Multiple | N/A^d^ | N/A^d^ | N/A^d^ | 8.3 | 7.0 | 8.0 | CDC unpublished data based on 2008 NHBS MSM cycle 2 |
| Fewer | N/A^d^ | N/A^d^ | N/A^d^ | 2.2 | 2.3 | 2.0 |  |
| Annual number of sex acts with all partners per HIV-uninfected person | | | | | | | |
| HET, by number of HIV transmission risk factors | | | | | | | |
| Multiple | ---------------------------205---------------------------- | | | | | | Calculated from Leichliter et al., (2010)^e^ |
| Fewer | ----------------------------34---------------------------- | | | | | |  |
| PWID | ----------------------------68---------------------------- | | | | | | Calculated from Reece et al. (2010a); Herbenick et al. (2010). Further detail on the calculation is available from the corresponding author. |
| MSM, overall |  |  |  |  |  |  |  |
| 13-17 | N/A^d^ | N/A^d^ | N/A^d^ | -------------30----------- | | | Ages 13–17: Herbenick et al. (2010)  Ages 18 and over: Wall et al. (2013) |
| 18-24 | N/A^d^ | N/A^d^ | N/A^d^ | -------------89------------ | | |  |
| 25-34 | N/A^d^ | N/A^d^ | N/A^d^ | -------------88------------ | | |  |
| 35-44 | N/A^d^ | N/A^d^ | N/A^d^ | -------------71------------ | | |  |
| 45-54 | N/A^d^ | N/A^d^ | N/A^d^ | -------------57------------ | | |  |
| 55-64 | N/A^d^ | N/A^d^ | N/A^d^ | -------------55------------ | | |  |
| 65+ | N/A^d^ | N/A^d^ | N/A^d^ | -------------46------------ | | |  |

(continued)

Table 6.10.	Sexual Partners and Sex Acts (continued)

|  | | Female | | | | | | Male | | | | Source | |
| --- | --- | --- | --- | --- | --- | --- | --- | --- | --- | --- | --- | --- | --- |
|  |  | Black | | Hispanic | | Other | | Black | Hispanic | Other | |  |  |
| Annual number of sex acts with all partners per HIV-uninfected person (continued) | | | | | | | | | | | | | |
| MSM with fewer HIV transmission risk factors | | | | | | | | | | | |  | |
| 13-17 | | N/A^d^ | | N/A^d^ | | N/A^d^ | | 29.0 | 27.5 | 28.4 | | Calculated^f^ | |
| 18-24 | | N/A^d^ | | N/A^d^ | | N/A^d^ | | 60.0 | 63.8 | 81.0 | |  |  |
| 25-34 | | N/A^d^ | | N/A^d^ | | N/A^d^ | | 69.4 | 66.8 | 74.0 | |  |  |
| 35-44 | | N/A^d^ | | N/A^d^ | | N/A^d^ | | 75.2 | 56.2 | 64.1 | |  |  |
| 45-54 | | N/A^d^ | | N/A^d^ | | N/A^d^ | | 45.6 | 51.3 | 48.2 | |  |  |
| 55-64 | | N/A^d^ | | N/A^d^ | | N/A^d^ | | 50.9 | 48.7 | 46.9 | |  |  |
| 65+ | | N/A^d^ | | N/A^d^ | | N/A^d^ | | 35.8 | 34.7 | 42.0 | |  |  |
| MSM with multiple HIV transmission risk factors | | | | | | | | | | | |  | |
| 13-17 | | N/A^d^ | | N/A^d^ | | N/A^d^ | | 36.6 | 40.4 | 31.5 | | Determined by calibration | |
| 18-24 | | N/A^d^ | | N/A^d^ | | N/A^d^ | | 110.3 | 110.7 | 91.8 | |  |  |
| 25-34 | | N/A^d^ | | N/A^d^ | | N/A^d^ | | 111.0 | 107.9 | 123.0 | |  |  |
| 35-44 | | N/A^d^ | | N/A^d^ | | N/A^d^ | | 79.3 | 81.8 | 82.3 | |  |  |
| 45-54 | | N/A^d^ | | N/A^d^ | | N/A^d^ | | 76.6 | 62.9 | 65.4 | |  |  |
| 55-64 | | N/A^d^ | | N/A^d^ | | N/A^d^ | | 56.6 | 66.3 | 71.4 | |  |  |
| 65+ | | N/A^d^ | | N/A^d^ | | N/A^d^ | | 50.0 | 60.3 | 63.5 | |  |  |
| Percentage of people without HIV’ vaginal sex acts with infected partners that are protected with a condom when partners are undiagnosed | | | | | | | | | | | | |  |
| HET/PWID | |  | |  | |  | |  |  | |  | |  |
| 13-17 | | 83.5% | | 68.6% | | 46.2% | | 83.5% | 68.6% | | 46.2% | | Reece et al. (2010b)^g^ |
| 18-24 | | 50.9% | | 41.8% | | 28.1% | | 50.9% | 41.8% | | 28.1% | |  |
| 25-34 | | 30.0% | | 24.7% | | 16.6% | | 30.0% | 24.7% | | 16.6% | |  |
| 35-44 | | 23.8% | | 19.6% | | 13.2% | | 23.8% | 19.6% | | 13.2% | |  |
| 45-54 | | 16.6% | | 13.6% | | 9.2% | | 16.6% | 13.6% | | 9.2% | |  |
| 55-64 | | 10.8% | | 8.9% | | 6.0% | | 10.8% | 8.9% | | 6.0% | |  |
| 65+ | | 6.8% | | 5.6% | | 3.8% | | 6.8% | 5.6% | | 3.8% | |  |
| MSM |  | | | | | |  | | | | | |  |
| 13–17 | N/A^d^ | | N/A^d^ | | N/A^d^ | | 96.2% | | 79.1% | | 53.3% | | Reece et al. (2010b); assumed to be same as male HETs |
| 18–24 | N/A^d^ | | N/A^d^ | | N/A^d^ | | 54.6% | | 44.9% | | 30.2% | |  |
| 25–34 | N/A^d^ | | N/A^d^ | | N/A^d^ | | 33.1% | | 27.2% | | 18.3% | |  |
| 35–44 | N/A^d^ | | N/A^d^ | | N/A^d^ | | 28.5% | | 23.5% | | 15.8% | |  |
| 45–54 | N/A^d^ | | N/A^d^ | | N/A^d^ | | 18.5% | | 15.2% | | 10.2% | |  |
| 55–64 | N/A^d^ | | N/A^d^ | | N/A^d^ | | 9.5% | | 7.9% | | 5.3% | |  |
| 65+ | N/A^d^ | | N/A^d^ | | N/A^d^ | | 6.8% | | 5.6% | | 3.8% | |  |

(continued)

Table 6.10.	Sexual Partners and Sex Acts (continued)

|  | | | Female | | | | | Male | | | | Source | |
| --- | --- | --- | --- | --- | --- | --- | --- | --- | --- | --- | --- | --- | --- |
|  |  |  | Black | | | Hispanic | Other | Black | | Hispanic | Other |  |  |
| Percentage of people without HIV’s anal sex acts with infected partners protected with a condom when partners are undiagnosed | | | | | | | | | | | |  | |
| With male partners | | |  | | | | |  | | | |  | |
| 13–17 | | | --------------13.2%------ | | | | | 56.1% | | 56.1% | 56.1% | Female-male partnerships: from Reece et al. (2010b).  Male-male partnerships: Determined in a previous calibration | |
| 18–24 | | |  |  |  |  |  | 56.1% | | 56.1% | 56.1% |  |  |
| 25–34 | | |  |  |  |  |  | 49.0% | | 49.0% | 49.0% |  |  |
| 35–44 | | |  |  |  |  |  | 45.5% | | 45.5% | 45.5% |  |  |
| 45–54 | | |  |  |  |  |  | 42.9% | | 42.9% | 42.9% |  |  |
| 55–64 | | |  |  |  |  |  | 41.3% | | 41.3% | 41.3% |  |  |
| 65+ | | |  |  |  |  |  | 40.3% | | 40.3% | 40.3% |  |  |
| With female partners | | | N/A | | | N/A | N/A | ---------------17.8%---------------- | | | | Calculated from Reece et al. (2010b)^h^ | |
| Percentage of uninfected MSM’s sex acts with other MSM that are insertive (vs. receptive), by age group | | | | | | | | | | | |  | |
| 13–17 | | | N/A^d^ | | | N/A^d^ | N/A^d^ | 42.7% | | 43.5% | 51.9% | Determined by calibration | |
| 18–24 | | | N/A^d^ | | | N/A^d^ | N/A^d^ | 30.4% | | 31.5% | 60.6% |  |  |
| 25–34 | | | N/A^d^ | | | N/A^d^ | N/A^d^ | 35.7% | | 35.2% | 69.1% |  |  |
| 35–44 | | | N/A^d^ | | | N/A^d^ | N/A^d^ | 41.2% | | 37.7% | 66.5% |  |  |
| 45–54 | | | N/A^d^ | | | N/A^d^ | N/A^d^ | 48.7% | | 48.3% | 64.7% |  |  |
| 55–64 | | | N/A^d^ | | | N/A^d^ | N/A^d^ | 50.3% | | 49.4% | 63.4% |  |  |
| 65+ | | | N/A^d^ | | | N/A^d^ | N/A^d^ | 49.9% | | 49.4% | 62.8% |  |  |
| If participating in AI, percentage of male–female sexual partnerships that include AI | | | | | | | | | | | | | |
| 13–17 | |  | | | ------------------------80.1%------------------------ | | | | | | | | Determined by calibration |
| 18–24 | |  | | | ------------------------80.3%------------------------ | | | | | | | |  |
| 25–34 | |  | | | ------------------------80.1%------------------------ | | | | | | | |  |
| 35–44 | |  | | | ------------------------80.0%------------------------ | | | | | | | |  |
| 45–54 | |  | | | ------------------------79.9%------------------------ | | | | | | | |  |
| 55–64 | |  | | | ------------------------79.8%------------------------ | | | | | | | |  |
| 65+ |  | | | ------------------------80.0%------------------------ | | | | | | | | |  |
| Percentage of Sexual Acts That Are Anal (vs. Vaginal) in Male-Female Partnerships with Anal Intercourse | | | | | | | | | | | | | |
| 13–17 |  | | | 14.3% | | | | | 31.6% | | | | Female: NSSHB data from Reece et al. (2010a)  Male: NSSHB data from Herbenick et al. (2010) |
| 18–24 |  | | | 19.0% | | | | | 24.6% | | | |  |
| 25–34 |  | | | 20.6% | | | | | 21.9% | | | |  |
| 35–44 |  | | | 16.6% | | | | | 20.5% | | | |  |
| 45–54 |  | | | 17.2% | | | | | 20.5% | | | |  |
| 55–64 |  | | | 23.0% | | | | | 25.1% | | | |  |
| 65+ |  | | | 36.2% | | | | | 21.0% | | | |  |

Table 6.10.	Sexual Partners and Sex Acts (continued)

|  | | | Female | | | | Male | | | | | | Source | |
| --- | --- | --- | --- | --- | --- | --- | --- | --- | --- | --- | --- | --- | --- | --- |
|  |  |  | Black | | Hispanic | Other | Black | | Hispanic | | Other | |  |  |
| Percentage of People Who Have Anal Intercourse in their Male-Female Sexual Partnerships^i^ | | | | | | | | | | | | | | |
| 13–17 |  | 2.7% | | 5.6% | | 3.7% | | 3.0% | | 5.3% | | 2.5% | | Calculated from Herbenick et al. (2010), Reece et al. (2010a), Dodge et al. (2010), and Finlayson et al. (2011) |
| 18–24 |  | 14.3% | | 29.5% | | 20.1% | | 6.5% | | 11.6% | | 5.4% | |  |
| 25–34 |  | 14.4% | | 28.8% | | 24.1% | | 23.3% | | 28.6% | | 25.3% | |  |
| 35–44 |  | 12.2% | | 21.0% | | 18.2% | | 15.8% | | 27.0% | | 22.8% | |  |
| 45–54 |  | 8.4% | | 12.3% | | 9.0% | | 12.5% | | 18.8% | | 16.0% | |  |
| 55–64 |  | 6.0% | | 7.6% | | 4.4% | | 8.0% | | 13.8% | | 6.5% | |  |
| 65+ |  | 9.3% | | 7.1% | | 1.7% | | 1.0% | | 10.0% | | 2.0% | |  |

Note: HET = heterosexual; HIV = human immunodeficiency virus; MSM = men who have sex with men; NHBS = National HIV Behavioral Surveillance; NSSHB = National Survey of Sexual Health and Behavior; PWID = people who inject drugs

^a^ Annual number of sex acts per partner were calculated as (annual number of sex acts with all partners if HIV-uninfected or HIV-infected and undiagnosed) ÷ (annual number of sexual partners). Because the denominator (annual number of partners) was stratified by race and the numerator (annual number of sexual acts) was not, we used data from the same source as the denominator (Leichliter, et a.l, 2010) to calculate overall non-race-specific estimates of annual number of partners. Those non-race-specific estimates were then used in the calculations.

^b^ Lin et al., 2016 reported 1.14 partners for all HETs (calculated from 2006-2008 NSFG data published in Chandra et al., 2011) and 5.73 for HETs with multiple HIV transmission risk factors (unpublished 2013 NHBS data). Those were combined with race- and sex-specific ratios from Leichliter et al. (2010) to back out race- and sex- specific estimates of number of partners for HETs with fewer and multiple HIV transmission risk factors, respectively.

^c^ Calculated as (annual number of sex acts with all partners if HIV-uninfected or HIV-infected and undiagnosed) ÷ (annual number of sexual partners).

^d^ Values are not reported for MSM in the “Female” column since MSM, by definition, only include males.

^e^ Used Leichliter et al., 2010 Table 1 values for the distribution of vaginal sex acts in the past 4 weeks by sex and race/ethnicity to calculate the annual number of total acts per year. Assumed that the top 20% represented people with multiple HIV transmission risk factors and bottom 80% people with fewer HIV transmission risk factors.

^f^ Calculated based on the overall average annual contacts per year for MSM, the annual number of sexual contacts for MSM with multiple HIV transmission risk factors (which is calibrated), the percentage of MSM with multiple HIV transmission risk factors, and MSM population sizes by race/ethnicity.

^g^ Race-specific and age-specific values from Reece et al. (2010b) are based on the percentage of past 10 vaginal intercourse events condom protected. White/other values based on "White" population.

^h^ Reece et al. (2010b) report the percentage of condom use for all males and MSM. The HET male-specific percentages were derived by taking the all-male condom use as a weighted average of HET and MSM condom use.

^i^ It is assumed that all male-female sexual partnerships include vaginal intercourse.

Table 6.11.	Other Risk Behaviors and Inputs that Determine Transmission Risk

| Risk Behavior | Value | Source |
| --- | --- | --- |
| Percentage of PWID actively injecting | 40% | Assumed^a^ |
| Annual number of injections per year per active PWID | 455 | Calculated from CDC unpublished data from the 2015 NHBS IDU cycle^b^ |
| Percentage of injections that are shared | *11.9%* | Determined by calibration |
| Annual number of needles shared across all partners | 11.3 | Calculated^c^ |
| Annual number of needle-sharing partners for PWID | 9.5 | Assumption that number of needle-sharing partners for PWID is equal to the number of sexual partners for PWID as estimated from CDC unpublished data from the 2009 NHBS IDU cycle 2 |
| Number of needles shared per partner | *1.2* | Calculated^d^ |
| Reduction in sexual partners for diagnosed PWH (vs. undiagnosed or uninfected) | HET: 10.2%  MSM: 6.8%  PWID: 10.2% | Unpublished analysis of 2009-2015 NHBS^e^ |
| Increase in percentage of people without HIV' sex acts with partners with HIV that are protected with a condom when partner is diagnosed vs. undiagnosed | 112% | Malekinejad et al. (2021); Reece et al. (2010b)^f^ |
| Percent reduction in annual number of sexual contacts per high-risk uninfected person due to COVID-19^g^ |  | Unpublished analysis of iSTAMP data^g^ |
| Time period 2 (2020) | 20.0% |  |
| Time period 3 (2021) | 10.0% |  |
| Time period 4 (2022) | 0.0% |  |
| Percentage of sex acts in which condom provides effective protection | | |
| VI | 80.2% | Weller and Davis (2002) |
| AI | 70% | Smith et al. (2015) |
| Reduction in number of needles shared with partners with HIV who are diagnosed versus undiagnosed or HIV-uninfected | 27% | Assumption based on approximately half of condom use effect from diagnosis on needle-sharing behaviors |

Note: AI = anal intercourse; NHBS = National HIV Behavioral Surveillance; PWH = people with HIV; PWID = people who inject drugs; VI = vaginal intercourse

^a^ Assumed that only 40% of PWID are actively using in a given year, based on an estimated ~600,000 PWID in the United States who have injected within the last 12 months derived from Lanksy et al. (2014), divided by an estimated ~1.6 million total PWID in the United States who have injected in their lifetime, per Tempalski et al. (2013).

Table 6.11.	Other Risk Behaviors (continued)

^b^ Calculated using data on the injection frequency in the past 12 months for injection drug users from the 2015 NHBS IDU cycle, which shows that among 10,487 PWIDs, 7,742 inject more than once a day, 1,131 once a day, 950 more than once a week, 659 once a week or less, and 5 had no response. Assuming those categories corresponded to 1.5 injections per day, 1 per day, 2 per week, 0.75 per week, and 0, respectively, the weighted average number of injections per person per year was 455.43.

^c^ Calculated as (Annual number of injections across all partners per year) x (Percentage of injections that are shared)

^d^ Calculated as (Number of needles shared across all partners) ÷ (Annual number of needle-sharing partners for PWID) 

^e^ Calculated from unpublished NHBS 2009-2015 data on the percentage of survey respondents living with HIV who were sexually active while aware of HIV infection versus the percentage that were sexually active while unaware of HIV infection.

^f^ 20% condom use during sex acts for HIV-uninfected and undiagnosed HIV-infected was based on population-wide estimates from Reece et al. (2010b). The relative risk of not always using condoms 12 months after diagnosis (Malekinejad et al., 2021) was transformed to relative risk reduction (RRR) by deducting them from 1, and multiplied by 100 to express as % reduction in risk of adverse outcome.

^g^ Unpublished iSTAMP analysis (Viguerie et al., 2023) that found a reduction in number of sexual partners by about 20% among Black and Latino MSM with multiple HIV transmission risk factors. Assumed half of the 2020 reduction in 2021, and no reduction in 2022. The model includes a setting to indicate whether to apply COVID effects on continuum of care, PrEP use, and risk behaviors in model calculations. The default setting is “no” so, by default, these effects are not applied.

##### 6.2.1 Per-Sex-Act Sexual and Needle Transmission Probabilities

The probability of an HIV-uninfected person acquiring HIV from a sex act with a partner with HIV varies by the disease stage and continuum-of-care status of the partner; circumcision status of the HIV-uninfected person; condom usage; transmission group; and type of sex act (i.e., vaginal vs. anal and insertive vs. receptive) (Boily et al., 2009; Leynaert et al., 1998; Osmond et al., 1988; Porco et al., 2004). The probabilities were each calculated as the product of a base probability for an HIV-infected person having condomless sex act with an uncircumcised (if male) HIV-uninfected partner and the relative risk of transmission by disease stage (base probabilities and relative risks listed in Table 6.9). Multiplicative reductions were then applied to those probabilities for sex acts involving circumcised HIV-uninfected partners, sex acts with the use of condoms and/or other prevention tools, and sex acts with partners with HIV who are VLS.

The base probabilities of transmission per condomless sex act with uncircumcised (if male) people without HIV were calibrated within bounds approximately equal to those reported in Patel et al. (2014). We applied the same sources and methods as were applied in Sorensen et al. (2012) to estimate the relative risk of transmission by disease stage. Sorensen and colleagues used clinical trial data, citing Wawer et al. (2005), on transmission risk from heterosexual partnerships by disease stage of the partner with HIV and overall per-act sexual risk to calculate transmission risk by disease stage.

The probability of transmission from a shared needle was calibrated and was assumed in the base-case not to vary by disease stage of the partner with HIV. For needles shared with a partner with HIV with VLS, a multiplicative reduction to that probability was applied.

##### 6.2.2 Number of Sex Acts and Needles Shared per Partner

The number of sex acts per partner was calculated as the annual number of sex acts with all partners divided by the annual number of sexual partners. We assumed no reduction in the number of sex acts across all partners for HIV-infected, diagnosed individuals versus people without HIV and HIV-infected, undiagnosed individuals. The number of sexual partners varied by transmission group, sex, number of HIV transmission risk factors, and race/ethnicity (see Table 6.10).

The number of needles shared per partner was calculated as the annual number of injections across all partners per year multiplied by the percentage of injections that are shared divided by the annual number of needle-sharing partners.

##### 6.2.3 Calculation of Per-Partnership Transmission Risk

Per-partnership transmission risk is represented by $\beta_{p1,p2}^{z,y,c}$ (beta), which is the probability of transmission for an individual without HIV in subpopulation *p1* per sexual or needle-sharing partnership from transmission risk type *z* (vaginal, anal, or needle) in a partnership type *y* (male-female partnership with vaginal intercourse only, male-male partnership with anal intercourse only, male-female partnership that includes both vaginal and anal intercourse, or needle-sharing) with a partner who is in subpopulation *p2* and compartment *c*. If the partner is uninfected, the risk is zero. The values of the betas for sexual and needle-sharing partnerships are calculated by using Equations (6.1) and (6.2), respectively.

Equation (6.1) is complex but has a simple structure:

1 − [(Probability of not getting infected by condomless receptive sex acts)*
(Probability of not getting infected by receptive sex acts with the use of condoms)*
(Probability of not getting infected by condomless insertive sex acts)*
(Probability of not getting infected by insertive sex acts with the use of condoms)].

$\beta_{p1,p2,t}^{z,y,c}$ =$\begin{matrix} 1-\left( 1-\left[ 1-\Delta_{c,z} \right]\left[ \Phi_{c,z} \right] \right)^{\left[ S_{p1} \right][1-\ddot{C}_{t}^{s=7}]\left[ 1-G_{z,p1,c} \right]\left[ V_{p1,p2} \right]\left[ \Omega_{p1,p2,z,y} \right]} \\ \left( 1-\left[ 1-d_{z} \right]\left[ 1-\Delta_{c,z} \right]\left[ \Phi_{c, z} \right] \right)^{\left[ S_{p1} \right][1-\ddot{C}_{t}^{s=7}]\left[ G_{z,p1,c} \right]\left[ V_{p1,p2} \right]\left[ \Omega_{p1,p2,z,y} \right]} \\ \left( 1-\left[ 1-b_{p1,p2,z} \right]\left[ 1-\Delta_{c,z} \right]\left[ \Gamma_{c,z} \right] \right)^{\left[ S_{p1} \right][1-\ddot{C}_{t}^{s=7}]\left[ 1-G_{z,p1,c} \right]\left[ 1-V_{p1,p2} \right]\left[ \Omega_{p1,p2,z,y} \right]} \\ \left( 1-\left[ 1-d_{z} \right]\left[ 1-b_{p1,p2,z} \right]\left[ 1-\Delta_{c,z} \right]\left[ \Gamma_{c,z} \right] \right)^{\left[ S_{p1} \right][1-\ddot{C}_{t}^{s=7}]\left[ G_{z,p1,c} \right]\left[ 1-V_{p1,p2} \right]\left[ \Omega_{p1,p2,z,y} \right]} \end{matrix}$

for z = {vaginal intercourse and anal intercourse}, *y* = {male-female partnerships with vaginal intercourse only, male-male partnership with anal intercourse only, male-female partnership with both vaginal and anal intercourse}, and p1 and p2 = {all subpopulations}	(6.1)

$\beta_{p1,p2,t}^{z,y,c}$ = $1-\left( 1-\left[ 1-\Delta_{c,z} \right]\left[ \Theta_{c} \right] \right)^{(E){(1-T}_{p1,p2,c})}$

for	*z* = {needle-sharing}, *y* = {needle-sharing}, and *p1* and
*p2* = {PWID subpopulations}	(6.2)

where

- S_p_ = number of annual sex acts for an individual without HIV per partner, HIV-uninfected or undiagnosed, by subpopulation *p*;
- G_z,p,c_ = percentage of sex acts of risk type z protected with a condom, given partner in compartment *c*, by subpopulation *p*;
- $\Delta_{c,z}$= percentage reduction in per-act transmission probability due to VLS, by compartment c and transmission risk type z;
- $\Phi_{c,z}$ and $\Gamma_{c,z}$= per-sex-act transmission probability for condomless receptive and insertive acts, respectively, of type z (vaginal or anal intercourse) with partner in compartment *c*; calculated as a product of *base probability of transmission per condomless sex act* and *relative risk of transmission per sex act by disease stage*, the latter of which varies by *c*;
- d_z_ = percentage reduction in per-sex-act transmission probability from an act of type z due to condom use;
- b_p1,p2,z_ = percentage reduction in per-insertive sex act transmission probability from an act of type z due to circumcision for an individual without HIV in subpopulation *p1* with a partner with HIV in subpopulation *p2* (0 if subpopulation *p1* is uncircumcised male or female);
- V_p1,p2_ = proportion of male-male sex acts by individuals in subpopulation *p1* with individuals in subpopulation *p2* that are receptive (0 if subpopulation *p1* or *p2* is not MSM);
- Ω_p1,p2,z,y_= proportion of sexual acts by individuals in subpopulation *p1* in partnerships of type *y* with individuals in subpopulation *p2* that are risk type *z* (where *z* = vaginal or anal);
- E = number of needles shared annually per needle-sharing partner by PWID who has never been diagnosed with HIV;
- Τ_p1,p2,c_ = reduction in number of needles shared between PWID populations *p1* and *p2* for diagnosed versus undiagnosed or uninfected (0 if compartment *c* is for undiagnosed or uninfected compartments); and
- Θ_c_ = probability of HIV transmission per needle shared with a partner with HIV in compartment *c*; calculated as a product of *base probability of transmission per shared needle* and *relative risk of transmission per needled shared by disease stage*, the latter of which varies by *c*.

#### 6.3 Calculation of Force of Infection for Individuals Not on PrEP and Not Participating in SSP

The force of infection λ is defined as the rate of infection per uninfected person across all sources. The method that we applied to calculate force of infection is based on the method applied in Long et al. (2010) but adapted to include multiple transmission groups, and mixing is determined explicitly by inputs defining percentage of partners by race/ethnicity, transmission groups, number of HIV transmission risk factors, and age groups. Long et al. was based on the heterosexual population only and assumed proportional mixing, in which “persons with many sexual partners are more likely to select a partner who similarly has many partners” (p. 779).

The force of HIV infection is a function of the number of vaginal intercourse, anal intercourse, and needle-sharing partners, per-partnership transmission risk (described in Section 6.2), prevalence of HIV among partners, and the distribution of partners with HIV among the different disease and care-continuum stages.

The force of HIV infection for people without HIV of any given subpopulation *p* is calculated as the sum of the forces of infection from five different sources of infection, four sexual forces of infection and one needle-sharing force of infection (listed and further described in Table 6.12), as stated in Equation (6.3):

${\lambda_{p}\left( t \right)=\lambda}_{p}^{z=1,x=1}\left( t \right)+\lambda_{p}^{z=2,x=2}\left( t \right)+\lambda_{p}^{z=1,x=3}\left( t \right)+\lambda_{p}^{z=2,x=3}\left( t \right)+\lambda_{p}^{z=3}(t)$	(6.3)

Table 6.12.	Five Sources of Infection that Contribute to Overall Force of Infection for Each Subpopulation

| Source of Infection | Relevant Subpopulations (*p*) and Sexual Transmission Participation Type (*x*) | Risk Type (*z*) | Betas ($\boldsymbol{\beta}_{\boldsymbol{p,p}\boldsymbol{2}}^{\boldsymbol{z,y,c}}$) Used in Calculation of Lambda |
| --- | --- | --- | --- |
| VI in people who only participate in VI in their male-female partnerships | p = {All}, x = VI only in male-female partnerships (1) | VI (z=1) | Beta for z = VI, y = male-female partnership that only includes VI |
| AI from male-male partnerships | p = {MSM}, x = AI only in male-male partnerships (2) | AI (z=2) | Beta for z = AI, y = male-male partnership that only includes AI |
| VI in people who participate in AI in their male-female partnerships | p = {All}, x = AI and VI in male-female partnerships (3) | VI (z=1) | Beta for z = VI, y = male-female partnership that includes AI;  Beta for z = VI, y = male-female partnership that only includes VI |
| AI in people who participate in AI in their male-female partnerships | p = {All}, x = AI and VI in male-female partnerships (3) | AI (z=2) | Beta for z = AI, y = male-female partnership that includes AI |
| Needle-sharing | p ={PWID}, x = N/A | Needle-sharing (z=3) | Beta for z = needle-sharing, y = needle-sharing |

Note: AI = anal intercourse; HET = heterosexual; MSM = men who have sex with men; PWID = people who inject drugs; VI = vaginal intercourse

Equation (6.4) then describes the calculation of the force of infection from risk type *z* for people without HIV in subpopulation *p* who participate in sexual transmission risk behaviors of each type x, at time *t*. Table 6.12 describes the relevant z, x, p, and y values applied in Equation (6.4).

$\lambda_{p}^{z,x}\left( t \right)=-\ln\left( 1-\left( \varrho_{p,x} \right)\sum_{y} \left( 1-\prod_{p2} \left[ \prod_{c} \left[ \left[ 1-\beta_{p1,p2,t}^{z,y,c} \right]^{\xi_{z,y,p,p2,c} (t)} \right] \right] \right) \right)$,	(6.4)

where

- $\varrho_{p,x}$ $\varrho_{x}$ $\varrho_{x}$ = Proportion of subpopulation p who participate in transmission risk participation type *x*, which is calculated based on the following:
  - the input *Percentage of people who have anal intercourse (AI) in their male-female partnerships*
  - the assumption that 100% of MSM have AI in their male-male partnerships, and
  - the assumption that 100% of all transmission groups have vaginal intercourse (VI) in their male-female partnerships.
- ξ _z,y,p,p2,c_ (t) = Number of partnerships of type y involving risk type z per individual without HIV in subpopulation p with infected partners in subpopulation p2 in compartment c at time t, which is calculated using the method outlined in Equation (6.5):

$$\xi_{z,y,p,p2,c}(t)=\left( Number of partnership type y partners in p2 per person in p \right)*\left( Percentage of individuals in p2 that are in infected and in c at time t \right)$$

$=(M_{p}^{z})(a_{p,p2})(O_{p,y})\left( \frac{X_{p2}^{c}\left( t \right)*s_{c}}{\sum_{c*} X_{p2}^{c*}(t)} \right)$ $=(m_{p}^{z})(Percentage of p^{'}s partners in p2)\left( \frac{X_{p2}^{c}}{\sum_{c=1to27} X_{p2}^{c}} \right)\left( \eta_{p,c}^{s} \right)$ $=(m_{p}^{z})(Percentageofp^{'}spartnersinp2)\left( \frac{X_{p2}^{c}}{\sum_{c=1to27} X_{p2}^{c}} \right)\left( \eta_{p,c}^{s} \right)$	(6.5)

where

- $M_{p}^{z}$ $m_{p}^{z}$ $m_{p}^{z}$ = annual number of partners for risk type z per person in p,
- $a_{p,p2}$= percentage of individuals in subpopulation p’s partners that are in p2, as determined by the mixing matrix described in Section 6.1,
- $O_{p,y}$= average percentage of partnerships for an individual in subpopulation p that are type y, given that the individual has partnerships of type y. It is calculated based on the following:
  - the input *Among people who have AI in their male-female partnerships, percentage of those partnerships with AI*, which distributes partnerships for people who have AI in male-female partnerships into partnerships with and without AI
  - the assumption that risk from male-female partnerships does not factor into the calculation of force of infection from male-male sexual partnerships with AI only (z = 2, x = 2)
  - the assumption that risk from male-male partnerships does not factor into the calculation of force of infection from VI in male-female partnerships with VI only (z = 1, x = 1) or from either VI or AI in male-female partnerships with both AI and VI (z = 1 or 2, x = 3).
- $s_{c}$ = binary indicator that model state c is an infected HIV state (1 = infected, 0 = not infected)
- Number of partnership type y partners in p2 per person in p =$(M_{p})(a_{p,p2})(O_{p,y})$.

This value ($\xi_{z,y,p,p2,c}$) equals 0 for uninfected compartments c = {1, 2, 3, 4 or 5}.

This method captures the following effects on HIV transmission:

- the impact of circumcision on the per-partnership transmission risk (beta)
- the impact of viral suppression on beta
- the impact of vaginal versus anal sexual risk in all transmission groups on beta
- the impact of the prevalence of HIV in the partner populations on the per-person force of infection (lambda)
- the impact of mixing patterns on lambda

This method has the following simplifying assumption:

- Sufficient partner supply always exists to support the distribution of partners specified by the inputs.

#### 6.4 Calculation of Force of Infection for Individuals on PrEP

Infection risk for people without HIV on PrEP (*c* = 2, 3, 4, 5) is calculated as the infection risk for people without HIV not on PrEP ($\lambda_{p}\left( t \right)$) reduced by a multiplicative factor. Those multiplicative factors are each calculated as the product of efficacy given full adherence (daily for oral or monthly for injectable) and a multiplier reflecting less-than-full adherence; those efficacy and multiplier inputs for oral and injectable PrEP are listed in Table 6.13.

Table 6.13.	Inputs for Defining Reduction in Infection Risk if on PrEP

| Input | Default value(s) | Sources for values / assumptions |
| --- | --- | --- |
| Efficacy of oral PrEP given daily adherence | HET: 99%  MSM: 99%  PWID: 84% | Daily adherence is defined 7 doses per week. The efficacy values are taken the CDC recommended values for defining 'Effectiveness of prevention strategies to reduce the risk of acquiring or transmitting HIV' (CDC, 2019c). |
| Efficacy of injectable PrEP given monthly adherence | HET: 99%  MSM: 99%  PWID: 84% | Full adherence is defined as getting injections on target injection date. HPTN083 and HPTN084 clinical trial results reported by Landovitz et al. (2021) and HIV Prevention Trials Network (2020) suggest almost 100% effectiveness of PrEP for MSM and HETs, respectively, and superiority of effectiveness for injectable versus oral PrEP. No trial results are reported for PWID. Since oral PrEP efficacy is already assumed to be almost 100% (especially for MSM and HETs), the conservative assumption was applied to use the same efficacy for injectable as for oral PrEP. |
| Multiplier on oral PrEP efficacy to reflect less-than-full adherence | High adherence: 0.99  Low adherence: 0.55 | Multipliers, when applied to efficacy input value of 0.99, are between the effectiveness estimates for 4+ doses per week and <4 doses per week, respectively, from Grant et al. (2014) (1.0 / 0.488) and Anderson et al (2012) (0.983 / 0.591); both analyses used Landovitz et al. (2021) study findings |

Table 6.13.	Inputs for Defining Reduction in Infection Risk if on PrEP (continued)

| Input | Default value(s) | Sources for values / assumptions |
| --- | --- | --- |
| Multiplier on injectable PrEP efficacy to reflect less-than-full adherence | High adherence: 0.99  Low adherence: 0.99 | Effectiveness measures by adherence are not reported in the clinical trials (Landovitz et al., 2021; HIV Prevention Trials Network, 2020). We applied 0.99 for both high- and low-adherence because, per CDC subject matter expert guidance provided on October 4, 2021, effectiveness does not wane with moderately increased intervals between doses. |

Note: HET = heterosexual; MSM = men who have sex with men; PWID = people who inject drugs

We assumed that individuals on PrEP are also already engaged in the health care system and tested for HIV regularly and, therefore, when infected with HIV, immediately diagnosed and linked to HIV care.

#### 6.5 Calculation of Force of Infection for Individuals in SSP

PWID may participate in a SSP in any year. Infection risk for people without HIV participating in a SSP is calculated as the infection risk for people without HIV not participating in a SSP reduced by a multiplicative factor (Table 6.14). We apply the multiplicative factor through a weighted reduction to needle-sharing infection risk for PWID without HIV so that the effect is only on the portion of PWID who are participating in SSPs.

Table 6.14.	Participation and Infection Risk Reduction for SSP

| Input | Value | Source |
| --- | --- | --- |
| Number of PWID without HIV served by SSPs annually |  |  |
| 2013-2014 |  | Des Jarlais et al. (2015); Handanagic et al. (2021)^a^ |
| Black | 6,513 |  |
| Hispanic/Latino | 6,528 |  |
| Other | 13,428 |  |
| 2015 |  | Des Jarlais et al. (2020); Adams et al. (2019); Handanagic et al. (2021)^b^ |
| Black | 35,109 |  |
| Hispanic/Latino | 35,189 |  |
| Other | 72,388 |  |
| 2016-2017 |  |  |
| Black | 53,662 |  |
| Hispanic/Latino | 53,784 |  |
| Other | 110,641 |  |
| 2018+ |  | Handanagic et al. (2021)^c^ |
| Black | 78,368 |  |
| Hispanic/Latino | 78,547 |  |
| Other | 161,582 |  |
| Percentage reduction in the annual rate of HIV transmission if PWID at risk for HIV is using SSP | 34% | Aspinal et al. (2014)^d^ |

Note: HET = heterosexual; MSM = men who have sex with men; PWID = people who inject drugs; SSP = syringe exchange program
^a^ Total number of PWID without HIV served by SSP calculated based on $21,920,618 total U.S. spending (2013$, $25,497,538 in 2021$) on syringe services in 2013 per Table 1 in Des Jarlais et al. (2015) divided by the cost per PWID served by SSP ($774.00 in 2016$ $873.85 in 2021$) per PWID participating in an SSP (per Teshale et al., 2019). Value then scaled by proportion of PWID who are HIV-negative, based on calculations for HOPE model initiation population. Number served by race/ethnicity derived from the distribution of HIV-negative PWID served by SSP in Handanagic et al. (2021).
^b^ Total number of PWID served by SSP calculated from total number of sterile syringes purchased by SSPs participating in the The Buyer’s Club (42,200,000 in 2015; 64,500,000 in 2016), the proportion of SSPs represented in The Buyer’s Club (Des Jarlais et al., 2020), the percentage of PWID who are HIV-negative, and the number of needles per user (number of needles per active PWID multiplied by the proportion of needles per HIV-negative SSP user obtained from SSPs (Adams et al., 2019 & Handanaric et al., 2021)). Number served by race/ethnicity derived from the distribution of HIV-negative PWID served by SSP in Handanagic et al. (2021).

^c^ Total number of PWID served by SSP calculated from Handanagic et al. (2021), which reported that 55% of HIV-negative PWID were receiving sterile syringes from SSPs in 2018. Number served by race/ethnicity derived from the distribution of HIV-negative PWID served by SSP in Handanagic et al. (2021).
^d^ A relative HIV incidence of 0.66 (95% CI: 0.43, 1.01) was reported for PWID participating in a syringe exchange program in Aspinall et al. (2014).

### 7. Differential Equations that Define the Model

This section outlines the differential equations that define this model. The equations are organized into subsections by compartments *c* based on disease stage *h*. The differential equations in the model are solved using Euler’s method with a time-step equal to 0.1 year. Table 7.1 lists the model’s 30 compartments.

Table 7.1.	Model Compartments

| Number | Description | Row-Column Designation for Each Compartment |
| --- | --- | --- |
| 1 | People without HIV / not on PrEP | A1 |
| 2 | People without HIV / on oral PrEP/ high adherence | A6 |
| 3 | People without HIV / on oral PrEP/ low adherence | A7 |
| 4 | People without HIV / on injectable PrEP/ high adherence | A8 |
| 5 | People without HIV / on injectable PrEP/ low adherence | A9 |
| 6 | PWH / acute stage / unaware of infection | B1 |
| 7 | PWH / acute stage / aware, but not linked to HIV care | B2 |
| 8 | PWH / acute stage / linked to HIV care, but not on ART | B3 |
| 9 | PWH / CD4>500 / unaware of infection | C1 |
| 10 | PWH / CD4>500 / aware, but not linked to HIV care | C2 |
| 11 | PWH / CD4>500 / linked to HIV care, but not on ART | C3 |
| 12 | PWH / CD4>500 / on ART, but not virally suppressed | C4 |
| 13 | PWH / CD4>500 / virally suppressed, which assumes persons are in care and on ART | C5 |
| 14 | PWH / CD4 350–500 / unaware of infection | D1 |
| 15 | PWH / CD4 350–500 / aware, but not linked to HIV care | D2 |
| 16 | PWH / CD4 350–500 / linked to HIV care, but not on ART | D3 |
| 17 | PWH / CD4 350–500 / on ART, but not virally suppressed | D4 |
| 18 | PWH / CD4 350–500 / virally suppressed, which assumes persons are in care and on ART | D5 |
| 19 | PWH / CD4 200–350 / unaware of infection | E1 |
| 20 | PWH / CD4 200–350 / aware, but not linked to HIV care | E2 |
| 21 | PWH / CD4 200–350 / linked to HIV care, but not on ART | E3 |
| 22 | PWH / CD4 200–350 / on ART, but not virally suppressed | E4 |
| 23 | PWH / CD4 200–350 / virally suppressed, which assumes persons are in care and on ART | E5 |
| 24 | PWH / CD4 < 200 / unaware of infection | F1 |
| 25 | PWH / CD4 < 200 / aware, but not linked to HIV care | F2 |
| 26 | PWH / CD4 < 200 / linked to HIV care, but not on ART | F3 |
| 27 | PWH / CD4 < 200 / on ART, but not virally suppressed | F4 |

(continued)

Table 7.1.	Model Compartments (continued)

| Number | Description | Row-Column Designation for Each Compartment |
| --- | --- | --- |
| 28 | PWH / CD4 < 200 / virally suppressed, which assumes persons are in care and on ART | F5 |
| 29 | Death among persons with CD4≥200 and susceptible people | N/A |
| 30 | Death among persons with CD4<200 | N/A |

Note: ART = antiretroviral therapy; PrEP = pre-exposure prophylaxis; N/A = not applicable

#### 7.1 Number of People without HIV

The numbers of people without HIV not on PrEP (*c =* 1) and on PrEP (c = 2, 3, 4, 5) within each subpopulation *p* are determined by Equations (7.1) through (7.5), respectively. For individuals not on PrEP, the number of people without HIV increases by aging into the observed population and people without HIV on PrEP stopping PrEP and decreases due to PrEP initiation. The number of people without HIV on PrEP increases due to initiation of PrEP and decreases due to stopping PrEP. For all susceptible compartments (c = 1 to 5), the number of people without HIV decreases by HIV infection or death. For all susceptible compartments, like all other main compartments, aging also shifts individuals between age groups.

$\frac{{dX}_{p}^{c=1}(t)}{dt}=\left[ \Lambda_{p}*f_{p} \right]\sum_{c=1}^{25} X_{p}^{c}(t=0)+\left[ Y_{a*=1}^{N=1} \right]X_{p}^{c=2}\left( t \right)+\left[ Y_{a*=2}^{N=1} \right]X_{p}^{c=3}\left( t \right)+\left[ Y_{a*=1}^{N=2} \right]X_{p}^{c=4}\left( t \right)+\left[ Y_{a*=2}^{N=2} \right]X_{p}^{c=5}\left( t \right)-\left[ \Psi_{p}\left( t \right)(1-\ddot{C}_{t}^{s=6})+\lambda_{p}\left( t \right)+(1+\ddot{C}_{t}^{s=8})\mu_{p}+\delta_{p}^{-} \right]X_{p}^{c=1}\left( t \right)+\left[ \delta_{p}^{+} \right]X_{p-1}^{c=1}(t)$	(7.1)

$\frac{{dX}_{p}^{c=2}(t)}{dt}=\left[ \Psi_{p}\left( t \right)* \alpha_{p}\left( t \right)* \Upsilon_{p}^{N=1}*(1-\ddot{C}_{t}^{s=6}) \right]X_{p}^{c=1}\left( t \right)-\left[ Y_{a*=1}^{N=1}+\left( 1-i_{p}* \Xi_{a*=1}^{N=1} \right)\lambda_{p}\left( t \right)+{(1+\ddot{C}_{t}^{s=8})\mu}_{p}+\delta_{p}^{-} \right]X_{p}^{c=2}\left( t \right)+ \left[ \delta_{p}^{+} \right]X_{p-1}^{c=2}(t)$	(7.2)

$\frac{{dX}_{p}^{c=3}(t)}{dt}=\left[ \Psi_{p}\left( t \right) * \alpha_{p}\left( t \right)* {(1- \Upsilon}_{p}^{N=1})(1-\ddot{C}_{t}^{s=6}) \right]X_{p}^{c=1}\left( t \right)-\left[ Y_{a*=2}^{N=1}+\left( 1-i_{p}* \Xi_{a*=2}^{N=1} \right)\lambda_{p}\left( t \right)+(1+\ddot{C}_{t}^{s=8})\mu_{p}+\delta_{p}^{-} \right]X_{p}^{c=3}\left( t \right)+\left[ \delta_{p}^{+} \right]X_{p-1}^{c=3}(t)$	(7.3)

$\frac{{dX}_{p}^{c=4}(t)}{dt}=\left[ \Psi_{p}\left( t \right) *\left( 1- \alpha_{p}\left( t \right) \right)* \Upsilon_{p}^{N=2}*(1-\ddot{C}_{t}^{s=6}) \right]X_{p}^{c=1}\left( t \right)-\left[ Y_{a*=1}^{N=2}+\left( 1-i_{p}* \Xi_{a*=1}^{N=2} \right)\lambda_{p}\left( t \right)+(1+\ddot{C}_{t}^{s=8})\mu_{p}+ \delta_{p}^{-} \right]X_{p}^{c=4}\left( t \right)+\left[ \delta_{p}^{+} \right]X_{p-1}^{c=4}(t)$	(7.4)

$\frac{{dX}_{p}^{c=5}(t)}{dt}=\left[ \Psi_{p}\left( t \right) *(1- \alpha_{p}\left( t \right))* {(1- \Upsilon}_{p}^{N=2})(1-\ddot{C}_{t}^{s=6}) \right]X_{p}^{c=1}\left( t \right)-\left[ Y_{a*=2}^{N=2}+\left( 1-i_{p}* \Xi_{a*=2}^{N=2} \right)\lambda_{p}\left( t \right)+{(1+\ddot{C}_{t}^{s=8})\mu}_{p}+\delta_{p}^{-} \right]X_{p}^{c=5}\left( t \right)+\left[ \delta_{p}^{+} \right]X_{p-1}^{c=5}(t)$	(7.5)

where

- $\chi_{p}^{c}(t)$ = number of individuals in the population in compartment *c* and demographic subpopulation *p* at time *t*;
- $\Lambda_{p}$ = constant rate of aging into the youngest age group in the modeled population per person (based on the size and distribution of that youngest group at *t* = 0) in subpopulation *p*;
- $f_{p}$ = relative adjustment to rate of aging into the youngest age group of the model for all *p* in the youngest age group (j = 1 if the youngest age group is 13–17 and j = 2 if youngest age group is 18–24).
- $Y_{a*}^{N}$ = annual probability of stopping PrEP if susceptible and on PrEP for PrEP type *N* and PrEP adherence level *a**;
- $\Psi_{p}(t)$ = annual probability of initiating PrEP (regardless of PrEP delivery mechanism), given eligible, for subpopulation *p* at time *t*;
- $\alpha_{p}(t)$ = percentage of PrEP initiators on oral (versus injectable) PrEP [*N* = 1], by subpopulation *p* at time *t*;
- $\Upsilon_{p}^{N}$ = percentage of PrEP initiators with high (versus low) adherence [*a** = 1], by subpopulation *p* and PrEP type *N*;
- *i*_p_ = percentage reduction in the annual rate of HIV transmission if individual without HIV in subpopulation *p* is on PrEP;
- $\Xi_{a*}^{N}$ = multiplier on efficacy of PrEP to account for less than full (daily for oral PrEP [*N* = 1] or monthly for injectable PrEP [*N* = 2]) adherence, by PrEP type *N* and adherence level *a**;
- $\mu_{p}$ = mortality rate among people without HIV in subpopulation *p*;
- $\delta_{p}^{+}$ = aging rates into (+) subpopulation *p* for all *p* in age groups older than the youngest age group included in the population (*j* = 2, 3, 4, 5, 6, 7 if youngest age group is 13–17 and *j* = 3, 4, 5, 6, 7 if youngest age group is 18–24); and
- $\delta_{p}^{-}$ = aging rates out of subpopulation *p* for all p in age groups younger than the oldest age group (j = 1, 2, 3, 4, 5, 6 if youngest age group is 13–17 and *j* = 2, 3, 4, 5, 6 if youngest age group is 18–24).

#### 7.2 Individuals with Acute HIV Infection

The numbers of individuals with acute HIV infections (*c =* 6, 7, 8) within each subpopulation *p* are determined by Equations (7.6) through (7.8), corresponding to continuum-of-care stages (*r*) 1 to 3, respectively. The numbers of individuals calculated in these equations vary in the factors that increase or decrease their values based on their continuum status; hence, the equations vary in the same way. Transitions that increase the values include HIV infection, diagnosis, and linkage to or departure from HIV care. Transitions that decrease the values include HIV progression, testing and notification of results, linkage to HIV care, ART initiation, and death. Aging also shifts individuals among age groups.

$\frac{{dX}_{p}^{c=6}\left( t \right)}{dt}=\left[ \lambda_{p}\left( t \right) \right]X_{p}^{c=1}\left( t \right)-\left( \omega_{p}^{c=6}\left( t \right)+\tau_{p}^{c=6}\left( t \right)+{(\sigma}_{p}^{c=6})(1+\ddot{C}_{t}^{s=8})+\delta_{p}^{-} \right)X_{p}^{c=6}\left( t \right)+\left[ \delta_{p}^{+} \right]X_{p-1}^{c=6}(t)$	(7.6)

$\frac{{dX}_{p}^{c=7}\left( t \right)}{dt}=\left[ \tau_{p}^{c=6}\left( t \right) \right]\left[ 1-\kappa_{p}\left( t \right) \right]X_{p}^{c=6}\left( t \right)+\left[ e_{p}\left( t \right) \right]X_{p}^{c=8}\left( t \right)-$	$\left[ \mathcal{l}_{p}^{c=7}\left( t \right)+\omega_{p}^{c=7}\left( t \right)+\left( \sigma_{p}^{c=7} \right)(1+\ddot{C}_{t}^{s=8})+\delta_{p}^{-} \right]X_{p}^{c=7}\left( t \right)+\left[ \delta_{p}^{+} \right]X_{p-1}^{c=7}(t)$	(7.7)

$$\frac{{dX}_{p}^{c=8}\left( t \right)}{dt}=\left[ \left( 1-i_{p}* \Xi_{a*=1}^{N=1} \right)\lambda_{p}\left( t \right) \right]X_{p}^{c=2}+\left[ \left( 1-i_{p}* \Xi_{a*=2}^{N=1} \right)\lambda_{p}\left( t \right) \right]X_{p}^{c=3}+\left[ \left( 1-i_{p}* \Xi_{a*=1}^{N=2} \right)\lambda_{p}\left( t \right) \right]X_{p}^{c=4}+\left[ \left( 1-i_{p}* \Xi_{a*=2}^{N=2} \right)\lambda_{p}\left( t \right) \right]X_{p}^{c=5}+\left[ \tau_{p}^{c=6}\left( t \right) \right]\left[ \kappa_{p}\left( t \right) \right]X_{p}^{c=6}\left( t \right)+\left[ \mathcal{l}_{p}^{c=7}\left( t \right) \right]X_{p}^{c=7}\left( t \right)-$$

$\left[ e_{p}\left( t \right)+[\gamma_{p}^{c=8}\left( t \right)][1-\ddot{C}_{t}^{s=3}]+\omega_{p}^{c=8}\left( t \right)+(\sigma_{p}^{c=8})(1+\ddot{C}_{t}^{s=8})+\delta_{p}^{-} \right]X_{p}^{c=8}\left( t \right)+{\left[ \delta_{p}^{+} \right]X}_{p-1}^{c=8}(t)$	(7.8)

where

- $\omega_{p}^{c}$ = rate of HIV progression from compartment *c* to the next disease stage if not on ART for subpopulation p;
- $\tau_{p}^{c}(t)$ = diagnosis rate from unaware compartment *c* to aware for subpopulation *p* at time *t*, for *c* = {6, 9, 14, 19, 24};
- $\sigma_{p}^{c}$ = mortality rate for PWH, by compartment *c* and subpopulation *p* at time *t*;
- $\gamma_{p}^{c=8}\left( t \right)$ = rate of ART initiation if linked to HIV care, by compartment c and subpopulation *p* at time *t*, for *c* = {11, 16, 21, 26};
- $e_{p}(t)$ = rate of departure from HIV care if linked to HIV care, by demographic subpopulation *p* at time *t*, for *c* = {8, 11, 16, 21, 26};
- $\kappa_{p}\left( t \right)$ = percentage of newly diagnosed individuals in subpopulation *p* who immediately link to care at diagnosis at time *t*; and
- $\mathcal{l}_{p}^{c=7}\left( t \right)$ = rate of linkage to HIV care among aware (not newly diagnosed) individuals in compartment *c* for subpopulation *p* at time *t*, for *c* = {7, 10, 15, 20, 25}.

#### 7.3 Individuals with Chronic HIV Infection and CD4 ≥ 200

The numbers of individuals with chronic HIV infection and CD4 ≥ 200 (*c =* 9 to 23), by demographic subpopulation *p*, are determined by Equations (7.9) through (7.18), corresponding to continuum-of-care stages (*r*) 1 to 5, respectively. Across these compartments, transitions that can increase the number of individuals in a particular compartment include HIV progression, testing and notification of results, linkage to or departure from HIV care, ART initiation (resulting in VLS or not), loss of ART, and loss of viral suppression. Transitions that can decrease the number of people with HIV (PWH) in any of these compartments include HIV progression, testing and notification of results, linkage to or departure from HIV care, ART initiation (resulting in VLS or not), loss of ART, loss of viral suppression, and death. Aging also shifts individuals among age groups.

*Equations for CD4 > 500 (h = 2):*

$\frac{{dX}_{p}^{c=9}(t)}{dt}=\left[ \omega_{p}^{c=6} \right]X_{p}^{c=6}\left( t \right)-\left[ \omega_{p}^{c=9}+\tau_{p}^{c=9}\left( t \right)+(\sigma_{p}^{c=9})(1+\ddot{C}_{t}^{s=8})+\delta_{p}^{-} \right]X_{p}^{c=9}\left( t \right)+\left[ \delta_{p}^{+} \right]X_{p-1}^{c=9}(t)$	(7.9)

$$\frac{{dX}_{p}^{c=10}\left( t \right)}{dt}=\left[ \omega_{p}^{c=7}\left( t \right) \right]X_{p}^{c=7}\left( t \right)+\left[ \tau_{p}^{c=9}\left( t \right) \right]\left[ 1-\kappa_{p}\left( t \right) \right]X_{p}^{c=9}\left( t \right)$$

$+\left[ e_{p}\left( t \right) \right]X_{p}^{c=11}\left( t \right)-\left[ \mathcal{l}_{p}^{c=10}\left( t \right)+\omega_{p}^{c=10}\left( t \right)+(\sigma_{p}^{c=10})(1+\ddot{C}_{t}^{s=8})+\delta_{p}^{-} \right]X_{p}^{c=10}\left( t \right)+\left[ \delta_{p}^{+} \right]X_{p-1}^{c=10}\left( t \right)$	(7.10)

$$\frac{{dX}_{p}^{c=11}\left( t \right)}{dt}=\omega_{p}^{c=8}X_{p}^{c=8}\left( t \right)+\tau_{p}^{c=9}\left( t \right)\kappa_{p}\left( t \right)X_{p}^{c=9}\left( t \right)+\mathcal{l}_{p}^{c=10}\left( t \right)X_{p}^{c=10}\left( t \right)$$

$+\left[ \varepsilon_{p}\left( t \right) \right]{[\ddot{1+C}_{t}^{s=4}]X}_{p}^{c=12}\left( t \right)-\left[ e_{p}\left( t \right)+{[\gamma}_{p}^{c=11}\left( t \right)][1-\ddot{C}_{t}^{s=3}]+\omega_{p}^{c=11}+{(\sigma}_{p}^{c=11})(1+\ddot{C}_{t}^{s=8})+\delta_{p}^{-} \right]X_{p}^{c=11}\left( t \right)+\left[ \delta_{p}^{+} \right]X_{p-1}^{c=11}(t)$	(7.11)

$$\frac{{dX}_{p}^{c=12}\left( t \right)}{dt}=\left[ 1-u \right]\left[ \gamma_{p}^{c=8}\left( t \right) \right]{[1-\ddot{C}_{t}^{s=3}]X}_{p}^{c=8}\left( t \right)+\left[ 1-u \right]\left[ \gamma_{p}^{c=11}\left( t \right) \right]X_{p}^{c=11}\left( t \right)+\left[ \eta_{p}\left( t \right) \right]{[\ddot{1+C}_{t}^{s=5}]X}_{p}^{c=13}\left( t \right)$$

$-\left[ \left( \varepsilon_{p}\left( t \right) \right)(\ddot{1+C}_{t}^{s=4})+\left( \zeta_{p}\left( t \right) \right)+H_{p}^{c=12}+{(\sigma}_{p}^{c=12}\left( t \right))(1+\ddot{C}_{t}^{s=8})+\delta_{p}^{-} \right]X_{p}^{c=12}\left( t \right)+\left[ \delta_{p}^{+} \right]X_{p-1}^{c=12}(t)$	(7.12)

$\frac{{dX}_{p}^{c=13}\left( t \right)}{dt}=\left[ u \right]\left[ \gamma_{p}^{c=8}\left( t \right) \right]X_{p}^{c=8}\left( t \right)+\left[ u \right]\left[ \gamma_{p}^{c=11}\left( t \right) \right]{[1-\ddot{C}_{t}^{s=3}]X}_{p}^{c=11}\left( t \right)$	$+\left[ \left( \zeta_{p}\left( t \right) \right) \right]X_{p}^{c=12}\left( t \right)+\left[ \vartheta_{p}^{c=18} \right]X_{p}^{c=18}\left( t \right)$	$-\left[ B^{c=13}+{(\eta}_{p}\left( t \right))\ddot{(1+C}_{t}^{s=5})+(\sigma_{p}^{c=13}\left( t \right))(1+\ddot{C}_{t}^{s=8})+\delta_{p}^{-} \right]X_{p}^{c=13}\left( t \right)+\left[ \delta_{p}^{+} \right]X_{p-1}^{c=13}(t)$	(7.13)

*Equations for CD4 200–500 (h =3 or 4):*

$\frac{{dX}_{p}^{c}(t)}{dt}=\left[ \omega_{p}^{c-5} \right]X_{p}^{c-5}\left( t \right)-\left[ \omega_{p}^{c}+\tau_{p}^{c}\left( t \right)+{(\sigma}_{p}^{c}(t))(1+\ddot{C}_{t}^{s=8})+\delta_{p}^{-} \right]X_{p}^{c}\left( t \right)+\left[ \delta_{p}^{+} \right]X_{p-1}^{c}(t)$
for c = {14, 19}	(7.14)

$$\frac{{dX}_{p}^{c}\left( t \right)}{dt}=\left[ \omega_{p}^{c-5} \right]X_{p}^{c-5}\left( t \right)+\left[ \tau_{p}^{c-1}\left( t \right) \right]\left[ 1-\kappa_{p}\left( t \right) \right]X_{p}^{c-1}\left( t \right)+\left[ e_{p}\left( t \right) \right]X_{p}^{c+1}\left( t \right)$$

$-\left[ \mathcal{l}_{p}^{c}\left( t \right)+\omega_{p}^{c}+(\sigma_{p}^{c}\left( t \right))(1+\ddot{C}_{t}^{s=8})+\delta_{p}^{-} \right]X_{p}^{c}\left( t \right)+\left[ \delta_{p}^{+} \right]X_{p-1}^{c}(t)$
for c = {15, 20}	(7.15)

$\frac{{dX}_{p}^{c}\left( t \right)}{dt}=\left[ \omega_{p}^{c-5} \right]X_{p}^{c-5}\left( t \right)+\left[ \tau_{p}^{c-2}\left( t \right) \right]\left[ \kappa_{p}\left( t \right) \right]X_{p}^{c-2}\left( t \right)+\left[ \mathcal{l}_{p}^{c-1}\left( t \right) \right]X_{p}^{c-1}\left( t \right)+\left[ \varepsilon_{p}\left( t \right) \right][\ddot{C}_{t}^{s=4}]X_{p}^{c+1}\left( t \right)$

$$-\left[ e_{p}\left( t \right)+(\gamma_{p}^{c}\left( t \right))(1-\ddot{C}_{t}^{s=3})+\omega_{p}^{c}+{(\sigma}_{p}^{c}(t))(1+\ddot{C}_{t}^{s=8})+\delta_{p}^{-} \right]X_{p}^{c}\left( t \right)+\left[ \delta_{p}^{+} \right]X_{p-1}^{c}\left( t \right)$$

for c = {16, 21}	(7.16)

$\frac{{dX}_{p}^{c}\left( t \right)}{dt}=\left[ {H_{p}^{c-5}} \right]X_{p}^{c-5}\left( t \right)+\left[ 1-u \right][\gamma_{p}^{c-1}\left( t \right)]{[1-\ddot{C}_{t}^{s=3}]X}_{p}^{c-1}\left( t \right)+\left[ \eta_{p}\left( t \right) \right]{[1+\ddot{C}_{t}^{s=5}]X}_{p}^{c+1}\left( t \right)$	$-\left[ \zeta_{p}\left( t \right)+{(\varepsilon}_{p}^{c})(\ddot{1+C}_{t}^{s=4})+H_{p}^{c}+(\sigma_{p}^{c}\left( t \right))(1+\ddot{C}_{t}^{s=8})+\delta_{p}^{-} \right]X_{p}^{c}\left( t \right)+\left[ \delta_{p}^{+} \right]X_{p-1}^{c}(t)$
for c = {17, 22}	(7.17)

$$\frac{{dX}_{p}^{c}\left( t \right)}{dt}=\left[ B^{c-5} \right]X_{p}^{c-5}\left( t \right)+\left[ u \right]\left[ \gamma_{p}^{c-2}\left( t \right) \right][1-\ddot{C}_{t}^{s=3}]X_{p}^{c-2}\left( t \right)+\left[ \zeta_{p}\left( t \right) \right]X_{p}^{c-1}\left( t \right)+\left[ \vartheta_{p}^{c+5} \right]X_{p}^{c+5}\left( t \right)$$

$$-\left[ B^{c}+(\eta_{p}\left( t \right))\ddot{(1+C}_{t}^{s=5})+\vartheta_{p}^{c}+{(\sigma}_{p}^{c}\left( t \right))(1+\ddot{C}_{t}^{s=8})+\delta_{p}^{-} \right]X_{p}^{c}\left( t \right)+\left[ \delta_{p}^{+} \right]X_{p-1}^{c}(t)$$

for c = {18, 23}	(7.18)

where

- $\varepsilon_{p}\left( t \right)$ = annual rate of dropping off of ART if ART-not-VLS, by demographic subpopulation *p* at time *t;*
- $\zeta_{p}\left( t \right)$= annual rate of becoming VLS if ART-not-VLS, by demographic subpopulation *p* at time *t;*
- $H_{p}^{c}$ = rate of HIV progression to the next disease stage from compartment *c,* if on ART-not-VLS, by demographic subpopulation p;
- $u$ = percentage of individuals who become VLS among those who initiate ART;
- $\eta_{p}\left( t \right)$ = rate of loss of viral load suppression if VLS, by demographic subpopulation *p* at time;
- $B^{c}$ = rate of HIV progression to the next disease stage from compartment *c,* if VLS; and
- $\vartheta_{p}^{c}$= rate of HIV regression to the previous disease stage from compartment *c*, if VLS, by demographic subpopulation p.

#### 7.4 Individuals with Chronic HIV Infection and CD4<200

The numbers of individuals with Chronic HIV Infection and CD4<200 (*c* = 24 to 28), by demographic subpopulation *p*, are determined by Equations (7.19) through (7.23), corresponding to continuum-of-care stages (*r*) 1 to 5, respectively. Across these compartments, transitions that can increase the number of individuals in a particular compartment include HIV progression, testing and notification of results, linkage to or departure from HIV care, ART initiation (resulting in VLS or not), loss of ART, and loss of viral suppression. Transitions that decrease the number of PWH in any of these compartments include testing and notification of results, linkage to and departure from HIV care, and death among persons with CD4<200. Aging also shifts individuals among age groups. These equations differ from the other sets of equations in that mortality is determined by death rates specifically for persons with HIV with CD4<200.

$\frac{{dX}_{p}^{c=24}\left( t \right)}{dt}=\omega_{p}^{c=19}X_{p}^{c=19}\left( t \right)-[\tau_{p}^{c=24}\left( t \right)+{(\sigma}_{p}^{c=24}\left( t \right))(1+\ddot{C}_{t}^{s=8})+\delta_{p}^{-}{]X}_{p}^{c=24}\left( t \right)+{[\delta}_{p}^{+}]X_{p-1}^{c=24}\left( t \right)$	(7.19)

$$\frac{{dX}_{p}^{c=25}\left( t \right)}{dt}=\left[ \tau_{p}^{c=24}\left( t \right) \right]\left[ 1-\kappa_{p}\left( t \right) \right]X_{p}^{c=24}\left( t \right)+\left[ \omega_{p}^{c=20} \right]X_{p}^{c=20}\left( t \right)$$

$+\left[ e_{p}\left( t \right) \right]X_{p}^{c=26}\left( t \right)-\left( \mathcal{l}_{p}^{c=25}\left( t \right)+\left[ \sigma_{p}^{c=25}\left( t \right) \right][1+\ddot{C}_{t}^{s=8}]+\delta_{p}^{-} \right)X_{p}^{c=25}\left( t \right)+\left[ \delta_{p}^{+} \right]X_{p-1}^{c=25}(t)$	(7.20)

$$\frac{{dX}_{p}^{c=26}\left( t \right)}{dt}=\left[ \tau_{p}^{c=24} \right] \left[ \kappa_{p}\left( t \right) \right]X_{p}^{c=24}\left( t \right)+\left[ \mathcal{l}_{p}^{c=25}\left( t \right) \right]X_{p}^{c=25}\left( t \right)+\left[ \omega_{p}^{c=21} \right]X_{p}^{c=21}\left( t \right)+\left[ \varepsilon_{p}\left( t \right) \right][1+\ddot{C}_{t}^{s=4}]X_{p}^{c=27}\left( t \right)$$

$-[{(\gamma}_{p}^{c=26}\left( t \right))(1-\ddot{C}_{t}^{s=3})+e_{p}\left( t \right)+(\sigma_{p}^{c=26}\left( t \right))(1+\ddot{C}_{t}^{s=8})+\delta_{p}^{-}]X_{p}^{c=26}\left( t \right)+\delta_{p}^{+}X_{p-1}^{c=26}(t)$	(7.21)

$\frac{{dX}_{p}^{c=27}\left( t \right)}{dt}=\left[ H_{p}^{c=22} \right]X_{p}^{c=22}\left( t \right)+\left[ 1-u \right]\left[ \gamma_{p}^{c=26}\left( t \right) \right]{[1-\ddot{C}_{t}^{s=3}]X}_{p}^{c=26}\left( t \right)+\left[ \eta_{p}\left( t \right) \right][1+\ddot{C}_{t}^{s=5}]X_{p}^{c=28}\left( t \right)$	$-\left[ (\varepsilon_{p}\left( t \right))(1+\ddot{C}_{t}^{s=4})+\zeta_{p}^{c=27}+{(\sigma}_{p}^{c=27}\left( t \right))(1+\ddot{C}_{t}^{s=8})+\delta_{p}^{-} \right]X_{p}^{c=27}\left( t \right)+\left[ \delta_{p}^{+} \right]X_{p-1}^{c=27}\left( t \right)$	(7.22)

$$\frac{{dX}_{p}^{c=28}\left( t \right)}{dt}=\left[ B^{c=23} \right]X_{p}^{c=23}\left( t \right)+\left[ {u][\gamma}_{p}^{c=26}\left( t \right) \right][1-\ddot{C}_{t}^{s=3}]X_{p}^{c=26}\left( t \right)+\left[ \zeta_{p}\left( t \right) \right]X_{p}^{c=27}\left( t \right)$$

$-\left[ {(\eta}_{p}\left( t \right))(\ddot{1+C}_{t}^{s=5})+\vartheta_{p}^{c=28}+(\sigma_{p}^{c=28}\left( t \right))(1+\ddot{C}_{t}^{s=8})+\delta_{p}^{-} \right]X_{p}^{c=28}\left( t \right)+\left[ \delta_{p}^{+} \right]X_{p-1}^{c=28}(t)$	(7.23)

#### 7.5 Absorbing States

The model’s absorbing states (*c* = 29 to 30) are compartments that hold individuals who have stopped being actively followed because they have died. The number of individuals by subpopulation *p* in the absorbing states are determined by Equations (7.24) and (7.25), respectively. Transitions that increase the values include death from all model compartments except those with CD4<200 and death among persons with HIV with CD4<200 and death. No transitions decrease the number of individuals in these states.

$\frac{dX_{p}^{c=29}(t)}{dt}={[1+\ddot{C}_{t}^{s=8}]\mu}_{p}\sum_{c=1}^{5} \left[ X_{p}^{c}(t) \right]+[1+\ddot{C}_{t}^{s=8}]\sum_{c=6}^{23} \left[ \sigma_{p}^{c}*X_{p}^{c}(t) \right]$	(7.24)

$\frac{dX_{p}^{c=30}(t)}{dt}=[1+\ddot{C}_{t}^{s=8}]\sum_{c=24}^{28} [\sigma_{p}^{c}X_{p}^{c}(t)]$	(7.25)

### 8. Analysis Scenarios

#### 8.1 Model Inputs For Disparities Analysis

This section describes the model inputs used to differentiate the baseline and intervention scenarios for our analysis. Table 8.1 includes the inputs used to produce the targeted model outputs in Table 8.2. We also describe how PrEP targets were determined to match AHEAD data.

Table 8.1	Relevant model inputs for the five different scenarios

| Input value | Population | Baseline | Continuum-only | Prevention-only | Continuum + Prevention | Maximum Feasible |
| --- | --- | --- | --- | --- | --- | --- |
| Relative risk of getting tested versus reference case^1^ | Black | 1.00 | 1.00 | 1.00 | 1.00 | 1.00 |
|  | H/L | 0.86 | 1.45 | 0.86 | 1.45 | 1.45 |
|  | Other | 1.21 | 1.00 | 1.21 | 1.00 | 1.00 |
| Annual probability of being LTC at diagnosis, by r/e (1 $\to$3)^2^ | Black | 0.79 | 0.86 | 0.79 | 0.86 | 0.98 |
|  | H/L | 0.83 | 0.86 | 0.83 | 0.86 | 0.98 |
|  | Other | 0.86 | 0.86 | 0.86 | 0.86 | 0.86 |
| Annual probability of being LTC after diagnosis, by r/e  (2 $\to$3)^2^ | Black | 0.15 | 0.17 | 0.15 | 0.17 | 0.30 |
|  | H/L | 0.16 | 0.23 | 0.16 | 0.23 | 0.30 |
|  | Other | 0.14 | 0.14 | 0.14 | 0.14 | 0.14 |
| Relative risk of initiating ART vs. HIV-stage-specific average ART initiation rates, by r/e (3 $\to$ 4/5)^2^ | Black | 1.00 | 1.01 | 1.00 | 1.01 | 2.00 |
|  | H/L | 0.99 | 1.01 | 0.99 | 1.01 | 2.00 |
|  | Other | 1.01 | 1.01 | 1.01 | 1.01 | 1.01 |
| Annual probability of becoming VLS if on ART but not VLS, by r/e  (4 $\to$5)^2^ | Black | 0.53 | 0.61 | 0.53 | 0.61 | 0.98 |
|  | H/L | 0.47 | 0.61 | 0.47 | 0.61 | 0.98 |
|  | Other | 0.61 | 0.61 | 0.61 | 0.61 | 0.61 |
| Annual probability of dropping off ART if on ART but not VLS, by r/e (4 $\to$3)^2^ | Black | 0.25 | 0.19 | 0.25 | 0.19 | 0.02 |
|  | H/L | 0.25 | 0.19 | 0.25 | 0.19 | 0.02 |
|  | Other | 0.19 | 0.19 | 0.19 | 0.19 | 0.19 |
| Annual probability of losing VLS and moving to on-ART-not-VLS, by r/e (5 $\to$ 4)^2^ | Black | 0.25 | 0.14 | 0.25 | 0.14 | 0.02 |
|  | H/L | 0.14 | 0.12 | 0.14 | 0.12 | 0.02 |
|  | Other | 0.19 | 0.19 | 0.19 | 0.19 | 0.19 |
| Number of uninfected PWID served by r/e targeted SSPs annually | Black | 78,368 | 78,368 | 114,020 | 114,020 | 117,355 |
|  | H/L | 78,547 | 78,547 | 81,526 | 81,526 | 83,686 |
|  | Other | 161,582 | 161,582 | 161,582 | 161,582 | 165,434 |
| Annual rate per eligible person of initiating PrEP targeted by r/e, transmission risk group, and sex | Black HET M | 0.0022 | 0.0022 | 0.0650 | 0.0650 | 0.0650 |
|  | H/L HET M | 0.0030 | 0.0030 | 0.0182 | 0.0182 | 0.0182 |
|  | Other HET M | 0.0025 | 0.0025 | 0.0025 | 0.0025 | 0.0010 |
|  | Black HET F | 0.0040 | 0.0040 | 0.0650 | 0.0650 | 0.0650 |
|  | H/L HET F | 0.0031 | 0.0031 | 0.0189 | 0.0189 | 0.0182 |
|  | Other HET F | 0.0021 | 0.0021 | 0.0021 | 0.0021 | 0.0010 |
|  | Black MSM | 0.5183 | 0.5183 | 1.0000 | 1.0000 | 1.0000 |
|  | H/L MSM | 0.2191 | 0.2191 | 0.6000 | 0.6000 | 0.9000 |
|  | Other MSM | 0.2651 | 0.2651 | 0.2651 | 0.2651 | 0.2200 |
|  | Black PWID | 0.0042 | 0.0042 | 0.0650 | 0.0650 | 0.0650 |
|  | H/L PWID | 0.0059 | 0.0059 | 0.0352 | 0.0352 | 0.0352 |
|  | Other PWID | 0.0112 | 0.0112 | 0.0112 | 0.0112 | 0.0112 |
| Annual rate of dropping off of PrEP | Oral | 0.92 | 0.92 | 0.92 | 0.92 | 0.72 |
|  | Injectable | 0.19 | 0.19 | 0.19 | 0.19 | 0.16 |

F = females; H/L = Hispanic/Latino; HET = Heterosexual; M = males; MSM = men who have sex with men; Other = race/ethnicities outside of Black and H/L; PWID = persons who inject drugs

^1^ Reference case corresponds to: Black, HET, CD4 > 500, age 13–17 years.

^2^ The numbers correspond to the following stages on the HIV continuum-of-care: 1. Undiagnosed; 2. Diagnosed, not in care; 3. In care, not on antiretroviral therapy (ART); 4. On ART, not virally suppressed (VLS); 5. VLS.

Table 8.2	Key health equity indicators in 2027 for the five different scenarios

| Continuum-of-care indicator (2027) | **Racial/ ethnic group** | Baseline | Scenario 1 (Continuum of Care) | Scenario 2 (Prevention) | Scenario 3 (Continuum of Care and Prevention) | Scenario 4 (Maximum Feasible) |
| --- | --- | --- | --- | --- | --- | --- |
| Percentage aware among PWH | Overall | 93.2% | 94.6% | 93.6% | 94.8% | 96.9% |
|  | Black | 93.3% | 94.5% | 93.7% | 94.8% | 97.5% |
|  | Hispanic/Latino | 90.7% | 94.5% | 91.4% | 94.9% | 97.6% |
|  | Other | 95.1% | 94.8% | 95.2% | 94.8% | 95.5% |
| Percentage linked to care among diagnosed PWH | Overall | 87.1% | 90.0% | 87.2% | 90.0% | 95.2% |
|  | Black | 85.4% | 89.9% | 85.5% | 90.0% | 97.7% |
|  | Hispanic/Latino | 86.1% | 90.2% | 86.3% | 90.4% | 97.4% |
|  | Other | 89.6% | 89.9% | 89.6% | 89.9% | 90.0% |
| Percentage viral load suppressed among diagnosed PWH, by year | Overall | 67.1% | 74.6% | 67.3% | 74.8% | 88.9% |
|  | Black | 61.8% | 75.3% | 62.0% | 75.4% | 95.9% |
|  | Hispanic/Latino | 66.3% | 74.7% | 66.6% | 75.0% | 95.2% |
|  | Other | 73.5% | 73.9% | 73.5% | 73.9% | 74.2% |
| Percentage in syringe services programs among PWID | Overall | 78.5% | 78.4% | 88.0% | 87.9% | 90.0% |
|  | Black | 60.6% | 60.5% | 88.1% | 87.9% | 90.1% |
|  | Hispanic/Latino | 84.7% | 84.6% | 87.9% | 87.8% | 89.9% |
|  | Other | 88.0% | 88.0% | 88.0% | 88.0% | 90.0% |
| Percentage on PrEP among PrEP-eligible at the end of the year^1^ | Overall | 31.3% | 31.5% | 59.5% | 59.7% | 70.7% |
|  | Black | 11.5% | 11.6% | 59.3% | 59.5% | 73.2% |
|  | Hispanic/Latino | 22.2% | 22.3% | 59.7% | 60.0% | 82.4% |
|  | Other | 59.5% | 59.6% | 59.5% | 59.6% | 59.6% |

^1^In HOPE, the number and percentage of MSM and HETs eligible for PrEP reflect findings from RTI’s analyses of PrEP indications reported among National Health and Nutrition Examination Survey (NHANES) respondents from 2009 through 2016 as described in Section 5. The resulting numbers of people with indications for PrEP in HOPE exceed the numbers in HIV surveillance reports. To better match HIV surveillance on the number of people eligible for PrEP for this analysis, we used 2022 HIV surveillance estimates when creating the scenarios for increasing PrEP use among Black and Hispanic/Latino groups.

These were based on numbers reported to be eligible for PrEP in 2020 among persons aged 16 years or older as reported by race and ethnicity in the AHEAD dashboard (USHHS, 2022). The same numbers were reported for 2019 in a previous surveillance report. We adjusted the numbers in each race and ethnicity group to match the overall total of 1,216,210 in 2019. We then applied an 0.54 percent annual growth rate, in line with historical population growth, to estimate that 1,269,754 people will be eligible for PrEP in 2027. We distributed the race- and ethnicity-specific numbers eligible for PrEP by applying the distributions across MSM, HET females, HET males, and PWID by race and ethnicity implied by the 2015 numbers eligible for PrEP reported in Smith, Van Handel, and Grey (2018; estimated distributions available on request). These AHEAD-eligible numbers were used as denominators for calculating percentage targets for PrEP in the current analysis. In other words, we determined the desired numbers on PrEP as 80% of AHEAD-eligible MSM and 50% of AHEAD-eligible HETs and PWID.

Using the AHEAD-eligible estimates, we calculated the estimated percentage of people eligible for PrEP who were on PrEP in the base and each alternative scenario. In the base scenario, 31.3% of people were estimated to be on PrEP (397,803) in 2027, with the highest percentage on PrEP estimated at 59.5% for the other group. The base scenario percentage on PrEP was 11.5% for the Black subgroup and 22.2% for the Hispanic/Latino group. Scenarios 2 and 3 increased the PrEP initiation rates by transmission group for Black and Hispanic/Latino people at high risk of HIV infection to result in percentages on PrEP of approximately 59.5% in 2027.

For the Maximum Feasible scenario, we sought to raise the percentages on PrEP for Black and Hispanic/Latino groups to 80% for MSM and 50% for HETs and PWID. We maintained the AHEAD percentage on PrEP of 59.5% for the other group. To achieve such large increases in the numbers on PrEP for Black and Hispanic/Latino groups, we increased the PrEP initiation rates by race and ethnicity and transmission group and reduced the dropout rates for PrEP. This required increasing PrEP initiation rates for Black MSM to 1.0 and for Hispanic/Latino MSM to 0.9. The resulting numbers estimated to be on PrEP in this scenario were almost 900,000 (359,000 for the Black group, 270,000 for the Hispanic/Latino group, and remaining at about 269,000 for the other group). The percentages on PrEP were 73.2% for Black AHEAD PrEP-eligible individuals (up from 11.5% in the base scenario) and 82.4% for Hispanic AHEAD PrEP-eligible individuals (up from 22.2% in the base scenario).

### 9. Calculation of Model Outcomes

In this section, we define the relevant inputs and methods applied in the calculation of the model’s key health outcomes, including HIV incidence and prevalence. Outcomes were collected for each analysis over a defined outcome collection period, which may or may not cover the entire model time horizon.

#### 9.1 Health Outcomes

Key health outcomes reported by the model include total number of new infections and HIV prevalence for all individuals actively moving through the model. They were calculated by using Equations (8.1) to (8.2), respectively.

$\text{F = }\sum_{\text{t}} \sum_{\text{p}} \left[ \left( \lambda_{p}(t) \right)\text{X}_{\text{p}}^{\text{c=1}}\text{(t)+}\left[ \left( 1-i_{p}* \Xi_{a*=1}^{N=1} \right)\lambda_{p}\left( t \right) \right]\text{X}_{\text{p}}^{\text{c=2}}\text{(t)+}\left[ \left( 1-i_{p}* \Xi_{a*=2}^{N=1} \right)\lambda_{p}\left( t \right) \right]\text{X}_{\text{p}}^{\text{c=3}}\text{(t)+}\left[ \left( 1-i_{p}* \Xi_{a*=1}^{N=2} \right)\lambda_{p}\left( t \right) \right]\text{X}_{\text{p}}^{\text{c=4}}\text{(t)+}\left[ \left( 1-i_{p}* \Xi_{a*=2}^{N=2} \right)\lambda_{p}\left( t \right) \right]\text{X}_{\text{p}}^{\text{c=5}}\text{(t)} \right]$	(8.1)

$\text{P}_{\text{t}}\text{ = }\sum_{c=6:28} \sum_{p} \left( X_{p}^{c}(t) \right)$	(8.2)

where

- F = cumulative number of new HIV infections (incidence) over the outcome collection period;
- P_t_ = HIV prevalence at time t;

#### 9.2 Economic Outcomes

In this section, we define the relevant inputs and methods applied in the calculation of the model’s economic outcomes, including the costs of testing, notification, and HIV treatment and care. All economic outcomes were computed using 2021 as a common cost year (Bureau of Labor Statistics, 2022).

##### 9.2.1 Testing Cost Inputs

Costs are applied for testing and notification. Testing is assumed to occur in both clinical and non-clinical settings. During scenarios in which an outreach intervention is applied, an additional outreach cost per test is also applied. Table 9.3 reports the inputs applied in the model used for calculating the costs related to testing.

Table 9.3.	Testing Cost Inputs

| Input | Value^a^ | Source |
| --- | --- | --- |
| Testing cost in clinical setting, including screen and second and third tests | | |
| Individual without HIV, rapid screen | 2010-2015: $22.13 (2012$)  2016+: $19.62 (2014$) | 2010-2015: Based on cost components from Hutchinson et al. (2011), Pinkerton et al. (2010), and Farnham et al. (2008)  2016+: Based on an average of INSTI, Determine, Unigold POC rapid cost (Hoenigl et al., 2015) and test performance data for Geenius (Bio-Rad, 2014). |
| Individual without HIV, conventional screen | 2010-2015: $8.24 (2012$)  2016+: $10.36 (2012$) | Based on cost components from Farnham et al. (2008) and Hutchinson et al. (2011, 2013), and adjusted to 2012$. |
| Individual with HIV, rapid screen | 2010-2015: $86.70 (2012$)  2016+: $66.70 (2014$) | 2010-2015: Based on cost components from Hutchinson et al. (2011, 2013), Pinkerton et al. (2010), and Farnham et al. (2008) and adjusted to 2012$. Assumes a repeat screen and a Western blot second test  2016+: Based on an average of INSTI, Determine, Unigold POC rapid cost (Hoenigl et al., 2015) and test performance data for Geenius (Bio-Rad, 2014). |
| Individual with HIV, conventional screen | 2010-2015: $60.02 (2012$)  2016+: $58.91 (2012$) | Based on cost components from Farnham et al. (2008) and Hutchinson et al. (2011, 2013) and adjusted to 2012$. Assumed Western blot second test before 2017, and a Geenius HIV 1/2 second test in 2017 and after. |
| NAT, applied for discrepant Western blot second test | $160.07 (2012$) | Hutchinson et al. (2013) |
| Additional cost per test performed in non-clinical (vs. clinical) setting | $52.66 (2005$) | Shrestha et al. (2008) |
| Notification costs |  |  |
| HIV-uninfected | $0.45 (2009$) | Hutchinson et al. (2011) |
| PWH, conventional screen | $5.88 (2009$) | Hutchinson et al. (2011) |
| PWH, rapid screen | $10.86 (2009$) | Hutchinson et al. (2011) |
| Outreach cost per test (when applied) | $13.67 (2005$) | Shrestha et al. (2008) |

Note: NAT = HIV nucleic acid amplification test

^a^ All cost inputs were converted to 2021$ in the calculation of economic outcomes.

##### 9.2.2 Calculation of Testing and Notification Costs

We calculated the cost of testing and notification by using Equations (9.5) to (9.9). The cost of testing varies by the type of test (rapid or conventional), the test result (positive or negative), the test setting (clinical versus non-clinical), and the test sensitivity, which varies by type of test. The cost for a NAT (HIV nucleic acid amplification test) is applied when the second test after a positive result is negative. Notification costs also vary by the type of test and test result. We assume the probability of notification does not change if a NAT is conducted. For ease of understanding, the calculations have been provided in words rather than symbols.

###### Number of Tests

Number of positive tests of individuals in *p* with HIV status *h*, taking test type *g*, at time *t* =
[Number of undiagnosed PWH in *p* with HIV status *h*]
x [Testing rate over time *t*, by *h* and *p*]
x [1 – (effect of COVID pandemic at time *t* on testing rates of people with HIV)]
x [Percentage of tests that are type *g*, by *p*]
x [Sensitivity of test type *g*, by *h*]
for *h* = {1 to 5} and all *p*, *g*, and *t*.	(9.5)

Number of negative tests of PWH (missed diagnoses) in *p* with HIV status *h*, taking test type *g*, at time *t* =
[Number of undiagnosed PWH in *p* with HIV status *h*]
x [Testing rate over time *t*, by *h* and *p*]
x [1 – (effect of COVID pandemic at time *t* on testing rates of people with HIV)]
x [Percentage of tests that are type *g*, by *p*]
x [1 − (Sensitivity of test type *g*, by *h*)]
for *h* = {1 to 5} and all *p*, *g*, and *t*.	(9.6)

Number of negative tests of HIV-uninfected (HIV status *h=0*) individuals in *p*, taking test type *g*, at time *t* =
[Number of people without HIV in *p*]
x [Testing rate over time *t*, by *h* and *p*]
x [1 – (effect of COVID pandemic at time *t* on testing rates of people without HIV)]
x [Percentage of tests that are type *g*, by *p*]
for *h* = {0} and all *p*, *g*, and *t*.	(9.7)

The total number of tests conducted is the sum of all positive and negative tests of PWH (calculated using Equations [9.5] and [9.6], respectively) and all negative tests of people without HIV (calculated using Equation [9.7]) across all *p*, *h*, *g*, and *t*.

###### Costs of Testing

Cost of testing individuals with HIV status *h* in *p* taking test type *g*, at time *t* =
([Number of positive tests of PWH in *p* with HIV status *h* = {1 to 5}, taking test type *g*, at time *t*] x [(Cost of positive test type *g* at *t*) + (Percentage of tests performed in non-clinical settings) x (Additional cost per test performed in non-clinical setting) + (1 − [Test sensitivity of second test, by HIV status *h*]) x (Cost of NAT)]
+ [Number of negative tests of individuals in *p* with HIV status *h*, taking test type *g*, at time *t*] x [(Cost of negative test type *g* at *t*) + (Percentage of tests performed in non-clinical settings) x (Additional cost per test performed in non-clinical setting)]
+ [Outreach cost per test, if applicable]) x (Discount factor at *t*)
for all *h*, *p*, *g*, and *t.*	(9.8)

The total cost of testing is the sum of the cost of testing (calculated using Equation [9.8]) across all *h*, *p*, *g*, and *t*.

###### Notification Costs

Cost of notification of HIV test results for individuals in *p* with HIV status *h* taking test type *g* at time *t* = (Probability of notification, by p and test type *g*, at time t)
x ([Number of positive tests of individuals in *p* with HIV status *h*, taking test type *g*, at time *t*] x [Cost of notification of positive results from test type *g*] + [Number of negative tests of individuals in *p* with HIV status *h*, taking test type *g*, at time *t*] x [Cost of notification of negative results from test type *g*]) x (Discount factor at *t*)
for all *h*, *p*, *g*, and *t.*	(9.9)

The total notification cost is the sum of the cost of notification (calculated using Equation [9.9]) across all *h*, *p*, *g*, and *t*.

##### 9.2.3 Calculation of Transitions Between HIV Continuum-of-Care Stages and Maintaining Viral Load Suppression

Costs are incurred for each individual who successfully transitions from one continuum-of-care stage (*r*) to a higher continuum-of-care stage. We also apply a cost for each individual who maintains VLS. These costs are reported in Table 9.4. We applied the same costs for these transitions and for maintenance of VLS as we estimated for the cost of an intervention that effectively results in the corresponding transition (as reported in Table 8.2) or lack thereof, in the case of maintenance of VLS.

Table 9.4.	Costs of Transitions along the HIV Continuum-of-Care and Maintenance of VLS

| Cost | Value | Source |
| --- | --- | --- |
| Cost for testing, diagnosis and notification of all tested individuals per unaware PWH effectively diagnosed (*r* = 1 to *r* = 2, *r* = 1 to *r* = 3) | $39,548 (2021$) per effectively diagnosed PWH | Equal to testing cost, as reported in Table 8.2. |
| Cost to effectively link one newly diagnosed person to care (*r* = 1 to *r* = 3) | $830 (2012$) | Assumed same as cost of linkage to care at diagnosis intervention, as reported in Table 8.2 |
| Cost to effectively link one previously diagnosed person to care (*r* = 2 to *r* = 3) | $1,660 (2012$) | Assumed same as cost of linkage to care after diagnosis intervention, as reported in Table 8.2 |
| Cost to prescribe ART to one individual linked to care but not on ART (*r* = 3 to *r* = 4, *r* = 3 to *r* = 5) | $131 (2021$) | Assumed same as cost of ART initiation intervention, as reported in Table 8.2 |
| Cost of one individual on ART but who is not VLS becoming VLS (*r* = 4 to *r* = 5) | $813 (2016$) | Assumed same as cost of ART adherence intervention, as reported in Table 8.2 |
| Annual cost of an individual who maintains VLS (and preventing loss of VLS from *r* = 5 to *r* = 4) | $813 (2016$) | Assumed same as cost of ART adherence intervention, as reported in Table 8.2 |

Note: ART = antiretroviral therapy, VLS = viral load suppression

##### 9.2.4 Calculation of HIV Treatment and Care Costs

HIV treatment and care costs, listed in Table 9.5, vary by both HIV status and care continuum status. The total HIV treatment and care costs accrued by the modeled population over the outcome collection period are calculated by using Equation (9.10).

Total treatment and care costs are calculated using Equation (9.10):

$\text{D = }\sum_{\text{p}} \sum_{\text{t}} \sum_{\text{c=6:28}} \left( \text{X}_{\text{p}}^{\text{c}}\left( \text{t} \right)\text{*}\text{J}_{\text{c}}\text{*}\frac{\text{1}}{\text{(1+U)}^{\text{t}}}\text{*q(t)} \right)$	(9.10)

where

- D = cumulative treatment and care costs over the outcome collection period,
- U = discount rate for costs, and
- J_c_ = annual treatment and care costs for an individual in compartment c

Table 9.5.	Annual HIV Treatment and Care Costs

| HIV Status | Care-Continuum Stage | Value^a^ | Source |
| --- | --- | --- | --- |
| Uninfected | N/A | $0 | Assumed |
| Acute | Unaware of infection | $4,372^b^ | Gebo et al. (2010), U.S. Department of Veterans Affairs (2018), Bingham et al. (2021) |
|  | Aware but not linked to HIV care | $4,372^b^ |  |
|  | Linked to HIV care but not on ART | $4,853^b^ |  |
| CD4 >500 | Unaware of infection | $4,372^b^ |  |
|  | Aware but not linked to HIV care | $4,372^b^ |  |
|  | Linked to HIV care but not on ART | $4,853^b^ |  |
|  | On ART | $34,846 |  |
| CD4 350–500 | Unaware of infection | $4,681^b^ |  |
|  | Aware but not linked to HIV care | $4,681^b^ |  |
|  | Linked to HIV care but not on ART | $5,255^b^ |  |
|  | On ART | $35,441 |  |
| CD4 200–350 | Unaware of infection | $5,978^b^ |  |
|  | Aware but not linked to HIV care | $5,978^b^ |  |
|  | Linked to HIV care but not on ART | $6,684^b^ |  |
|  | On ART | $37,560 |  |
| CD4 < 200 | Unaware of infection | $13,453^b^ |  |
|  | Aware but not linked to HIV care | $13,453 |  |
|  | Linked to HIV care but not on ART | $14,716 |  |
|  | On ART | $49,467 |  |

^a^ Costs in all stages include healthcare utilization costs (Inpatient, outpatient, emergency department) estimated from Gebo et al. (2010). Costs for “Linked to HIV care but not on ART” and “On ART” also include CD4 and viral load tests, genotype costs, and phenotype costs from the same source. ARV costs for those in ART health states based on the average cost of ART regimens as described in Bingham et al. (2021) using 2019 Federal Supply Schedule annual drug acquisition costs.

^b^ Reported in 2006$ for all costs other than “On ART” and 2019$ for “On ART”; converted to 2021$ in the calculation of economic outcomes.

##### 9.2.5 Calculation of SSP Costs

The annual costs for SSP are incurred for all PWID participating in a. The annual costs for SSP are the same as those outlined in Table 8.2. The total SSP costs accrued by the modeled population over the outcome collection period are discounted.

##### 9.2.6 Calculation of PrEP Costs

The annual (pro-rated) costs for PrEP listed in Table 9.6 are incurred for all individuals on PrEP. The total PrEP costs accrued by the modeled population over the outcome collection period are discounted. These costs assume HIV testing while receiving PrEP every 3 months.

Table 9.6.	Annual PrEP Costs

| Input | Value | Source |
| --- | --- | --- |
| Annual drug costs given full adherence | Oral: $10,249 (2022$)  Injectable: $21,865 (2022$) | Oral: U.S. Department of Veterans Affairs (2022); Huang et al. (2022b)^a^  Injectable: U.S. Department of Veterans Affairs (2022)^b^ |
| Annual screening, monitoring and administration costs given full adherence | Oral: $1,797 (2017$)  Injectable: $1,797 (2017$) | Shrestha et al. (2022) |
| Total annual costs given full adherence | Oral: $11,946^c^ (2021$)  Injectable: $23,233^c^ (2021$) |  |
| Percentage of full annual cost for oral PrEP to account for less than full (daily) adherence | High adherence: 88%  Low adherence: 22% | Landovitz et al., (2022)^d^ |
| Percentage of full annual cost for oral PrEP to account for less than full (monthly) adherence | High adherence: 89%  Low adherence: 62% | Assumed^e^ |

^a^ Calculation of weighted average annual costs of Truvada, Descovy, and generic emtricitabine-tenofovir (Truvada) costs reported in April 2022 Federal Supply Schedule (U.S. Department of Veterans Affairs, 2022). Monthly costs weighted by usage reported in Huang et al. (2022b).

^b^ April 2022 Federal Supply Schedule (U.S. Department of Veterans Affairs, 2022) for bi-monthly CABOTEGRAVIR 600MG/3ML INJ,SUSP,SA,KIT

^c^ Sum of annual drug costs and annual screening, monitoring and administration costs, converted to 2021$

^d^ High adherence to oral PrEP was defined as 4+ doses per week. In the HPTN 083 trial, 34.2% of an "adherence" subset of 372 HPTN083 (injectable PrEP trial for MSM) participants on oral PrEP took all 7 doses, 41.9% took 4-6.9 doses, 17.7% took some but <4 doses per week, and 5.3% no doses per week of samples over 81-week period (Landovitz et al., 2020). Weighted average of percentage of doses taken for high adherence is (34.2%*100%+41.9%*[4.0+6.9]/2/7)/(34.2%+41.9%) = 87.8%. Weighted average of percentage of doses taken for low adherence is (5.3%*0%+17.7%*[0.1+3.9]/2/7)/( 5.3%+17.7%) = 22.0%.

^e^ High adherence to injectable PrEP was defined as getting injections within 2 weeks of target date. Therefore, we assumed high adherence frequency to be every 9 weeks on average (mean of 8-10 weeks to reflect period within 2 weeks of target 8-week interval) and low adherence every 13 weeks on average (mean of 11 to 15 weeks, the 5-week period beyond 2 weeks after the target 8-week interval). These frequencies equate to 88.9% and 61.5% of injections, respectively, given within a year versus ideal frequency.

### 10. Model Calibration and Validation

#### 10.1 Model Calibration to Published Data

We calibrated a subset of the model’s inputs so that key model outcomes in the first time period approximated CDC surveillance data. This section outlines the process that was applied to calibrate the model.

##### 10.1.1 Establish Calibration Outcome Targets

The model outcomes targeted to match surveillance data are listed in Table 9.3. We aimed for the model’s outcomes to approximate the published point estimates within an acceptable range. We used published confidence intervals for those ranges when available; otherwise we established ranges based on relative percentage variation around the point estimates.

##### 10.1.2 Establish Inputs to Vary in Calibration

We identified inputs to vary and specified both a priority weight and a range of acceptable values for each (Tables 9.1 and 9.2). Most parameters selected to be varied were selected because limited to no source data were available. We also calibrated most rates of flow along the continuum-of care so that the distribution of PWH matched that observed in surveillance data, even though estimates of some of those rates (testing, linkage to HIV care, and other rates) are reported in the published literature. In addition, we also varied some inputs to which the model was highly sensitive but for which the published literature did not offer high confidence in specific values; per-act transmission risk was a key example of such an input.

Table 9.1.	Bounds and Final Values of Continuum-of-Care Parameters Varied in Calibration^a^

| Parameter | Calibrated Value | | Lower Bound | Upper Bound |
| --- | --- | --- | --- | --- |
| Annual rate of getting tested per reference case | | |  |  |
| Black, HET, CD4 > 500, 13-17 | *0.0790* | | 0.030 | 0.300 |
| Relative risk of getting tested versus reference case | | |  |  |
| By race/ethnicity | | |  |  |
| Hispanic/Latino | 0.8640 | | 0.500 | 1.500 |
| White/other | 1.2127 | | 0.100 | 1.500 |
| By transmission group |  | |  |  |
| MSM | 4.0310 | | 1.000 | 8.000 |
| PWID | 4.6357 | | 1.000 | 8.000 |
| By disease stage |  | |  |  |
| Acute | 0.9613 | | 0.500 | 1.500 |
| CD4 350–500 | 2.1291 | | 1.000 | 3.000 |
| CD4 200–350 | 5.2216 | | 1.250 | 6.000 |
| CD4 <200 | 4.3662 | | 1.500 | 6.000 |
| By age group |  | |  |  |
| 18–24 | 0.6348 | | 0.500 | 3.000 |
| 25–34 | 2.1245 | | 0.500 | 3.000 |
| 35–44 | 2.2451 | | 0.500 | 3.000 |
| 45–54 | 2.2803 | | 0.500 | 3.000 |
| 55–64 | 2.2693 | | 0.500 | 3.000 |
| 65+ | 2.1262 | | 0.500 | 3.000 |
| Annual probability of diagnosed individual linked to care after (versus immediately at) diagnosis | | |  |  |
| Black | 0.1457 | | 0.500 | 0.350 |
| Hispanic | 0.1611 | | 0.500 | 0.350 |
| White/other | 0.1356 | | 0.500 | 0.350 |
| Relative risk of linkage to HIV care after (versus immediately at) diagnosis, by disease stage | | |  |  |
| CD4 200–350 | | 6.7088 | 1.000 | 8.000 |
| CD4 < 200 | | 5.5057 | 1.000 | 8.000 |
| Annual probability of dropping out of care if linked to HIV care, not on ART | | |  |  |
| Black | | 0.4689 | 0.010 | 0.500 |
| Hispanic | | 0.4674 | 0.010 | 0.500 |
| White/other | | 0.4472 | 0.010 | 0.500 |

(continued)

Table 9.1.	Bounds and Final Values of Continuum-of-Care Parameters Varied in Calibration^a^ (continued)

| Parameter | | Calibrated Value | Lower Bound | Upper Bound |
| --- | --- | --- | --- | --- |
| Annual probability of dropping off of ART if ART-not-VLS and movement to linked-to- care | | |  |  |
| Black | 0.2456 | | 0.010 | 0.300 |
| Hispanic | 0.2524 | | 0.010 | 0.300 |
| White/other | 0.1942 | | 0.010 | 0.300 |
| Annual probability of departing from VLS and movement to ART-not-VLS | | |  |  |
| Black | 0.2502 | | 0.030 | 0.350 |
| Hispanic | 0.1370 | | 0.030 | 0.350 |
| White/other | 0.1917 | | 0.030 | 0.600 |
| Relative risk of losing VLS and moving to ART-not-VLS, by transmission group | | |  |  |
| MSM | 2.7241 | | 0.500 | 4.000 |
| PWID | 2.2686 | | 0.500 | 4.000 |
| Relative risk of losing VLS and moving to ART-not-VLS, by age group | | |  |  |
| 18–24 | 2.1103 | | 0.500 | 4.000 |
| 25–34 | 3.3217 | | 0.500 | 4.000 |
| 35–44 | 2.9540 | | 0.500 | 4.000 |
| 45–54 | 0.9078 | | 0.500 | 4.000 |
| 55–64 | 0.8251 | | 0.500 | 4.000 |
| 65+ | 0.9865 | | 0.500 | 4.000 |
| Relative risk of ART initiation by race/ethnicity, versus HIV-stage-specific average ART initiation rates | | | | |
| Black | | 1.0007 | 0.900 | 1.100 |
| Hispanic | | 0.9856 | 0.900 | 1.100 |
| White/other | | 1.0073 | 0.900 | 1.100 |
| Annual probability of becoming VLS if ART-not-VLS | | |  |  |
| Black | | 0.5307 | 0.010 | 0.700 |
| Hispanic | | 0.4728 | 0.010 | 0.700 |
| White/other | | 0.6064 | 0.010 | 0.700 |
| Relative risk of becoming VLS if ART-not-VLS, by transmission group | | |  |  |
| MSM | | 2.5789 | 0.200 | 3.000 |
| PWID | | 0.8112 | 0.200 | 3.000 |

(continued)

Table 9.1.	Bounds and Final Values of Continuum-of-Care Parameters Varied in Calibration^a^ (continued)

| Parameter | | Calibrated Value | | Lower Bound | | Upper Bound |
| --- | --- | --- | --- | --- | --- | --- |
| Relative risk of becoming VLS if ART-not-VLS, by age group | | |  | |  | |
| 18–24 | 1.4856 | | 0.500 | | 4.000 | |
| 25–34 | 0.7746 | | 0.500 | | 4.000 | |
| 35–44 | 2.8782 | | 0.500 | | 4.000 | |
| 45–54 | 2.7286 | | 0.500 | | 4.000 | |
| 55–64 | 2.6828 | | 0.500 | | 4.000 | |
| 65+ | 3.2693 | | 0.500 | | 4.000 | |

Note: ART = antiretroviral therapy; HET = heterosexual; PWID = people who inject drugs; VLS = viral load suppressed

^a^ Calibrated values reported in this version of the technical report are from calibration set LB20230224_2 .

Table 9.2.	Bounds and Final Values of Inputs Defining Behaviors and Infectivity Varied in Calibration^a^

| Parameter | Calibrated Value | Lower Bound | Upper Bound |
| --- | --- | --- | --- |
| Percentage of sexual partners by transmission group and sex (mixing matrix) | | | |
| HET M: HET F | 0.9670 | 0.950 | 0.999 |
| HET F: HET M | 0.9765 | 0.960 | 0.980 |
| HET F: PWID M | 0.0027 | 0.000 | 0.010 |
| MSM: HET F | 0.1314 | 0.000 | 0.450 |
| MSM: PWID F | 0.0025 | 0.000 | 0.010 |
| PWID M: PWID F | 0.4788 | 0.100 | 0.800 |
| PWID F: PWID M | 0.3947 | 0.100 | 0.800 |
| Percentage of sexual partners by number of HIV transmission risk factors (mixing matrix) | | | |
| HET—Fewer: Fewer | 0.9333 | 0.850 | 1.000 |
| HET/PWID—Multiple: Multiple | 0.7724 | 0.600 | 0.990 |
| MSM—Fewer: Fewer | 0.8226 | 0.600 | 0.990 |
| MSM—Multiple: Multiple | 0.6333 | 0.450 | 0.990 |
| Percentage of sexual partners by race (mixing matrix) for HETs and PWID | | | |
| Black: Hispanic | 0.0275 | 0.005 | 0.050 |
| Black: White/other | 0.0779 | 0.035 | 0.120 |
| Hispanic: Black | 0.0684 | 0.010 | 0.100 |
| Hispanic: White/other | 0.2698 | 0.150 | 0.400 |
| White/other: Black | 0.0345 | 0.010 | 0.100 |
| White/other: Hispanic | 0.0494 | 0.010 | 0.100 |

(continued)

Table 9.2.	Bounds and Final Values of Inputs Defining Behaviors and Infectivity Varied in Calibration^a^ (continued)

| Parameter | Calibrated Value | Lower Bound | Upper Bound |
| --- | --- | --- | --- |
| Percentage of sexual partners by race (mixing matrix) for MSM | | | |
| Black: Hispanic | 0.0248 | 0.005 | 0.050 |
| Black: White/other | 0.0904 | 0.035 | 0.250 |
| Hispanic: Black | 0.0790 | 0.010 | 0.100 |
| Hispanic: White/other | 0.3225 | 0.150 | 0.400 |
| White/other: Black | 0.0310 | 0.010 | 0.100 |
| White/other: Hispanic | 0.0426 | 0.010 | 0.100 |
| Percentage of needle-sharing partners by transmission group and sex (mixing matrix) | | | |
| PWID M: PWID F | 0.1864 | 0.10 | 0.30 |
| PWID F: PWID M | 0.4496 | 0.25 | 0.70 |
| Percentage of sexual partners by age groups for MSM (mixing matrix) | | | |
| 13–17:18–24 | 0.4296 | 0.200 | 0.600 |
| 13–17:25–34 | 0.2003 | 0.050 | 0.300 |
| 13–17:35–44 | 0.0256 | 0.000 | 0.050 |
| 13–17:45–54 | 0.0050 | 0.000 | 0.010 |
| 13–17:55–64 | 0.0050 | 0.000 | 0.010 |
| 13–17:65+ | 0.0050 | 0.000 | 0.010 |
| 18–24:13–17 | 0.1050 | 0.050 | 0.200 |
| 18–24:25–34 | 0.3770 | 0.200 | 0.500 |
| 18–24:35–44 | 0.1001 | 0.050 | 0.150 |
| 18–24:45–54 | 0.0232 | 0.000 | 0.050 |
| 18–24:55–64 | 0.0231 | 0.000 | 0.050 |
| 18–24:65+ | 0.0230 | 0.000 | 0.050 |
| 25–34:13–17 | 0.0399 | 0.000 | 0.080 |
| 25–34:18–24 | 0.2279 | 0.150 | 0.350 |
| 25–34:35–44 | 0.2436 | 0.150 | 0.350 |
| 25–34:45–54 | 0.0382 | 0.000 | 0.080 |
| 25–34:55–64 | 0.0246 | 0.000 | 0.050 |
| 25–34:65+ | 0.0249 | 0.000 | 0.050 |
| 35–44:13–17 | 0.0245 | 0.000 | 0.050 |
| 35–44:18–24 | 0.1004 | 0.050 | 0.150 |
| 35–44:25–34 | 0.2245 | 0.150 | 0.300 |
| 35–44:45–54 | 0.1894 | 0.100 | 0.300 |
| 35–44:55–64 | 0.0488 | 0.000 | 0.100 |
| 35–44:65+ | 0.0249 | 0.000 | 0.050 |

(continued)

Table 9.2.	Bounds and Final Values of Inputs Defining Behaviors and Infectivity Varied in Calibration^a^ (continued)

| Parameter | Calibrated Value | Lower Bound | Upper Bound |
| --- | --- | --- | --- |
| 45–54:13–17 | 0.0253 | 0.000 | 0.050 |
| 45–54:18–24 | 0.0453 | 0.010 | 0.080 |
| 45–54:25–34 | 0.0851 | 0.050 | 0.120 |
| 45–54:35–44 | 0.1998 | 0.100 | 0.300 |
| 45–54:55–64 | 0.2039 | 0.100 | 0.300 |
| 45–54:65+ | 0.0517 | 0.000 | 0.100 |
| 55–64:13–17 | 0.0050 | 0.000 | 0.010 |
| 55–64:18–24 | 0.0448 | 0.010 | 0.080 |
| 55–64:25–34 | 0.0538 | 0.010 | 0.100 |
| 55–64:35–44 | 0.0984 | 0.050 | 0.150 |
| 55–64:45–54 | 0.1956 | 0.100 | 0.300 |
| 55–64:65+ | 0.2043 | 0.100 | 0.300 |
| 65+:13–17 | 0.0050 | 0.000 | 0.010 |
| 65+:18–24 | 0.0050 | 0.000 | 0.010 |
| 65+:25–34 | 0.0242 | 0.000 | 0.050 |
| 65+:35–44 | 0.1183 | 0.050 | 0.200 |
| 65+:45–54 | 0.1419 | 0.050 | 0.250 |
| 65+:55–64 | 0.2410 | 0.100 | 0.400 |
| Base probability of transmission per condomless sex act | |  |  |
| Vaginal insertive | 0.0005 | 0.0001 | 0.0008 |
| Vaginal receptive | 0.0005 | 0.0004 | 0.0009 |
| Anal insertive | 0.0007 | 0.0003 | 0.0011 |
| Anal receptive | 0.0111 | 0.0080 | 0.0138 |
| Probability of transmission per shared needle | *0.0023* | 0.0010 | 0.005 |
| Percentage of injections that are shared | *0.1190* | 0.050 | 0.300 |
| Reduction in transmission per shared needle if VLS vs. not VLS | *0.7907* | 0.500 | 0.990 |
| Annual number of sexual contacts per uninfected person for MSM with multiple HIV transmission risk factors | |  |  |
| Black |  |  |  |
| 13–17 | 36.57 | 30.00 | 42.00 |
| 18–24 | 110.27 | 89.20 | 127.00 |

(continued)

Table 9.2.	Bounds and Final Values of Inputs Defining Behaviors and Infectivity Varied in Calibration^a^ (continued)

| Parameter | Calibrated Value | Lower Bound | Upper Bound |
| --- | --- | --- | --- |
| 25–34 | 111.04 | 87.60 | 125.00 |
| 35–44 | 79.25 | 71.20 | 100.00 |
| 45–54 | 76.63 | 57.00 | 81.00 |
| 55–64 | 56.65 | 55.00 | 78.00 |
| 65+ | 49.98 | 45.00 | 65.00 |
| Hispanic |  |  |  |
| 13–17 | 40.41 | 30.00 | 42.00 |
| 18–24 | 110.65 | 89.20 | 127.00 |
| 25–34 | 107.93 | 87.60 | 125.00 |
| 35–44 | 81.77 | 71.20 | 100.00 |
| 45–54 | 62.88 | 57.00 | 81.00 |
| 55–64 | 66.26 | 55.00 | 78.00 |
| 65+ | 60.31 | 45.00 | 65.00 |
| White/other |  |  |  |
| 13–17 | 31.50 | 30.00 | 42.00 |
| 18–24 | 91.78 | 89.20 | 127.00 |
| 25–34 | 122.98 | 87.60 | 125.00 |
| 35–44 | 82.28 | 71.20 | 100.00 |
| 45–54 | 65.44 | 57.00 | 81.00 |
| 55–64 | 71.39 | 55.00 | 78.00 |
| 65+ | 63.49 | 45.00 | 65.00 |
| Percentage of MSM sexual contacts that are insertive | |  |  |
| Black |  |  |  |
| 13–17 | 0.4270 | 20.0% | 80.0% |
| 18–24 | 0.3041 | 20.0% | 80.0% |
| 25–34 | 0.3568 | 20.0% | 80.0% |
| 35–44 | 0.4120 | 20.0% | 80.0% |
| 45–54 | 0.4871 | 20.0% | 80.0% |
| 55–64 | 0.5034 | 20.0% | 80.0% |
| 65+ | 0.4991 | 20.0% | 80.0% |
| Hispanic |  |  |  |
| 13–17 | 0.4346 | 20.0% | 80.0% |
| 18–24 | 0.3149 | 20.0% | 80.0% |

(continued)

Table 9.2.	Bounds and Final Values of Inputs Defining Behaviors and Infectivity Varied in Calibration^a^ (continued)

| Parameter | Calibrated Value | Lower Bound | Upper Bound |
| --- | --- | --- | --- |
| 25–34 | 0.3523 | 20.0% | 80.0% |
| 35–44 | 0.3770 | 20.0% | 80.0% |
| 45–54 | 0.4833 | 20.0% | 80.0% |
| 55–64 | 0.4938 | 20.0% | 80.0% |
| 65+ | 0.4936 | 20.0% | 80.0% |
| White/other |  |  |  |
| 13–17 | 0.5188 | 20.0% | 80.0% |
| 18–24 | 0.6061 | 20.0% | 80.0% |
| 25–34 | 0.6913 | 20.0% | 80.0% |
| 35–44 | 0.6653 | 20.0% | 80.0% |
| 45–54 | 0.6470 | 20.0% | 80.0% |
| 55–64 | 0.6340 | 20.0% | 80.0% |
| 65+ | 0.6275 | 20.0% | 80.0% |
| If participating in AI, percentage of male–female sexual partnerships that include AI | |  |  |
| 13–17 | 0.8007 | 65.0% | 95.0% |
| 18–24 | 0.8030 | 65.0% | 95.0% |
| 25–34 | 0.8007 | 65.0% | 95.0% |
| 35–44 | 0.7995 | 65.0% | 95.0% |
| 45–54 | 0.7995 | 65.0% | 95.0% |
| 55–64 | 0.7985 | 65.0% | 95.0% |
| 65+ | 0.7999 | 65.0% | 95.0% |

(continued)

Table 9.2.	Bounds and Final Values of Inputs Defining Behaviors and Infectivity Varied in Calibration^a^ (continued)

| Parameter | Calibrated Value | Lower Bound | Upper Bound |
| --- | --- | --- | --- |
| Length of time (in years) in each HIV stage if ART-not-VLS | |  |  |
| CD4 > 500 |  |  |  |
| 13-44 | 31.5496 | 5.00 | 50.00 |
| 45-64 | 38.8867 | 5.00 | 50.00 |
| 65+ | 32.4708 | 5.00 | 50.00 |
| CD4 350–500 |  |  |  |
| 13-44 | 24.3891 | 5.00 | 50.00 |
| 45-64 | 41.7688 | 5.00 | 50.00 |
| 65+ | 44.2003 | 5.00 | 50.00 |
| CD4 200–350 |  |  |  |
| 13-44 | 47.2797 | 5.00 | 50.00 |
| 45-64 | 22.4162 | 5.00 | 50.00 |
| 65+ | 16.1552 | 5.00 | 50.00 |
| Annual rate of progressing one disease stage while VLS | | | |
| CD4 > 500 | 0.0389 | 0.010 | 0.080 |
| CD4 350–500 | 0.0393 | 0.010 | 0.080 |
| CD4 200–350 | 0.0315 | 0.010 | 0.080 |
| Annual rate of improving one disease stage while VLS | | | |
| CD4 350–500 |  |  |  |
| 13-44 | 0.5067 | 0.25 | 0.65 |
| 45-64 | 0.4734 | 0.25 | 0.65 |
| 65+ | 0.4575 | 0.25 | 0.65 |
| CD4 200–350 |  |  |  |
| 13-44 | 0.5295 | 0.25 | 0.65 |
| 45-64 | 0.4949 | 0.25 | 0.65 |
| 65+ | 0.4723 | 0.25 | 0.65 |
| CD4 < 200 |  |  |  |
| 13-44 | 0.4900 | 0.25 | 0.65 |
| 45-64 | 0.5117 | 0.25 | 0.65 |
| 65+ | 0.4922 | 0.25 | 0.65 |

Note: AI = anal intercourse; ART = antiretroviral therapy; HET = heterosexual; PWID = people who inject drugs; VLS = viral load suppressed

^a^ Calibrated values reported in this version of the technical report are from calibration set LB20230224_2.

##### 10.1.3 Identification of Base and Alternative Input Sets

We then used MATLAB’s Optimization Toolbox (Mathworks; Natick, Massachusetts) to identify local optimal input sets starting with multiple sets of randomly sampled values for all calibrated inputs. We observed for each set of inputs the number of targets that were out of bounds and the two measures of goodness-of-fit: the “Out-of-bounds penalty measure” (inspired by a similar measure applied in Tian et al. [2016]) and the “target error measure,” as defined in Equations 9.1 and 9.2.

$\text{Out-of-bounds penalty measure for each input set:}\text{}$

$$\text{}$$

$\text{=}\sum_{i=1}^{NumTargets} \left( [{priority weight}_{i}]\left[ {penalty}_{i} \right]\left[ \frac{\left| {Model outcome value}_{i}-{Target value}_{i} \right|}{\left( {Model outcome value}_{i}+{Target value}_{i} \right)^{2}} \right] \right)$	(9.1)

where the penalty was set to 1000 if model outcome *i* was outside of the target range and 1 otherwise.

$\text{Target error measure for each input set:}\text{}$

$$\text{}$$

$\text{=}\sum_{i=1}^{NumTargets} \left( [{priority weight}_{i}]\left[ \frac{\left| {Model outcome value}_{i}-{Target value}_{i} \right|}{{Target value}_{i}} \right] \right)$	(9.2)

The optimization’s objective was to minimize the out-of-bounds penalty measure, given the model’s values, target values, and assigned priority weights for each targeted outcome. Among those optimized sets, we then selected one set to apply in the base analysis and 10 alternative sets for supporting uncertainty analyses. The base and alternative sets were selected so that they reflected the sets with both the lowest out-of-bounds penalty measure values and the fewest number of outcomes out of bounds.

The values in the identified base analysis set are reported in Tables 9.1 and 9.2. The model’s outcomes given the identified base analysis set are shown in Table 9.3.

Table 9.3.	Values Generated by the Model using the Base Analysis Set vs. Target Values and Bounds Considered for Outcomes Targeted in Calibration

| Outcome Name | | Model Values | | Target Value | Lower Bound | | Upper Bound | Priority Weight | Source for Target Values and Bounds |
| --- | --- | --- | --- | --- | --- | --- | --- | --- | --- |
| Estimated number of new infections in the United States in 2019 | | | | | | | |  | CDC (2021a) |
| Total | | *37,100* | | 34,800 | 32,600 | | 37,100 | 0.5 |  |
| Black | | 13,748 | | 14,300 | 12,900 | | 15,700 | 3 |  |
| Hispanic/Latino | | 11,589 | | 10,200 | 8,900 | | 11,600 | 3 |  |
| Other | | 11,763 | | 10,300 | 8,240 | | 12,360 | 3 |  |
| HET Male | | 2,828 | | 2,400 | 1,700 | | 3,100 | 0.5 |  |
| HET Female | | 5,514 | | 5,300 | 4,600 | | 6,100 | 0.5 |  |
| MSM | | 26,226 | | 24,420 | 19,536 | | 29,304 | 0.5 |  |
| PWID Male | | 1,746 | | 1,480 | 1,184 | | 1,776 | 0.5 |  |
| PWID Female | | 786 | | 1,100 | 750 | | 1,400 | 0.5 |  |
| Number of PWH deaths in 2019 | | | | | | | |  | CDC (2021b) |
| Total aware deaths | | 19,322 | | 15,463 | 12,370 | | 18,556 | 1 |  |
| Aware AIDS deaths | | 5,135 | | 4,794 | 3,835 | | 5,753 | 1 | CDC (2023)^a^ |
| Unaware AIDS deaths | | 343 | | 250 | 150 | | 350 | 1 | Krentz et al. (2014)^b^ |
| HIV prevalence in 2019 | |  | |  | |  |  |  | CDC (2021a) |
| Total | | *1,219,149* | | 1,189,700 | | 1,154,009 | 1,225,391 | 0.5 |  |
| Black | | 485,710 | | 479,300 | | 464,921 | 493,679 | 1 |  |
| Hispanic/Latino | | 294,658 | | 294,200 | | 285,374 | 303,026 | 1 |  |
| Other | | 438,781 | | 416,200 | | 403,714 | 428,686 | 1 |  |
| HIV prevalence in 2019 compared to 2010 | | | | | | | |  |  |
| Ratio of LR HET prevalence in 2019 vs 2010 | *1.07* | | 1.15 | | | 1.00 | 1.38 | 0.5 | CDC (2021a) |
| Ratio of HR HET prevalence in 2019 vs 2010 | *1.15* | | 1.15 | | | 1.00 | 1.38 | 0.5 |  |
| Distribution of PWH across the continuum of care in 2019 | | | | | | | |  | CDC (2021a) |
| Diagnosed | |  | |  | |  |  |  |  |
| Total | | *438,781* | | 86.7% | | 86.1% | 87.2% | 1 |  |
| Black | | 92.0% | | 86.6% | | 85.7% | 87.5% | 3 |  |
| Hispanic/Latino | | 89.5% | | 83.6% | | 82.6% | 84.7% | 3 |  |
| Other | | 94.3% | | 88.9% | | 86.3% | 91.6% | 3 |  |

(continued)

Table 9.3.	Values Generated by the Model using the Base Analysis Set vs. Target Values and Bounds Considered for Outcomes Targeted in Calibration (continued)

| Outcome Name | Model Values | Target Value | | Lower Bound | | Upper Bound | Priority Weight | Source for Target Values and Bounds |
| --- | --- | --- | --- | --- | --- | --- | --- | --- |
| Distribution of PWH across the continuum of care in 2019 (continued) | | | | | | |  | CDC (2021b) |
| In care |  |  | |  | |  |  |  |
| Total | *85.3%* | 76.0% | | 73.7% | | 78.3% | 1 |  |
| Black | 83.8% | 73.7% | | 71.4% | | 75.9% | 3 |  |
| Hispanic/Latino | 84.5% | 74.0% | | 71.8% | | 76.2% | 3 |  |
| Other | 87.4% | 79.9% | | 77.5% | | 82.3% | 3 |  |
| Viral load suppressed (among diagnosed) |  |  | |  | |  |  | CDC (2021b) |
| Total | *64.2%* | 64.7% | | 61.7% | | 67.7% | 1 |  |
| Black | 58.9% | 60.8% | | 56.8% | | 64.8% | 3 |  |
| Hispanic/Latino | 63.7% | 64.6% | | 60.6% | | 68.6% | 3 |  |
| Other | 70.3% | 71.3% | | 67.3% | | 75.3% | 3 |  |
| Population in 2019 |  |  | |  | |  |  | Derived from U.S. Census Bureau (2020) |
| Aged 13–34 | 86,454,887 | 86,456,215 | | 77,810,593 | | 95,101,836 | 0^a^ |  |
| Aged 35–64 | 111,429,292 | 111,423,643 | | 100,281,278 | | 122,566,007 | 0^a^ |  |
| Aged 65+ | 48,160,341 | 48,181,584 | | 43,363,425 | | 52,999,742 | 0^a^ |  |
| Estimated number of people on PrEP in the US in 2019 | | | | | | |  | USHHS (2022) |
| Total | *278,664* | 275,315 | 261,549 | | 289,081 | | 0^a^ |  |
| Male | 258,314 | 254,265 | 241,552 | | 266,978 | | 0^a^ |  |
| Female | 20,351 | 21,050 | 19,998 | | 22,103 | | 0^a^ |  |
| Black | 37,387 | 37,248 | 35,386 | | 39,110 | | 0^a^ |  |
| Hispanic/Latino | 44,729 | 45,473 | 43,199 | | 47,747 | | 0^a^ |  |
| Other | 196,548 | 192,594 | 182,964 | | 202,224 | | 0^a^ |  |
| Estimated number of people on PrEP in the US in 2021 | | | | | | |  | USHHS (2022) |
| Total | *370,244* | 366,819 | 348,478 | | 385,160 | | 0^a^ |  |
| Male | 342,042 | 338,732 | 321,795 | | 355,669 | | 0^a^ |  |
| Female | 28,203 | 28,087 | 26,683 | | 29,491 | | 0^a^ |  |
| Black | 51,186 | 51,891 | 49,296 | | 54,486 | | 0^a^ |  |
| Hispanic/Latino | 62,523 | 63,953 | 60,755 | | 67,151 | | 0^a^ |  |
| Other | 256,536 | 250,975 | 238,426 | | 263,524 | | 0^a^ |  |

Note: ART = antiretroviral therapy; HET = heterosexual; PWID = people who inject drugs; VLS = viral load suppressed; USHHS = U.S. Department of Health and Human Services.

^a^ CDC Wonder Database (2023). All deaths with AIDS listed as cause of death in 2019. 5,044 AIDS deaths among PWH were identified; 250 AIDS deaths among unaware PWH (Krentz et al., 2014) were subtracted to obtain an estimated 4,794 AIDS deaths among aware PWH.

^b^ Krentz et al. (2014) found that 3% of AIDS deaths occurring among the undiagnosed in Calgary, Canada. This was assumed to be slightly higher in the US, giving an estimate of roughly 250 (about 5% of 4,794).

#### 10.2 Internal and External Validation of the Model

We conducted a thorough quality check of the model’s inputs, calculations, and differential equations. An earlier version of this model was also reviewed by experts in differential equation modeling of HIV, Drs. Michael Pickles and Marie-Claude Boily, both of the Imperial College of London.

We compared our model’s outcomes to CDC surveillance data as listed in Table 9.3. Estimated new infections by transmission group and distributions of PWH across the continuum-of-care stages were used for calibration as described in Section 9.1. In section 10.2.1 below we discuss the strengths and limitations of calibration set LB20230224_2 for this analysis.

##### 10.2.1 Strengths and Limitations of Calibration Set LB20230224_2

Although the calibrated values for percent diagnosed and percent LTC among diagnosed in 2019 are higher than surveillance data for 2019 (CDC, 2021b), what matters most for this analysis is matching the direction and magnitude of differences between race and ethnicity groups. The calibrated values perform well according to this criterion.

For example, 2019 surveillance data show that percent diagnosed is highest for PWH in the other group (88.9%) and lowest for Hispanic PWH (83.6%); percent diagnosed among Black PWH are in the middle (86.6%). Our calibrated values for percent diagnosed follow this same pattern, with 94.3%, 89.5%, and 92.0% diagnosed among PWH for other, Hispanic, and Black groups, respectively.

Additionally, 2019 surveillance data show that linkage to care among diagnosed PWH is highest for the other group (79.9%), with lower, but similar, values for Hispanic (74.0%) and Black PWH (73.7%). Our calibrated values for percent linked to care among diagnosed PWH follow this same pattern, and are 87.4%, 84.5% and 83.8% for other, Hispanic, and Black groups, respectively. Further, the calibrated values for both continuum outcomes are still lower than, and therefore preferable to, calibrated values we observed in other calibration sets with similar performance.

Calibrated values for deaths among PWH in 2019 are slightly higher than the upper bound from surveillance data. Surveillance data show approximately 15,500 deaths in 2019, with bounds from 12,400 to 18,600. Our calibrated number of deaths is 19,600, slightly higher (5%) than the upper bound. However, the number of AIDS deaths, an important target to achieve, is within the range of surveillance data. Additionally, calibration sets that were within the surveillance target range for all deaths among PWH did not perform as well overall or in terms of care continuum absolute and relative values as our selected calibration set.

The calibrated value for the number of new infections in HET females is slightly above the range for this subgroup (6,300, where the surveillance target range is 4,600 to 6,100), but is only 3% higher than the upper bound, so we considered it an acceptable value.

### 11. Model Sensitivity and Uncertainty Analyses

To explore the HOPE model’s sensitivity to its input values for this particular analysis, we conducted an uncertainty analysis. We also examined the sensitivity of the model to the COVID-19 effects. Details on the methods and results of this analysis are outlined in the sections below.

#### 11.1 Uncertainty Analysis

Uncertainty analysis is used to evaluate a model outcome’s variability that is due to the uncertainty of model input values that are estimated. Unlike a one-way sensitivity analysis, which estimates the effect of the changes in the value of an individual input on the model outcome, uncertainty analysis considers the variability of the model outcome based on the collective uncertainty of the estimated input values.

To consider the uncertainty of the IRRs for the baseline scenario in 2035 due to the selected values of the calibrated parameters, we ran the model using 10 additional sets of values for those inputs that resulted in model outcomes that approximated the targeted surveillance measures. These results were run without COVID-19 effects. The same process and selection criteria were applied as for the set used in the base-case analysis, as outlined in Section 9.1.3. We focused on the baseline scenario results to observe how the model reflected current disparities in HIV care, given that the effects of the interventions on disparities would be dependent on the level of existing disparities. We then observed the range of the IRRs for Black vs Other populations and Hispanic vs Other populations in 2035 across all calibrated runs to assess the robustness of our findings.

We found that the IRRs for Black vs Other ranged from 6.52 to 9.04 in 2035, with a standard deviation of 0.80. The IRRs for Hispanic vs Other ranged from 3.52 to 4.87, with a standard deviation of 0.39. These findings demonstrate that extent of our current baseline disparity predictions are not a reflection of our chosen calibration set, which predicts an IRR of 6.52 in 2035 for Black vs Other and 4.13 for Hispanic vs Other, and may even underestimate the severity of the disparities if no future actions are taken. While the uncertainty analyses were run with COVID-19 effects, the IRRs for the baseline scenario with and without COVID-19 effects differed by <0.3%.

References

Adams M, An Q, Broz D, et al. (2019) Distributive Syringe Sharing and Use of Syringe Services Programs (SSPs) Among Persons Who Inject Drugs. AIDS Behav. 2019;23(12):3306-3314. doi:10.1007/s10461-019-02615-4

Ahmad FB, Cisewski JA, Miniño A, Anderson RN. Provisional Mortality Data — United States, 2020. Erratum: Vol. 70, No. 14. MMWR Morb Mortal Wkly Rep 2021;70:900. DOI: http://dx.doi.org/10.15585/mmwr.mm7024a4external icon

Althoff KN, Justice AC, Gange SJ, et al. Virologic and immunologic response to HAART, by age and regimen class. *AIDS*. 2010 Oct 23;24(16):2469–79. Erratum in: *AIDS*. 2011 Jan 28;25(3):397.

Anderson PL, Glidden DV, Liu A, Buchbinder S, Lama JR, Guanira JV, et al. Emtricitabine-tenofovir concentrations and pre-exposure prophylaxis efficacy in men who have sex with men. Sci Transl Med. 2012;4(151):151ra125. doi:10.1126/scitranslmed.3004006

Arias E. United States life tables, 2008. *National Vital Statistics Reports*. 2012;61(3). Hyattsville, MD: National Center for Health Statistics.

Aspinall EJ, Nambiar D, Goldberg DJ, Hickman M, Weir A, Van Velzen E, Palmateer N, Doyle JS, Hellard ME, Hutchinson SJ. Are needle and syringe programmes associated with a reduction in HIV transmission among people who inject drugs: a systematic review and meta-analysis. Int J Epidemiol. 2014 Feb;43(1):235-48.

Bavinton BR, Pinto AN, Phanuphak N, et al.; Opposites Attract Study Group. Viral suppression and HIV transmission in serodiscordant male couples: an international, prospective, observational, cohort study. Lancet HIV 2018;5:e438–47. 10.1016/S2352-3018(18)30132-2

Bio-Rad. Geenius HIV 1/2 supplemental assay: Instructions for use. Updated October 2014. Available at: <https://www.fda.gov/downloads/BiologicsBloodVaccines/BloodBloodProducts/ApprovedProducts/PremarketApprovalsPMAs/UCM420735.pdf>. Accessed September 11, 2018.

Boily MC, Pickles M, Lowndes CM, et al. Positive impact of a large-scale HIV prevention programme among female sex workers and clients in South India. *AIDS*. 2013 Jun 1;27(9):1449–60.

Bosh KA, Johnson AS, Hernandez AL, et al. (2020) Vital Signs: Deaths Among Persons with Diagnosed HIV Infection, United States, 2010-2018. MMWR Morb Mortal Wkly Rep. 2020 Nov 20;69(46):1717-1724. doi: 10.15585/mmwr.mm6946a1. PMID: 33211683; PMCID: PMC7676640.

Chavez P, Wesolowski L, Patel P, et al. Evaluation of the performance of the Abbott ARCHITECT HIV Ag/Ab combo assay. *J Clin Virol.* 2011;52(Suppl. 1, December):S51–5.

Centers for Disease Control and Prevention (2023). National Vital Statistics System, Mortality 2018-2021 on CDC WONDER Online Database, released in 2021. National Center for Health Statistics. Data are from the Multiple Cause of Death Files, 2018-2021, as compiled from data provided by the 57 vital statistics jurisdictions through the Vital Statistics Cooperative Program. Available at: <http://wonder.cdc.gov/ucd-icd10-expanded.html>. Accessed March 27, 2023.

Centers for Disease Control and Prevention. Monitoring selected national HIV prevention and care objectives by using HIV surveillance data—United States and 6 dependent areas, 2020. HIV Surveillance Supplemental Report 2022;27(No. 3). http://www.cdc.gov/hiv/library/reports/hiv-surveillance.html. Published May 2022. Accessed March 31, 2023.

Centers for Disease Control and Prevention (2021a). Estimated HIV incidence and prevalence in the United States, 2015–2019. HIV Surveillance Supplemental Report 2021;26(No. 1). http://www.cdc.gov/hiv/library/reports/hiv-surveillance.html. Published May 2021. Accessed June 11, 2021.

Centers for Disease Control and Prevention (2021b). Monitoring selected national HIV prevention and care objectives by using HIV surveillance data—United States and 6 dependent areas, 2019. HIV Surveillance Supplemental Report 2021;26(No. 2). http://www.cdc.gov/hiv/library/reports/hiv-surveillance.html. Published May 2021. Accessed June 11, 2021.

Centers for Disease Control and Prevention (2019a). Estimated HIV incidence and prevalence in the United States, 2010–2016. HIV Surveillance Supplemental Report 2019;24(No. 1). http://www.cdc.gov/ hiv/library/reports/hiv-surveillance.html. Published February 2019. Accessed June 21st, 2019.

Centers for Disease Control and Prevention (2019b). HIV Infection Risk, Prevention, and Testing Behaviors Among Men Who Have Sex With Men—National HIV Behavioral Surveillance, 23 U.S. Cities, 2017. HIV Surveillance Special Report 22. https://www.cdc.gov/hiv/library/reports/hiv-surveillance.html. Published February 2019. Accessed February 2, 2020.

Centers for Disease Control and Prevention (2019c). Effectiveness of prevention strategies to reduce the risk of acquiring or transmitting HIV. 19 November 2019. https://www.cdc.gov/hiv/risk/estimates/preventionstrategies.html. Accessed 7 July 2021.

Centers for Disease Control and Prevention (2018b). Diagnoses of HIV infection among adolescents and young adults in the United States and 6 dependent areas, 2011–2016. HIV Surveillance Supplemental Report 2018;23(No. 3). http://www.cdc.gov/hiv/library/reports/hivsurveillance.html. Published May 2018. Accessed July 12, 2019.

Centers for Disease Control and Prevention (2018c). Diagnoses of HIV infection among adults aged 50 years and older in the United States and dependent areas, 2011–2016. HIV Surveillance Supplemental Report 2018;23(No. 5). http://www.cdc.gov/hiv/library/reports/hivsurveillance.html. Published August 2018. Accessed July 15, 2019.

Centers for Disease Control and Prevention (2018d): US Public Health Service: Preexposure prophylaxis for the prevention of HIV infection in the United States—2017 Update: a clinical practice guideline. https://www.cdc.gov/hiv/pdf/risk/prep/cdc-hiv-prep-guidelines-2017.pdf. Published March 2018, Accessed May 2020

Centers for Disease Control and Prevention (2017a). CDC-Funded HIV Testing: United States, Puerto Rico, and the U.S. Virgin Islands, 2017. http://www.cdc.gov/hiv/library/reports/index.html. Published 2018. Accessed June 11, 2019.

Centers for Disease Control and Prevention (2017b). Preexposure prophylaxis for the prevention of HIV infection in the United States—2017 Update: a clinical practice guideline. Available at: <https://www.cdc.gov/hiv/pdf/risk/prep/cdc-hiv-prep-guidelines-2017.pdf>. Accessed March 17, 2022. .

Centers for Disease Control and Prevention (CDC). Monitoring selected national HIV prevention and care objectives by using HIV surveillance data—United States and 6 dependent areas—2013. *HIV Surveillance Supplemental Report, 2015*. 2015;20(No. 2). Available at: <http://www.cdc.gov/hiv/library/reports/surveillance/>. Accessed August 28, 2015.

Centers for Disease Control and Prevention (2014). Behavioral and Clinical Characteristics of Persons Receiving Medical Care for HIV Infection—Medical Monitoring Project, United States, 2010. HIV Surveillance Special Report 9. http://www.cdc.gov/hiv/library/reports/surveillance/#special. Published October 2014. Accessed 10Sept2019.

Chan PA, Patel RR, Mena L, et al. Long-term retention in pre-exposure prophylaxis care among men who have sex with men and transgender women in the United States. J Int AIDS Soc. 2019;22(8):e25385. doi:10.1002/jia2.25385

Chandra A, Mosher WD, Copen C, Sionean C. Sexual behavior, sexual attraction, and sexual identity in the United States: data from the 2006-2008 National Survey of Family Growth. Natl Health Stat Report. 2011;(36):1-36.

Chen YH, Farnham PG, Hicks KA, Sansom SL. Estimating the HIV Effective Reproduction Number in the United States and Evaluating HIV Elimination Strategies. Journal of Public Health Management and Practice. 2021 Aug 2.

Cohen MS, Chen YQ, McCauley M, et al. Antiretroviral therapy for the prevention of HIV-1 transmission. N Engl J Med 2016;375:830-9.

Coy KC, Hazen RJ, Kirkham HS, Delpino A, Siegler AJ. Persistence on HIV preexposure prophylaxis medication over a 2-year period among a national sample of 7148 PrEP users, United States, 2015 to 2017. J Int AIDS Soc. 2019;22(2):e25252. doi:10.1002/jia2.25252

Crepaz N, Song R, Lyss S, Hall HI. (2020) Trends in Time From HIV Diagnosis to First Viral Suppression Following Revised US HIV Treatment Guidelines, 2012-2017. J Acquir Immune Defic Syndr. 2020 Sep 1;85(1):46-50. doi: 10.1097/QAI.0000000000002398. PMID: 32379083.

Des Jarlais DC et al. (2020) Expansion of Syringe Service Programs in the United States, 2015-2018. Am J Public Health. 2020;110(4):517-519. doi:10.2105/AJPH.2019.305515

Des Jarlais DC, Nugent A, Solberg A, Feelemyer J, Mermin J, Holtzman D. Syringe service programs for persons who inject drugs in urban, suburban, and rural areas—United States, 2013. *MMWR Morb Mortal Wkly Rep*. 2015 Dec 11;64(48):1337-41.

Dodge B, Reece M, Herbenick D, et al. Sexual health among U.S. black and Hispanic men and women: a nationally representative study. *J Sex Med.* 2010;7(suppl 5):330–45.

Fiebig EW, Wright DJ, Rawal BD, et al. Dynamics of HIV viremia and antibody seroconversion in plasma donors: implications for diagnosis and staging of primary HIV infection. *AIDS*. 2003 Sep 5;17(13):1871–9.

Finlayson T, Le B, Smith A, et al. HIV risk, prevention, and testing behaviors among men who have sex with men—National HIV Behavioral Surveillance System, 21 U.S. cities, United States, 2008. *MMWR*. 2011;60(No. SS–14).

Fleishman, JA, Yehia BR, Moore RD, et al. Disparities in receipt of antiretroviral therapy among HIV-infected adults (2002–2008). *Medical Care*. 2012;50:419–27.

Gardner EM, McLees MP, Steiner JF, et al. The spectrum of engagement in HIV care and its relevance to test-and-treat strategies for prevention of HIV infection. *Clin Infect Dis*. 2011;52:793–800.

Glick SN, Morris M, Foxman B, et al. A comparison of sexual behavior patterns among men who have sex with men and heterosexual men and women. *J Acquir Immune Defic Syndr*. 2012;60:83–90.

Grant RM, Anderson PL, McMahan V, et al. Uptake of pre-exposure prophylaxis, sexual practices, and HIV incidence in men and transgender women who have sex with men: a cohort study. Lancet 2014;14:820-9.

Hall HI, Byers RH, Ling Q, Espinoza L. Racial/ethnic and age disparities in HIV prevalence and disease progression among men who have sex with men in the United States. American journal of public health. 2007 Jun;97(6):1060-6.

Handanagic S, et al. (2021) HIV Infection and HIV-Associated Behaviors Among Persons Who Inject Drugs - 23 Metropolitan Statistical Areas, United States, 2018. MMWR Morb Mortal Wkly Rep. 2021;70(42):1459-1465. Published 2021 Oct 22. doi:10.15585/mmwr.mm7042a1

Herbenick D, Reece M, Schick V, et al. Sexual behaviors, relationships, and perceived health status among adult women in the United States: results from a national probability sample. *J Sex Med*. 2010;7:277–90.

HIV Prevention Trials Network (HPTN). (2020). HPTN 084 -A Phase 3 Double Blind Safety and Efficacy Study of Long-Acting Injectable Cabotegravir Compared to Daily Oral TDF/FTC for Pre-Exposure Prophylaxis in HIV-Uninfected Women. Available at https://www.hptn.org/news-and-events/press-releases/hptn-084-study-demonstrates-superiority-of-cab-la-to-oral-tdfftc-for. Accessed June 24, 2021.

Holloway IW, Krueger EA, Meyer IH, Lightfoot M, Frost DM, Hammack PL. Longitudinal trends in PrEP familiarity, attitudes, use and discontinuation among a national probability sample of gay and bisexual men, 2016-2018. PLoS One. 2020;15(12):e0244448. Published 2020 Dec 31. doi:10.1371/journal.pone.0244448

Huang YA, Zhu W, Wiener J, Kourtis AP, Hall HI, Hoover KW, Impact of Coronavirus Disease 2019 (COVID-19) on Human Immunodeficiency Virus (HIV) Pre-exposure Prophylaxis Prescriptions in the United States—A Time-Series Analysis, Clinical Infectious Diseases, 2022a; <https://doi.org/10.1093/cid/ciac038>.

Huang YA, Tao G, Smith DK, Hoover KW. Persistence With Human Immunodeficiency Virus Pre-exposure Prophylaxis in the United States, 2012-2017. Clin Infect Dis. 2021;72(3):379-385. doi:10.1093/cid/ciaa037

Huang YA, Zhu W, Smith DK, Harris N, Hoover KW. HIV Preexposure Prophylaxis, by Race and Ethnicity — United States, 2014–2016. MMWR Morb Mortal Wkly Rep 2018;67:1147–1150. DOI: http://dx.doi.org/10.15585/mmwr.mm6741a3external icon.

Huang YA, Hutchinson AB, Hollis ND, et al. Notification following new positive HIV test results. *Int J STD & AIDS*. 2016;27(10):868-872. DOI: 10.1177/0956462415598090

Hutchinson AB, Ethridge SF, Wesolowski LG, et al. Costs and outcomes of laboratory diagnostic algorithms for the detection of HIV. *J Clin Virol*. 2013 Dec; 58 Suppl 1:e2–7.

Introcaso CE, Xu F, Kilmarx PH, et al. Prevalence of circumcision among men and boys aged 14 to 59 Years in the United States, National Health and Nutrition Examination Surveys 2005–2010. *Sex Transm Dis*. 2013;40:521–5.

Jacobson EU, Hicks KA, Carrico J, Purcell DW, Green TA, Mermin JH, Farnham PG. Optimizing HIV prevention efforts to achieve EHE incidence targets. JAIDS Journal of Acquired Immune Deficiency Syndromes. 2022 Apr 1;89(4):374-80.

Jacobson E, Hicks K, Tucker E, Farnham P, Sansom S. Effects of reaching national goals on HIV incidence, by race and ethnicity, in the United States. *J Public Health Manag Pract.* 2018 Jul 1;24(4):E1-8.

Juusola JL, Brandeau ML, Owens DK, et al. The cost-effectiveness of preexposure prophylaxis for HIV prevention in the United States in men who have sex with men. *Ann Intern Med*. 2012 Apr 17;156(8):541–50.

Kaplan EH, Heimer R. A model-based estimate of HIV infectivity via needle sharing. *J Acquir Immune Defic Syndr*. 1992;5:1116–8.

Khurana N, Yaylali E, Farnham PG, Hicks KA, Allaire BT, Jacobson E, Sansom SL. Impact of Improved HIV Care and Treatment on PrEP Effectiveness in the United States, 2016–2020. *JAIDS Journal of Acquired Immune Deficiency Syndromes.* 2018 Aug 1;78(4):399-405.

Kochanek KD, Xu J, Arias E. Mortality in the United States, 2019. NCHS data brief, no 395. Hyattsville, MD: National Center for Health Statistics. 2020.

Krebs, E., Enns, B., Wang, L., Zang, X., Panagiotoglou, D., Del Rio, C., ... & Marshall, B. (2019). Developing a dynamic HIV transmission model for 6 US cities: An evidence synthesis. PloS one, 14(5), e0217559.

Krentz, H. B., MacDonald, J., & John Gill, M. (2014, March). High mortality among human immunodeficiency virus (HIV)-infected individuals before accessing or linking to HIV care: a missing outcome in the cascade of care?. In Open forum infectious diseases (Vol. 1, No. 1). Oxford University Press.

Krueger, A. L., Van Handel, M., Dietz, P. M., Williams, W. O., Johnson, A. S., Klein, P. W., ... & Purcell, D. W. (2019). Factors Associated with State Variation in Mortality Among Persons Living with Diagnosed HIV Infection. Journal of Community Health, 1-11.

Landovitz RJ, Donnell D, Clement ME, et al. Cabotegravir for HIV Prevention in Cisgender Men and Transgender Women. N Engl J Med. 2021;385(7):595-608. doi:10.1056/NEJMoa2101016

Lansky, A., Johnson, C., Oraka, E., Sionean, C., Joyce, M. P., DiNenno, E., & Crepaz, N. (2015). Estimating the Number of Heterosexual Persons in the United States to Calculate National Rates of HIV Infection. PloS one, 10(7), e0133543. https://doi.org/10.1371/journal.pone.0133543

Leichliter JS, Chesson HW, Sternberg M, et al. The concentration of sexual behaviours in the USA: a closer examination of subpopulations. *Sex Transm Infect*. 2010;86(Suppl 3):iii45–51.

Leynaert B, Downs AM, de Vincenzi I; European Study Group on Heterosexual Transmission of HIV. Heterosexual transmission of human immunodeficiency virus: variability of infectivity throughout the course of infection. *Am J Epidemiol*. 1998;148:88–96.

Lin F, Farnham PG, Shrestha RK, Mermin J, Sansom SL. Cost Effectiveness of HIV Prevention Interventions in the U.S. Am J Prev Med. 2016;50(6):699-708. doi:10.1016/j.amepre.2016.01.011

Long E, Brandeau M, Owens D. The cost-effectiveness and population outcomes of expanded HIV screening and antiretroviral treatment in the United States. *Ann Intern Med*. 2010;153:778–89.

Malekinejad M, Blodgett J, Horvath H, et al. (2021) Change in Condom Use in Populations Newly Aware of HIV Diagnosis in the United States and Canada: A Systematic Review and Meta-Analysis. AIDS Behav. 2021 Jan 2. doi: 10.1007/s10461-020-03113-8. Epub ahead of print. PMID: 33389321.

Morgan E, Ryan DT, Newcomb ME, Mustanski B. High Rate of Discontinuation May Diminish PrEP Coverage Among Young Men Who Have Sex with Men. AIDS Behav. 2018 Nov;22(11):3645-3648. doi: 10.1007/s10461-018-2125-2. PMID: 29728950; PMCID: PMC6204096.

North American AIDS Cohort Collaboration on Research and Design (NA-ACCORD). Mortality estimates 2010 v2. Data on file. April 29, 2014.

OraQuick ADVANCE® [Package Insert]. Bethlehem, PA: OraSure Technologies; 2004

Osmond D, Bacchetti P, Chaisson RE, et al. Time of exposure and risk of HIV infection in homosexual partners of men with AIDS. *Am J Public Health*. 1988;78:944–8.

Patel D, Williams WO, Wright C, et al. (2022) HIV Testing Services Outcomes in CDC-Funded Health Departments During COVID-19. J Acquir Immune Defic Syndr. 2022;91(2):117-121. doi:10.1097/QAI.0000000000003049

Patel P, Borkowf CB, Brooks JT, Lasry A, Lansky A, Mermin J. Estimating per-act HIV transmission risk: a systematic review. *AIDS*. 2014 Jun 19;28(10):1509-19

Pilcher CD, Louie B, Facente S, et al. Performance of rapid point-of-care and laboratory tests for acute and established HIV infection in San Francisco. *PLoS One*. 2013 Dec 12;8(12):e80629.

Pinkerton S, Abramson P, Holtgrave D, editors. The Bernoulli-process model of HIV transmission: applications and implications. In: *Handbook of economic evaluation of HIV prevention programs*. New York, NY: Plenum Press; 1998.

Porco TC, Martin JN, Page-Shafer KA, et al. Decline in HIV infectivity following the introduction of highly active antiretroviral therapy. *AIDS*. 2004;18:81–8.

Reece M, Herbenick D, Schick V, et al. Sexual behaviors, relationships, and perceived health among adult men in the United States: results from a national probability sample. *J Sex Med*. 2010a;7:291–304.

Reece M, Herbenick D, Schick V, et al. Condom use rates in a national probability sample of males and females aged 14 to 94 in the United States. *J Sex Med*. 2010b;7(Suppl 5):266–76.

Rodger AJ, Cambiano V, Bruun T, et al.(2016); PARTNER Study Group. Sexual activity without condoms and risk of HIV transmission in serodifferent couples when the HIV-positive partner is using suppressive antiretroviral therapy. JAMA 2016;316:171–81. 10.1001/jama.2016.5148

Rodger AJ, Cambiano V, Bruun T, et al. (2019); PARTNER Study Group. Risk of HIV transmission through condomless sex in serodifferent gay couples with the HIV-positive partner taking suppressive antiretroviral therapy (PARTNER): final results of a multicentre, prospective, observational study. Lancet. 2019 Jun 15;393(10189):2428-2438. doi: 10.1016/S0140-6736(19)30418-0

Rosenberg ES, Purcell DW, Grey JA, Hankin-Wei A, Hall E, Sullivan PS. Rates of prevalent and new HIV diagnoses by race and ethnicity among men who have sex with men, US states, 2013–2014. Annals of epidemiology. 2018 Apr 30.

Sabin CA, Smith CJ, Delpech V, Anderson J, Bansi L, Gilson R, Schwenk A, Leen C, Gazzard B, Porter K, Mackie N. The associations between age and the development of laboratory abnormalities and treatment discontinuation for reasons other than virological failure in the first year of highly active antiretroviral therapy. *HIV Medicine*. 2009 Jan;10(1):35-43.

Siegfried N, Muller M, Deeks JJ, et al. Male circumcision for prevention of heterosexual acquisition of HIV in men. *Cochrane Database Syst Rev*. 2009;2:CD003362. doi: 10.1002/14651858.CD003362.pub2

Smith DK, Herbst JH, Zhang X, et al. Condom effectiveness for HIV prevention by consistency of use among men who have sex with men in the United States. *J Acquir Immune Defic Syndr*. 2015 Mar 1;68(3):337–44.

Sorensen SW, Sansom SL, Brooks JT, et al. A mathematical model of comprehensive test-and-treat services and HIV incidence among men who have sex with men in the United States. *PLoS One*. 2012;7:e29098.

Tempalski B, Pouget ER, Cleland CM, Brady JE, Cooper HL, Hall HI, Lansky A, West BS, Friedman SR. Trends in the population prevalence of people who inject drugs in US metropolitan areas 1992–2007. *PloS One*. 2013 Jun 5;8(6):e64789.

Teshale EH, Asher A, Aslam MV, Augustine R, Duncan E, Rose-Wood A, Ward J, Mermin J, Owusu-Edusei K, Dietz PM. Estimated cost of comprehensive syringe service program in the United States. PloS one. 2019 Apr 26;14(4):e0216205.

Tian Y, Hassmiller Lich K, et al. Linked sensitivity analysis, calibration, and uncertainty analysis using a system dynamics model for stroke comparative effectiveness research. *Med Decis Making*. 2016 Apr 18;pii: 0272989X16643940. [Epub ahead of print]

U.S. Census Bureau. Monthly Estimates of the Resident Population by Sex, Race Alone or in Combination, and Hispanic Origin for the United States, States, and Counties: July 1, 2018 to Dec 1, 2018 Source: U.S. Census Bureau. Accessed March 16, 2020.

U.S. Census Bureau. Monthly Estimates of the Resident Population by Sex, Race Alone or in Combination, and Hispanic Origin for the United States: April 1, 2010 to July 1, 2019. U.S. Census Bureau, Population Division. June, 2020. Accessed June 11, 2021

U.S. Census Bureau. Annual Estimates of the Resident Population by Sex, Single Year of Age, Race, and Hispanic Origin for the United States: April 1, 2010 to July 1, 2016. U.S. Census Bureau, Population Division. June 2017. Available at: https://www.census.gov/programs-surveys/popest.html. Accessed September 11, 2018.

U.S. Census Bureau. Census 2010, Summary file 1, Tables QT-P2, PCT12J, and PCT12H. 2010. Available at: <http://factfinder2.census.gov>. Accessed March 20, 2014.

U.S. Department of Health and Human Services. 2022. America's HIV Epidemic Analysis Dashboard (AHEAD). https://ahead.hiv.gov/. Accessed 12/8/22.

Viguerie, A., Farnham, P.G., Lyles, C. McGowan, R., Sullivan, P., & Sanchez, T. Analysis of iSTAMP Data (CDC Internal report; not for distribution without written permission of the iSTAMP project team). 2023. Report shared with RTI on February 24, 2023.

Viguerie A, Song R, Johnson AS, Lyles CM, Hernandez A, Farnham PG. Isolating the effect of COVID-19 related disruptions on HIV diagnoses in the United States in 2020. JAIDS Journal of Acquired Immune Deficiency Syndromes. 2022 May 13:10-97.

Vlahov D, Wang C, Ompad D, et al. Mortality risk among recent-onset injection drug users in five U.S. cities. *Substance Use & Misuse*. 2008;43:413-28.

Wall SD, Olcott EW, Gerberding JL. AIDS risk and risk reduction in the radiology department. *Am J Roentgenol*. 1991;157:911–7.

Wall KM, Stephenson R, & Sullivan PS. Frequency of Sexual Activity With Most Recent Male Partner Among Young, Internet-Using Men Who Have Sex With Men in the United States. Journal of Homosexuality, 60(10), 1520–1538. http://doi.org/10.1080/00918369.2013.819256.

Wawer MJ, Gray RH, Sewankambo NK, et al. Rates of HIV-1 transmission per coital act, by stage of HIV-1 infection, in Rakai, Uganda. *J Infect Dis*. 2005;191:1403–19.

Weller S, Davis K. Condom effectiveness in reducing heterosexual HIV transmission. *Cochrane Database Syst Rev*. 2002;1:CD003255.

Woodring J, Kruszon-Moran D, McQuillan G. (2015) HIV infection in U.S. household population aged 18–59: Data from National Health and Nutrition Examination Survey, 2007–2012. National health statistics reports; no 83. Hyattsville, MD: National Center for Health Statistics. 2015

Wu L, Schumacher C, Chandran A, et al. Patterns of PrEP Retention Among HIV Pre-exposure Prophylaxis Users in Baltimore City, Maryland. J Acquir Immune Defic Syndr. 2020;85(5):593-600. doi:10.1097/QAI.0000000000002506

Zaric GS, Barnett PG, Brandeau ML. HIV transmission and the cost-effectiveness of methadone maintenance. Am J Public Health. 2000; 90(7):1100–1111. [PubMed: 10897189]

Zhu W, Huang YA, Weiner J, Neblett-Fanfair R, Kourtis AP, Hall HI, Hoover KW. Impact of the COVID–19 pandemic on prescriptions for antiretroviral drugs for HIV treatment in the United States, 2019–2021, AIDS: July 15, 2022 - Volume - Issue - 10.1097/QAD.0000000000003315 doi: 10.1097/QAD.0000000000003315

Appendix A:
Definitions

Table A.1.	Definitions of Symbols Applied in This Document

| Symbol | Definition |
| --- | --- |
| **Latin Alphabet** | |
| $a_{p1,p2}$ | Percentage in subpopulation p1’s partners that are in p2, as determined by the mixing matrix |
| *B^c^* | Rate of HIV progression to the next disease stage from compartment *c,* if VLS; applies to individuals with HIV, CD4 ≥ 200, and VLS, (*c* = 13, 18, 23) only |
| *b_p1,p2,z_* | Percentage reduction in per-insertive sex act transmission probability from an act of type z due to circumcision for an individual without HIV in subpopulation *p1* with an partner with HIV in subpopulation *p2* |
| $\ddot{C}_{t}^{s}$ | Effect of COVID pandemic type s at time t |
| *d_z_* | Percentage reduction in per-sex-act transmission probability from an act of type z due to condom use |
| *E* | Number of needles shared annually per needle-sharing partner by a person who injects drugs (PWID) who has never been diagnosed with HIV |
| $e_{p}(t)$ | Rate of departure from HIV care if in care but not on ART, by demographic subpopulation *p* at time *t*; applies to (*c =* 8, 11, 16, 21, 26) only |
| *F* | Cumulative number of HIV infections over the time horizon |
| *f_p_* | Relative adjustment to rate of aging into the youngest age group of the model for all p in the youngest age group (j = 1 if the youngest age group is 13–17 and j = 2 if youngest age group is 18 to 24) |
| ḟ_t_ | Year number of time step *t* (1 to Ṫ) |
| *G_z,p,c_* | Percentage of sex acts of risk type z protected with a condom, given partner in compartment *c*, by subpopulation *p* |
| $H_{p}^{c}$ | Rate of HIV progression to the next disease stage from compartment *c,* if on ART, not VLS, by subpopulation p; applies to individuals with HIV, CD4 ≥ 200, and on ART, not VLS (*c* = 12, 17, 22) only |
| *i*_p_ | Percentage reduction in the annual rate of HIV transmission if HIV- individual in subpopulation *p* is on PrEP |
| *L* | Cumulative life-years in the modelled population over the time horizon |
| ${}_{p}^{c}(t)$ | Rate of linkage to HIV care among aware (not newly diagnosed) individuals, by demographic subpopulation *p* at time *t*; applies to individuals aware of their infection but not in care or on ART (*c* =7, 10, 15, 20, 25) only |
| $M_{p}^{z}$ | Annual number of partners of risk type z per person in demographic subpopulation *p* |
| m_p_^z^ | Number of partners of risk type z per individual in subpopulation p |
| $O_{p,y}$ | Percentage of subpopulation p’s partnerships that are type y, given partnerships of type y |

(continued)

Table A.1.	Definitions of Symbols Applied in This Document (continued)

| Symbol | Definition |
| --- | --- |
| **Latin Alphabet (continued)** | |
| *P(t)* | HIV prevalence at time *t*, defined as the number of individuals with HIV |
| *Q* | Discount rate for QALYs |
| *q(t)* | Length of model’s computational time step, in years, at time t |
| *R_c_* | Health utility for individuals in compartment c |
| *S _p_* | Number of annual sex acts for an individual without HIV per partner, HIV-uninfected or undiagnosed, by subpopulation *p* |
| *s_c_* | Binary indicator that model state *c* is an infected HIV state (1 = infected, 0 = not infected) |
| *Τ*_p1,p2,c_ | Reduction in number of needles shared for diagnosed versus undiagnosed or uninfected (0 if compartment *c* is for undiagnosed or uninfected compartments or if either subpopulations *p1* or *p2* are not PWID) |
| $u$ | Percentage of the individuals who become VLS among those who initiate ART |
| $V_{p1,p2}$ | Proportion of male-male sex acts by individuals in subpopulation p1 with individuals in subpopulation p2 that are receptive (0 if subpopulation p1 or p2 is not MSM), so that (1 − $V_{p1,p2}$) is the fraction of insertive sex acts |
| W | Cumulative quality-adjusted life-years in the modelled population over the time horizon |
| $w_{c,g}$ | Test sensitivity by compartment *c* and type of test *g* |
| $Y_{a*}^{N}$ | Annual probability of stopping PrEP, if susceptible and on-PrEP for PrEP type N and PrEP adherence level a* |
| **Greek Alphabet** | |
| $\alpha_{p}(t)$ | Percentage of PrEP initiators on oral (versus injectable) PrEP, by subpopulation p at time t |
| $\beta_{p1,p2,t}^{z,y,c}$ | Probability of transmission at time t for an individual without HIV in subpopulation *p1* per sexual or needle-sharing partnership from risk type *z* (vaginal, anal, or needle) in a partnership type *y* (male-female partnership with vaginal intercourse only, male-male partnership with anal intercourse only, male-female partnership that includes anal intercourse, or needle-sharing) with a partner who is in subpopulation *p2* and compartment *c* |
| $\Gamma_{c,z}$ | Per-sex-act transmission probability for insertive condomless sex (act) of type z (vaginal or anal intercourse) with infected partner in compartment *c* |
| $\Upsilon_{p}^{N}$ | Percentage of PrEP initiators with high (versus low) adherence, by subpopulation p and PrEP type N |
| $\alpha_{p}(t)$ | Percentage of PrEP initiators on oral (versus injectable) PrEP, by subpopulation p at time t |

(continued)

Table A.1.	Definitions of Symbols Applied in This Document (continued)

| Symbol | Definition |
| --- | --- |
| **Greek Alphabet** | |
| $\beta_{p1,p2,t}^{z,y,c}$ | Probability of transmission at time t for an individual without HIV in subpopulation *p1* per sexual or needle-sharing partnership from risk type *z* (vaginal, anal, or needle) in a partnership type *y* (male-female partnership with vaginal intercourse only, male-male partnership with anal intercourse only, male-female partnership that includes anal intercourse, or needle-sharing) with a partner who is in subpopulation *p2* and compartment *c* |
| $\Gamma_{c,z}$ | Per-sex-act transmission probability for insertive condomless sex (act) of type z (vaginal or anal intercourse) with infected partner in compartment *c* |
| $\Upsilon_{p}^{N}$ | Percentage of PrEP initiators with high (versus low) adherence, by subpopulation p and PrEP type N |
| $\gamma_{p}^{c}(t)$ | Rate of ART initiation if linked to HIV care, by compartment c and subpopulation *p* at time *t*; applies to individuals linked to HIV care and not on ART (*c =* 8, 11, 16, 21, 26) |
| $\Delta_{c,z}$ | Percentage reduction in per-act transmission probability due to viral load suppression, by compartment c and transmission risk type *z* |
| $\delta_{p}^{+}$and$\delta_{p}^{-}$ | Aging rates into (+) and out of (–) demographic subpopulation *p* |
| $\epsilon_{p}(t)$ | Percentage of screens that are rapid (type of test g = 1) overall, by subpopulation p at time t |
| $\epsilon_{p}^{v}(t)$ | Percentage of screens that are rapid (type of test g = 1) in setting v, by subpopulation p at time t |
| $\varepsilon_{p}(t)$ | Annual rate of dropping off of ART if ART-not-VLS, by demographic subpopulation *p* at time *t*; applies to individuals who are ART-not-VLS (*c =* 12, 17, 22, 27) only |
| $\eta_{p}(t)$ | Annual rate of loss of viral load suppression if VLS, by demographic subpopulation *p* at time *t*; applies to individuals with HIV and who are VLS (*c* = 13, 18, 23, 28) only |
| Θ *_c_* | Probability of HIV transmission per needle shared with a Partner with HIV in compartment c |
| $\vartheta_{p}^{c}$ | Rate of HIV regression to the previous disease stage from compartment *c*, if VLS, by subpopulation p; applies to individuals with HIV, CD4 ≤500, and VLS, (*c* = 18, 23, 28) only |
| $\kappa_{p}(t)$ | Percentage of newly diagnosed individuals in subpopulation *p* immediately linked to HIV care at time *t* |
| $\Lambda_{p}$ | Constant rate of aging into the modeled population per person in subpopulation *p* at the start of the model |
| $\lambda_{p}(t)$ | Force of HIV infection for people without HIV (across all sexual and needle-sharing risks) in subpopulation *p* at time *t* |

(continued)

Table A.1.	Definitions of Symbols Applied in This Document (continued)

| Symbol | Definition |
| --- | --- |
| **Greek Alphabet (continued)** | |
| $\lambda_{p}^{z,x}(t)$ | Force of HIV infection from risk type *z* for people without HIV in subpopulation *p* who participate in sexual transmission risk behaviors of each type x, at time *t.* It is a function of the probability of infection from partners with HIV, percentage of partnerships that include risk type *z*, number of partners with HIV, and the distribution of partners across HIV stages and the continuum of care. |
| $\mu_{p}$ | Mortality rate among persons without HIV |
| ξ _z,y_,_p1,p2,c_ | Number of partnerships of type y involving risk type z per individual without HIV in subpopulation p1 with infected partners in subpopulation p2 in compartment c. |
| $\text{π}_{\text{p}}^{\text{c}}$(t) | Rate of testing of undiagnosed individuals in compartment *c*, at time *t* by demographic subpopulation *p* |
| $\text{ϖ}_{\text{p,g}}$(t) | Probability of notification given a confirmed positive test result for a previously undiagnosed individual in demographic subpopulation *p* and type of test *g* at time *t* |
| $\varrho_{p,x}$ | Proportion of subpopulation p with transmission risk participation type x, which is defined by the percentage of people who have AI in their male-female partnerships as well as the assumptions that 100% of MSM have AI in their male-male partnerships and 100% of all transmission groups have VI in their male-female partnerships |
| $\sigma_{p}^{c}(t)$ | Mortality rate if HIV-infected, by demographic subpopulation *p* and compartment *c,* at time *t* |
| $\tau_{p}^{c}(t)$ | Diagnosis rate based on test and notification of unaware PWH in compartment *c*, progressing them from unaware (*r =* 1) to aware (*r* = 2 or *r =* 3), by subpopulation *p* |
| $\Phi_{c,z}$ | Per-sex-act transmission probability for receptive condomless intercourse of type z (vaginal or anal intercourse) with infected partner in compartment *c* |
| $\varphi_{v}$ | Percentage of screens that occur in setting *v* (clinical or non-clinical) |
| $\Psi_{p}\left( t \right)$ | Annual probability of initiating PrEP, given eligible, for subpopulation *p* at time *t* |
| $\Xi_{a*}^{N}$ | Multiplier on efficacy of PrEP to account for less than daily adherence, by PrEP type N and adherence level a* |
| Ω_p1,p2,z,y_ | Proportion of sexual acts by individuals in subpopulation p1 in partnerships of type y with individuals in subpopulation p2 that are risk type z (where z = vaginal or anal) |
| $\omega_{p}^{c}$ | Rate of HIV progression to the next disease stage from compartment *c*, by subpopulation p; applies to individuals with HIV, CD4 ≥ 200, and not on ART (*c* = 6 to 8, 9 to 11, 14 to 16, 19 to 21) only |
| $\zeta_{p}\left( t \right)$ | Annual rate of becoming VLS if ART-not-VLS, by demographic subpopulation *p* at time *t* |

Note: AI = anal intercourse; ART = antiretroviral therapy; PrEP = pre-exposure prophylaxis; PWID = people who inject drugs; VI = vaginal intercourse; VLS = viral load suppression.

Table A.2.	Definitions of Indices Applied in This Document

| Index | | Definition | Number of Categories | | | Categories (Represented by) |
| --- | --- | --- | --- | --- | --- | --- |
| a* | | PrEP adherence level | 2 | | | High adherence (1)  Low adherence (2) |
| c | | Compartment | 30 | | | 28 main compartments for individuals actively moving through the model (further defined by disease stage [*h*] and continuum-of-care stage [*r*]) and 2 compartments for individuals who were no longer actively followed in the model due to death among persons with CD4≥200 and death among persons with CD4<200.  1: People without HIV / not on PrEP (A1)  2: People without HIV / on oral PrEP / high adherence (A6)  3: People without HIV / on oral PrEP / low adherence (A7)  4: People without HIV / on injectable PrEP / high adherence (A8)  5: People without HIV / on injectable PrEP / low adherence (A9)  6: PWH / acute stage / unaware of infection (B1)  7: PWH / acute stage / aware, but not linked to HIV care (B2)  8: PWH / acute stage / linked to HIV care, but not on ART (B3)  9: PWH / CD4>500 / unaware of infection (C1)  10: PWH / CD4>500 / aware, but not linked to HIV care (C2)  11: PWH / CD4>500 / linked to HIV care, but not on ART (C3)  12: PWH / CD4>500 / on ART, not VLS (C4)  13: PWH / CD4>500 / VLS (C5)  14: PWH / CD4 350–500 / unaware of infection (D1)  15: PWH / CD4 350–500 / aware, but not linked to HIV care (D2)  16: PWH / CD4 350–500 / linked to HIV care, but not on ART (D3)  17: PWH / CD4 350–500 / on ART, not VLS (D4)  18: PWH / CD4 350–500 / VLS (D5)  19: PWH / CD4 200–350 / unaware of infection (E1)  20: PWH / CD4 200–350 / aware, but not linked to HIV care (E2)  21: PWH / CD4 200–350 / linked to HIV care, but not on ART (E3)  22: PWH / CD4 200–350 / on ART, not VLS (E4)  23: PWH / CD4 200–350 / VLS (E5)  24: PWH / CD4 < 200 / unaware of infection (F1)  25: PWH / CD4 < 200 / aware, but not linked to HIV care (F2)  26: PWH / CD4 < 200 / linked to HIV care, but not on ART (F3)  27: PWH / CD4 < 200 / on ART, not VLS (F4)  28: PWH / CD4 <200 / VLS (F5)  29: Death among persons with CD4≥200  30: Death among persons with CD4<200 |
| g | | Test type | 2 | | | Rapid screen (1)  Conventional screen (2) |
| h | | Disease status | 6 | | | People without HIV (0)  PWH with acute HIV infection (1)  PWH with CD4 count greater than 500 cells/mm^3^ (2)  PWH with CD4 count between 350 cells/mm^3^ and 500 cells/mm3 (3)  PWH with CD4 count between 200 cells/mm^3^ and 350 cells/mm3 (4)  PWH with CD4 count less than 200 cells/mm^3^ (5) |
| j | | Age group | 7 | | | 13–17 years (1)  18–24 years (2)  25–34 years (3)  35–44 years (4)  45–54 years (5)  55–64 years (6)  65+ years (7) |
| k | Number of HIV transmission risk factors | | | 2 | Fewer (1)  Multiple (2) | |
| l | Transmission group | | | 3 | HETs (1)  MSM (2)  PWID (3) | |
| m | Sex | | | 2 | Male (1)  Female (2) | |
| N | PrEP type | | | 2 | Oral PrEP (1)  Injectable PrEP (2) | |

(continued)

Table A.2.	Definitions of Indices Applied in This Document (continued)

| Index | Definition | Number of Categories | Categories (Represented by) |
| --- | --- | --- | --- |
| n | Circumcision status | 2 | Uncircumcised (1)  Circumcised (2) |
| o | Race/ethnicity | 3 | Black (1)  Hispanic/Latino (2)  White/other (3) |
| p | Subpopulation | 273 | Combinations defined by age group (j), number of HIV transmission risk factors (k), transmission group (l), sex (m), circumcision status (n), and race/ethnicity (o). PWID all have multiple HIV transmission risk factors, MSM are all male, and only males are stratified by circumcision status, therefore not all combinations of these stratifications are represented. |
| q | Allocation period | 3 | Allocation period 1 (1)  Allocation period 2 (2)  Allocation period 3 (3) |
| r | Care-Continuum stage | 5 | People without HIV or PWH unaware of infection (1)  PWH aware of infection, not on ART or in HIV care (2)  PWH linked to HIV care, not on ART (3)  PWH on ART, not VLS (4)  PWH with VLS (5) |
| s | Type of COVID effect | 8 | COVID effects on testing rates of people without HIV (1) and PWH (2)  COVID effects on ART initiation rates (3)  COVID effects on rates of dropping off of ART (4) and losing VLS (5)  COVID effects on PrEP initiation rates (6)  COVID effects on numbers of sexual contacts (7)  COVID effects on mortality rates (8) |
| t | Time/timestep | 1 | -- |
| v | Setting for HIV screen | 2 | Clinical (1)  Nonclinical (2) |
| x | Sexual transmission risk participation type | 3 | Vaginal intercourse only in male-female partnerships (1)  Anal intercourse only in male-male partnerships (2)  Anal intercourse in male-female partnerships (3) |

(continued)

Table A.2.	Definitions of Indices Applied in This Document (continued)

| Index | Definition | Number of Categories | Categories (Represented by) |
| --- | --- | --- | --- |
| y | Partnership type | 4 | Male-female partnership that only includes vaginal intercourse (1)  Male-male partnership that only includes anal intercourse (2)  Male-female partnership that includes anal intercourse (3)  Needle-sharing (4) |
| z | Transmission risk type | 3 | Vaginal intercourse (1)  Anal intercourse (2)  Needle-sharing (3) |

Note: ART = antiretroviral therapy; PrEP = pre-exposure prophylaxis; PWH = people with HIV; PWID = people who inject drugs; VLS = viral suppression
